# A Serology Strategy for Epidemiological Studies Based on the Comparison of the Performance of Seven Different Test Systems - The Representative COVID-19 Cohort Munich

**DOI:** 10.1101/2021.01.13.21249735

**Authors:** Laura Olbrich, Noemi Castelletti, Yannik Schälte, Mercè Garí, Peter Pütz, Abhishek Bakuli, Michael Pritsch, Inge Kroidl, Elmar Saathoff, Jessica Michelle Guggenbuehl Noller, Volker Fingerle, Ronan Le Gleut, Leonard Gilberg, Isabel Brand, Philine Falk, Alisa Markgraf, Flora Deák, Friedrich Riess, Max Diefenbach, Tabea Eser, Franz Weinauer, Silke Martin, Ernst-Markus Quenzel, Marc Becker, Jürgen Durner, Philipp Girl, Katharina Müller, Katja Radon, Christiane Fuchs, Roman Wölfel, Jan Hasenauer, Michael Hoelscher, Andreas Wieser, On behalf of the KoCo19-Study Team

## Abstract

**Background:** Serosurveys are essential to understand SARS-CoV-2 exposure and enable population-level surveillance, but currently available tests need further in-depth evaluation. We aimed to identify testing-strategies by comparing seven seroassays in a population-based cohort.

**Methods:** We analysed 6,658 samples consisting of true-positives (n=193), true-negatives (n=1,091), and specimens of unknown status (n=5,374). For primary testing, we used Euroimmun-Anti-SARS-CoV-2-ELISA-IgA/IgG and Roche-Elecsys-Anti-SARS-CoV-2; and virus-neutralisation, GeneScript®cPass™, VIRAMED-SARS-CoV-2-ViraChip®, and Mikrogen-*recom*Line-SARS-CoV-2-IgG, including common-cold CoVs, for confirmatory testing. Statistical modelling generated optimised assay cut-off-thresholds.

**Findings:** Sensitivity of Euroimmun-anti-S1-IgA was 64.8%, specificity 93.3%; for Euroimmun-anti-S1-IgG, sensitivity was 77.2/79.8% (manufacturer’s/optimised cut-offs), specificity 98.0/97.8%; Roche-anti-N sensitivity was 85.5/88.6%, specificity 99.8/99.7%. In true-positives, mean and median titres remained stable for at least 90-120 days after RT-PCR-positivity. Of true-positives with positive RT-PCR (<30 days), 6.7% did not mount detectable seroresponses. Virus-neutralisation was 73.8% sensitive, 100.0% specific (1:10 dilution). Neutralisation surrogate tests (GeneScript®cPass™, Mikrogen-*recom*Line-RBD) were >94.9% sensitive, >98.1% specific. Seasonality had limited effects; cross-reactivity with common-cold CoVs 229E and NL63 in SARS-CoV-2 true-positives was significant.

**Conclusion:** Optimised cut-offs improved test performances of several tests. Non-reactive serology in true-positives was uncommon. For epidemiological purposes, confirmatory testing with virus-neutralisation may be replaced with GeneScript®cPass™ or *recom*Line-RBD. Head-to-head comparisons given here aim to contribute to the refinement of testing-strategies for individual and public health use.

## Introduction

In December 2019, a cluster of atypical pneumonia of unknown origin was described in the region of Wuhan, Hubei province, China. Subsequently, a novel coronavirus was identified as the causative pathogen: SARS-CoV-2 (Severe Acute Respiratory Syndrome Coronavirus 2).(1) As the virus spread rapidly across the globe, the Corona Virus Disease 2019 (COVID-19) was declared a pandemic on March 12^th^, 2020.

Direct detection of viral nucleic acids or the virus itself in bodily fluids is considered the reference standard of acute infection; primarily identified through the use of nasopharyngeal swabs or other respiratory probes.(2) Additionally, serodiagnostics are valuable in order firstly to identify past infections, asymptomatic or symptomatic, and secondly to elucidate transmission dynamics within the population; both being highly relevant to inform evidence-based political decision making.(3, 4)

Several serological test systems have been introduced since the beginning of the pandemic.(5) Most target one of two specific viral structures: parts of the trimeric CoV spike (S1-2) complex or the nucleocapsid (N) envelope protein.(6) While the receptor binding domain (RBD) of S1 folds out to bind to the angiotensin-converting enzyme 2 (ACE2) receptor, the N-protein is involved in viral assembly and replication.(7) To select the most suitable system for different applications, independent evaluations and comparisons of their sensitivities and specificities on different cohorts are of utmost importance; yet, as Cheng et al. state, validations of these tests are often poorly described and evaluations show several shortcomings.^(7)^ Firstly, as most validation studies have been performed with severely sick subjects, proposed cut-off values are ambiguous.(4, 8) Those individuals will likely have higher titres than asymptomatic subjects,(6, 9-12) possibly resulting in a less reliable performance of the test system, or the potential need for different cut-off values in these populations.(13) Secondly, it remains unclear whether the antibodies are serocorrelates of protection, highlighting the gap in knowledge of the underlying biology.(3, 7) And lastly, cross-reactivity with other CoVs has been reported, but its effects on the test readouts remain uncertain.(14-16)

Here we present the serological testing systems applied within the Representative COVID-19 Cohort Munich (KoCo19), a prospective sero-incidence study initiated in Munich, Germany, in April 2020.(17, 18) Three independent primary tests (Euroimmun Anti-S1-SARS-CoV-2-ELISA-IgA & -IgG and Elecsys Anti-SARS-CoV-2 Roche N pan-Ig) and a number of confirmatory tests (direct viral neutralisation, GeneScript®cPass™, Mikrogen-*recom*Line-RBD IgG line immunoassay, VIRAMED-SARS-CoV-2-ViraChip® microarray) were assessed in a head-to-head cross-comparison. The tests were conducted on samples from (i.) RT-PCR positive individuals (true-positives), (ii.) blood donors from the pre-COVID-19 era (true-negatives), and (iii.) subjects with unknown disease status (unknown). Thus, we were able to generate reliable performance estimates for both primary and confirmatory tests by using true-positive and true-negative individuals from the aforementioned cohorts and to subsequently derive optimised cut-offs for our cohort.

## Methods

### Study design and participants

In total, we included 6,658 samples, derived from a set of SARS-CoV-2 RT-PCR positives (“true-positives”, n=193), individuals from historical cohorts, blood donors without any indication of SARS-CoV-2 infection (“true-negatives”, n=1,091), and specimen of unknown status (n=5,374); details on the cohort characteristics, including collection-timepoints, can be found in the appendix (p.1; p.4 supplemental table 1).

**Table 1:**
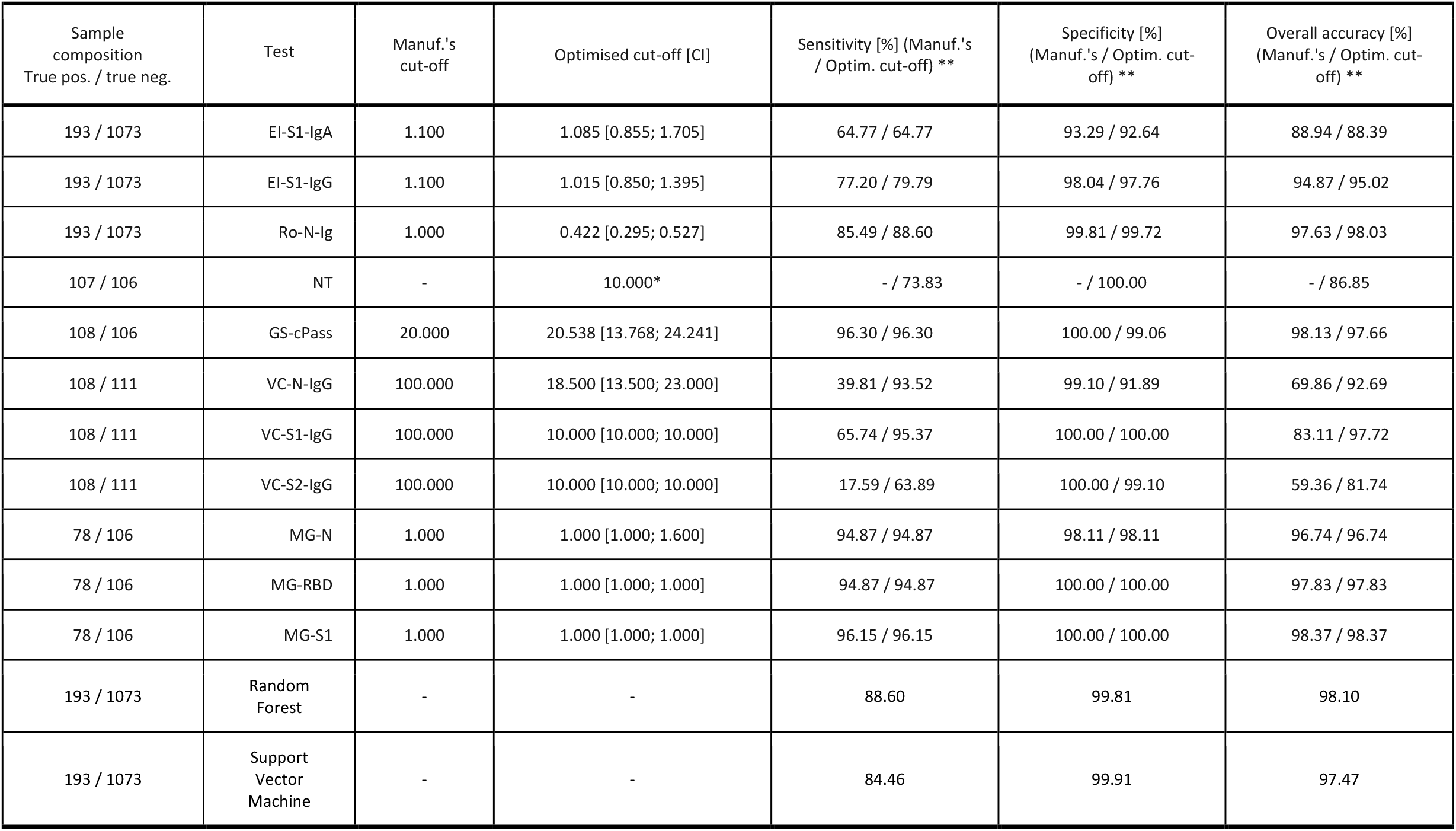
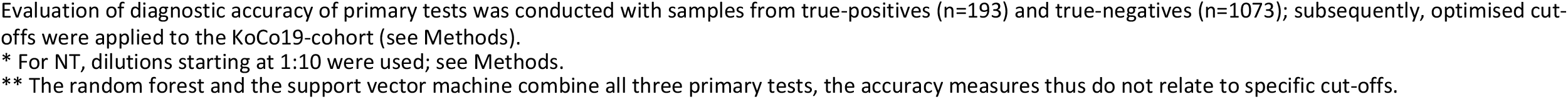
Manufacturer’s and optimised cut-off, sensitivity, specificity and accuracy

| Sample composition<br>True pos. / true neg. | Test | Manuf.'s cut-off | Optimised cut-off [CI] | Sensitivity [%] (Manuf.'s / Optim. cut-off) ** | Specificity [%] (Manuf.'s / Optim. cut-off) ** | Overall accuracy [%] (Manuf.'s / Optim. cut-off) ** |
| --- | --- | --- | --- | --- | --- | --- |
| 193 / 1073 | El-S1-IgA | 1.100 | 1.085 [0.855; 1.705] | 64.77 / 64.77 | 93.29 / 92.64 | 88.94 / 88.39 |
| 193 / 1073 | El-S1-IgG | 1.100 | 1.015 [0.850; 1.395] | 77.20 / 79.79 | 98.04 / 97.76 | 94.87 / 95.02 |
| 193 / 1073 | Ro-N-Ig | 1.000 | 0.422 [0.295; 0.527] | 85.49 / 88.60 | 99.81 / 99.72 | 97.63 / 98.03 |
| 107 / 106 | NT | - | 10.000* | - / 73.83 | - / 100.00 | - / 86.85 |
| 108 / 106 | GS-cPass | 20.000 | 20.538 [13.768; 24.241] | 96.30 / 96.30 | 100.00 / 99.06 | 98.13 / 97.66 |
| 108 / 111 | VC-N-IgG | 100.000 | 18.500 [13.500; 23.000] | 39.81 / 93.52 | 99.10 / 91.89 | 69.86 / 92.69 |
| 108 / 111 | VC-S1-IgG | 100.000 | 10.000 [10.000; 10.000] | 65.74 / 95.37 | 100.00 / 100.00 | 83.11 / 97.72 |
| 108 / 111 | VC-S2-IgG | 100.000 | 10.000 [10.000; 10.000] | 17.59 / 63.89 | 100.00 / 99.10 | 59.36 / 81.74 |
| 78 / 106 | MG-N | 1.000 | 1.000 [1.000; 1.600] | 94.87 / 94.87 | 98.11 / 98.11 | 96.74 / 96.74 |
| 78 / 106 | MG-RBD | 1.000 | 1.000 [1.000; 1.000] | 94.87 / 94.87 | 100.00 / 100.00 | 97.83 / 97.83 |
| 78 / 106 | MG-S1 | 1.000 | 1.000 [1.000; 1.000] | 96.15 / 96.15 | 100.00 / 100.00 | 98.37 / 98.37 |
| 193 / 1073 | Random Forest | - | - | 88.60 | 99.81 | 98.10 |
| 193 / 1073 | Support Vector Machine | - | - | 84.46 | 99.91 | 97.47 |

Evaluation of diagnostic accuracy of primary tests was conducted with samples from true-positives (n=193) and true-negatives (n=1073); subsequently, optimised cut-offs were applied to the KoCo19-cohort (see Methods).
\* For NT, dilutions starting at 1:10 were used; see Methods.
\*\* The random forest and the support vector machine combine all three primary tests, the accuracy measures thus do not relate to specific cut-offs.

The study was approved by the Ethics Commission of the Faculty of Medicine at LMU Munich (20-275-V) and the protocol is available online (www.koco19.de).^17^

### Laboratory Assays

All described analyses were performed using EDTA-plasma samples (appendix pp.1 for further details on assays performed, and p.7 supplemental table 3 for details on platforms and units applied).

Euroimmun Anti-SARS-CoV-2-ELISA anti-S1 IgA/IgG (hereafter called EI-S1-IgG, EI-S1-IgA; Euroimmun, Lübeck, Germany) test kits were used according to the manufacturer’s instructions. Measurement values were obtained using the quotient of the optical density measurement provided by the manufacturer’s software. We evaluated Elecsys Anti-SARS-CoV-2 Roche anti-N pan-Ig (hereafter called Ro-N-Ig; Roche, Mannheim, Germany) in accordance with the manufacturer’s guidelines. Values reported are the Cut-Off-Index (COI) of the individual samples. Operative replicates of the same samples were performed to assess reliability of primary assay performance.

For confirmatory testing, we conducted micro-virus neutralisation assays (NT) as described previously,(19) with the exception that confluent cells were incubated instead of adding cells following neutralisation reaction (appendix pp.1). We classified samples with a titre <1:10 as “NT-negative” and samples with a titre ≥1:10 as “NT-positive”. The dilution steps indicated are <10, 10, 20, 40, and >80.

SARS-CoV-2 surrogate virus neutralisation test (GS-cPass; GenScript®, Piscataway, New Jersey, USA) was used to measure binding inhibition, according to the manufacturer’s instructions. The inhibition was calculated in percentages, ranging from −30 to 100.

For SARS-CoV-2 ViraChip® microarray (VIRAMED Biotech AG, Planegg, Germany; hereafter named VC-N-IgA/IgM/IgG; VC-S1-IgA/IgM/IgG; VC-S2-IgA/IgM/IgG) execution followed the manufacturer’s instructions. We obtained measurement values by the automated ELISA-processor in arbitrary units.

As outlined by the manufacturer, we conducted the *recom*Line SARS-CoV-2 IgG line immunoassay (MG-S1, MG-N, MG-RBD; Mikrogen, Neuried, Germany), values below the cut-off of 1 were categorized “negative” without quantitative information. Common-cold CoV-2 targets of 229E, NL63, OC43, and HKU1 were included in the aforementioned assay.

### Statistical Analysis

Prior to analysis, we cleaned and locked the data. For the analyses and visualisation, we used the software R, version 4.0.2. Only one sample per individual was included in the statistical analyses; in case of individuals with multiple blood samples, we only considered the sample with the most complete dataset. For multiple measurements with complete datasets, we only included the first measurement; for operational replicates we used the latest. We subsequently carried out sensitivity and specificity analyses for true-negative and true-positive samples over all the tests performed.

We report square roots R of coefficients of determination for association among continuous variables. For paired sample comparisons, we applied Wilcoxon-sign-rank tests, whereas for multiple group comparisons we applied Kruskal-Wallis tests, followed by post-hoc Dunn tests using the Benjamini-Yekutieli adjustment for pairwise comparisons.(20)

Using true-positives and true-negatives, we determined optimised cut-off thresholds and their confidence intervals by a nonparametric bootstrap. In a similar way, we trained random forest and support vector machine classifiers. We calculated estimates for sensitivities, specificities, and overall prediction accuracies for all considered cut-off values and classifiers. This calculation was done on out-of-sample observations to avoid overfitting and thus overoptimistic performance measures. Details on the algorithms are outlined in the appendix (pp.2).

### Data & Code sharing

Data are accessible subject to data protection regulations upon reasonable request to the corresponding author. To facilitate reproducibility and reuse, the code used to perform the analyses and generate the figures was made available on GitHub (https://github.com/koco19/lab_epi) and has been uploaded to ZENODO (http://doi.org/10.5281/zenodo.4300922, DOI 10.5281/zenodo.4300922) for long-term storage.

## Results

We assessed SARS-CoV-2 antibodies in a total of 6,658 independent samples using EI-S1-IgA (n=6,657), EI-S1-IgG (n=6,658), and Ro-N-Ig (n=6,636) (figure 1, appendix p.4 supplemental table 1). Supplemental table 2 presents an overview of all tests performed (appendix p.5); table 1 outlines sensitivity and specificity of both primary and confirmatory tests by applying both manufacturers’ and optimised cut-offs.

**Figure 1:**
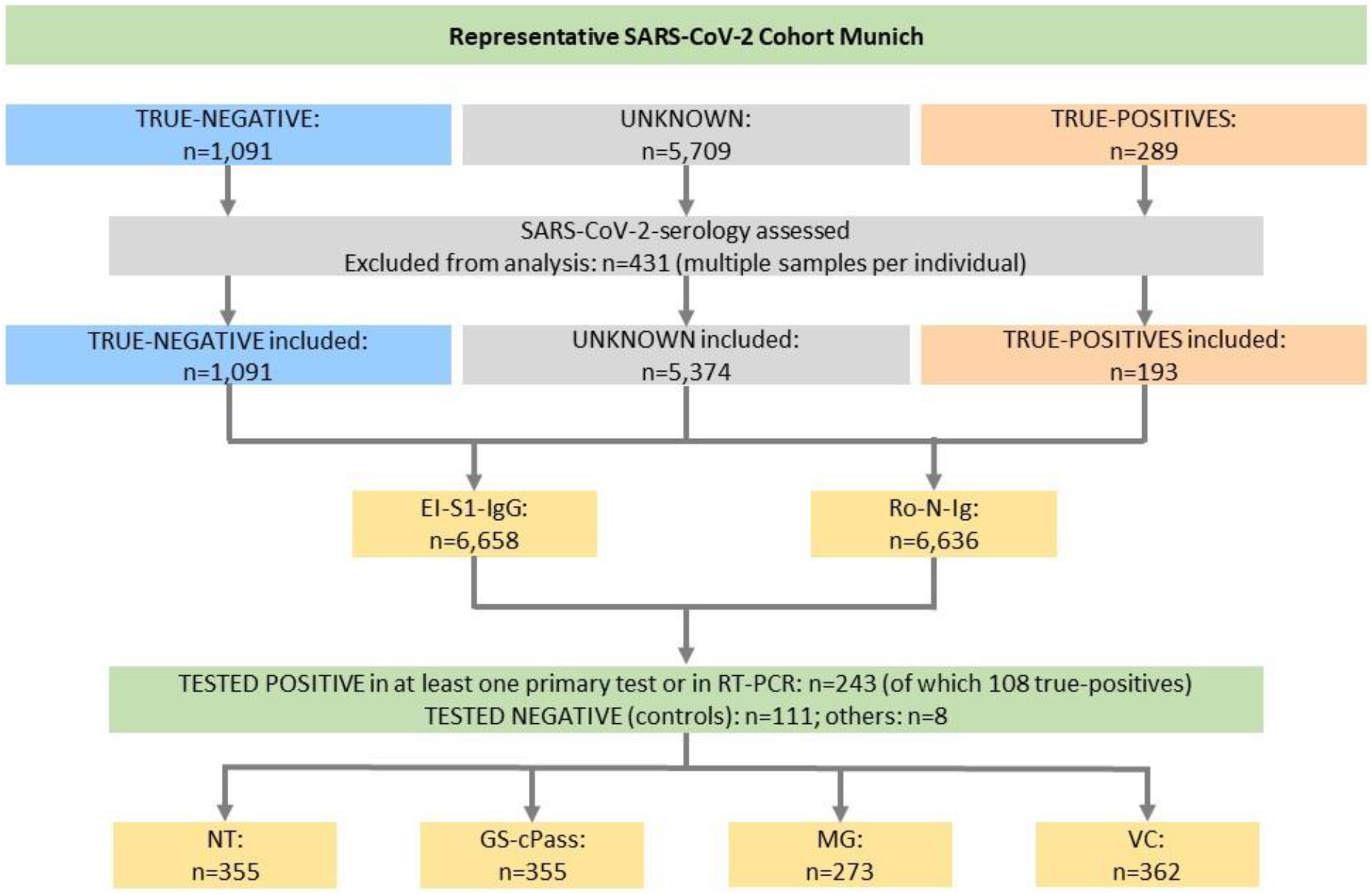
Cohort composition and characterisation of the study participants. True-positives were defined as subjects with a positive RT-PCR; true-negatives as blood donors from the pre-COVID-19 era. In addition, we included individuals recruited into the Representative COVID-19 Cohort Munich (KoCo19), 100 of which were considered as true-negatives. For each participant, a single sample was used for statistical analysis. Among individuals with longitudinal measurements, the blood sample with the most complete dataset was retained. For similar datasets, the earliest measurement was considered. For operational replicates, the latest measurement was used.

### Performance of Primary Tests

When we applied optimised cut-offs to the true-positive and true-negative samples, EI-S1-IgA had a sensitivity of 64.8% and a specificity of 92.6%, while EI-S1-IgG had a sensitivity of 79.8% and a specificity of 97.8% (table 1). The measurement value distribution of the EI-assays is depicted in figure 2A. In the second half of the study period, a decrease in overall IgA-positivity was observed, while overall IgG-positivity remained stable. Subsequent retesting of biobanked samples of the first weeks of sampling with kits of later batches demonstrated overall lower IgA-positivity rates than in the first measurements. This was likely caused by a batch effect, as retesting of samples from the second half of the sampling period did not reproduce these findings; for the first period, it led to changes in classification from positive to negative in the operational replicates (depicted as “positive-negative” in figure 2B and appendix p.8 supplemental figure 1A).

**Figure 2:**
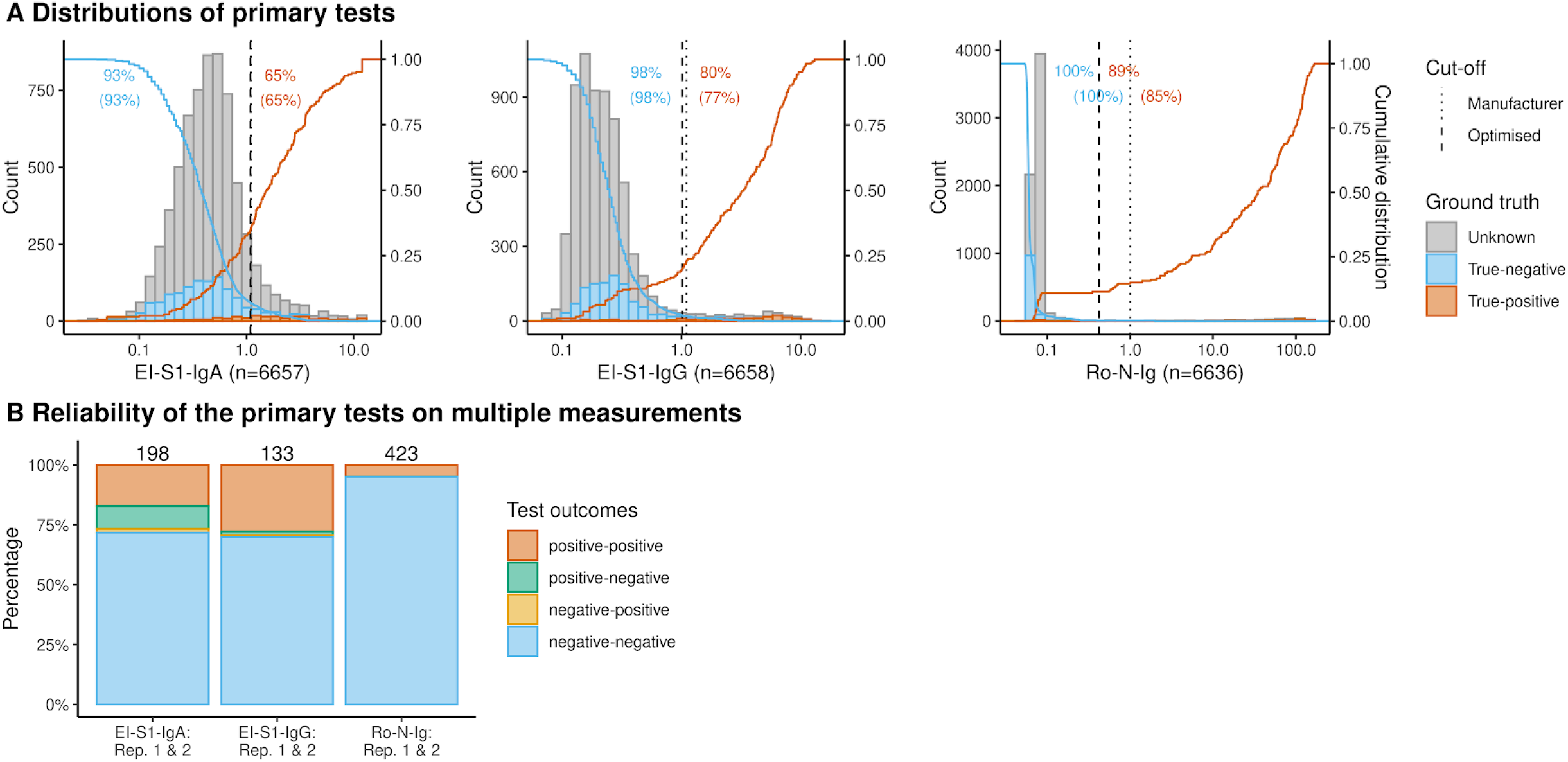
Performance of primary tests. Results of primary tests for true-negatives (*blue*), true-positives (*orange*), and individuals with unknown SARS-CoV-2 status (*grey*). Performance of EI-S1-IgA (*left*), EI-S1-IgG (*centre*), and Ro-N-Ig (*right*). Dotted lines mark the manufacturer’s cut-off value (between indeterminate and positive for EI, and between negative and positive in Ro) and dashed lines mark the optimised cut-off value as determined in this study. Orange and blue solid lines represent the percentage of test results for true-positives and true-negatives above (*blue*) or below (*orange*) the value on the x-axes, respectively. Orange and blue numbers give the percentage of true-positives and true-negatives, which were correctly detected by the test (without brackets: manufacturers’ cut-offs; within brackets: optimised cut-offs). Raw values for EI-S1-IgA show a slightly asymmetric but unimodal distribution for the overall population. The EI-S1-IgG raw values show a bimodal distribution. Ro-N-Ig raw values/COI demonstrate a narrow distribution with the bulk of values in the range COI 0.1 and below, whereas a clearly separated second population above COI 10 was observed. For EI-S1-IgG and Ro-N-Ig, the cut-offs separate the blue and orange subpopulations more reliably than for EI-S1-IgA. Consistency of the primary tests on operational replicates (manufacturer’s cut-offs). The changes from positive to negative status in EI-S1-IgA was most likely caused by a batch effect. Reliability of the other primary tests was higher.

The sensitivity and specificity of Ro-N-Ig with optimised cut-offs were 88.6% and 99.7%, respectively (table 1). Re-testing measurement values confirmed a correlation of R=1 for the quantitative measurement values without a single classification change (0/423) (figure 2B, appendix p.8 supplemental figure 1B). For all three assays, few samples tested false-negative (appendix p.26, supplemental figure 8).

For evaluation of primary test concordance, we excluded EI-S1-IgA due to inferior performance in sensitivity and specificity. The concordance between EI-S1-IgG and Ro-N-Ig was 98.5% (6,538/6,636), where 4.0% (264/6,636) of the samples were classified as positive and 94.5% (6,274/6,636) as negative unanimously. Of the remaining samples (1.5%; 98/6,636), 56.1% (55/98) were classified as positive by El-S1-IgG and negative by Ro-N-Ig (figure 4A), while the remaining 43.9% (43/98) were negative for EI-S1-IgG and positive for Ro-N-Ig. To clarify their true serostatus, confirmatory testing was performed where possible (appendix p.26 supplemental figure 8).

To assess the dependence of result read-out on sampling timepoints, we considered the temporal distribution of baseline titres over the sampling period (appendix pp.11 supplemental figure 2). Here, we considered the mean and median sample values above and below cut-off separately and found them to be comparable over the whole sampling period. Samples from blood donors were available from two distinct sample timepoints (before and after the common cold season). We noticed discrete baseline titre increases in spring without significant changes in overall positivity rate (appendix p.14 supplemental figure 3).

**Figure 3:**
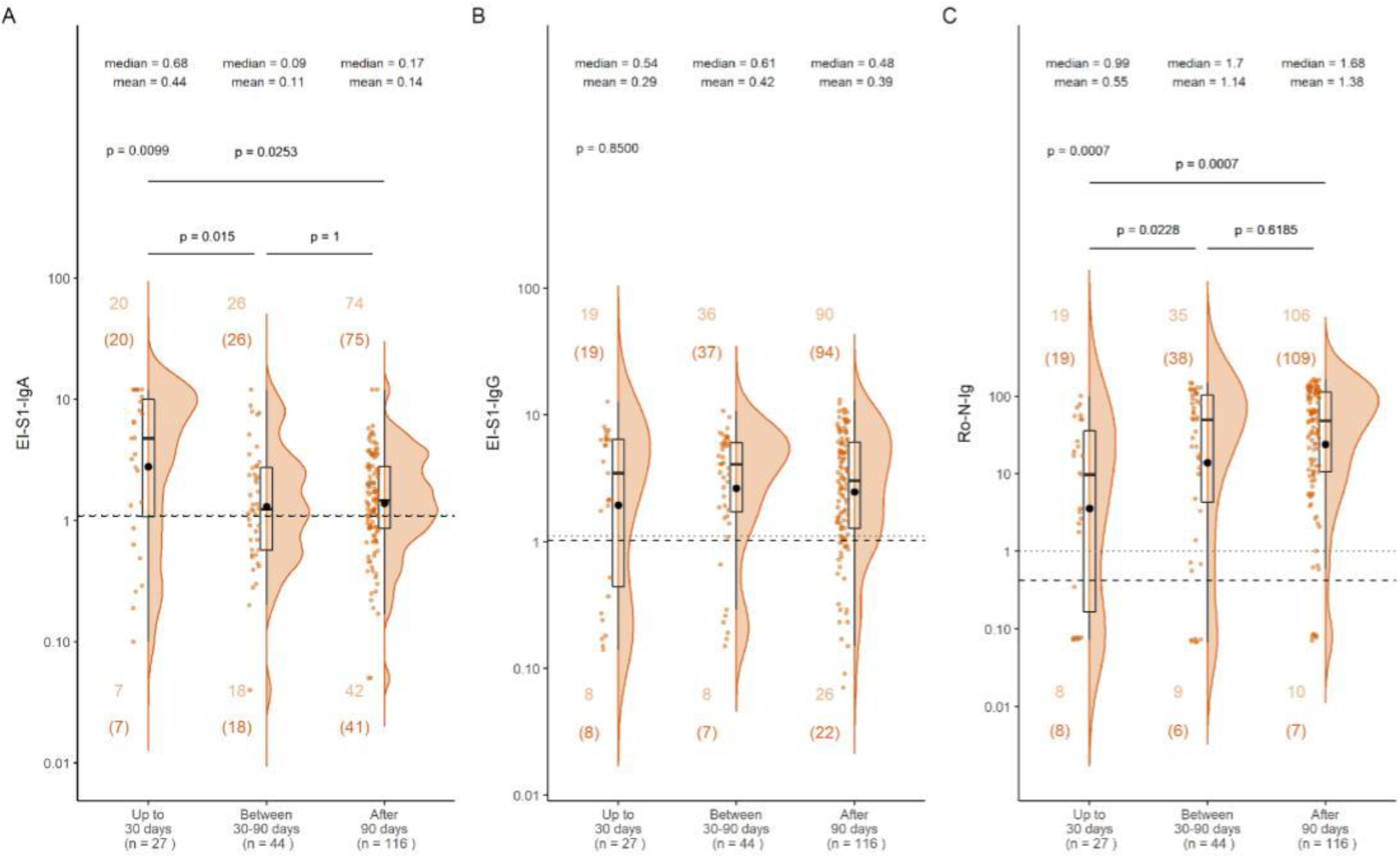
Time-dependence in primary tests for RT-PCR true-positives. Titre values of the 187 true-positives with available data on time between RT-PCR and blood sampling for (A) EI-S1-IgA, (B) EI-S1-IgG, and (C) Ro-N-Ig. The read-outs were categorized according to the time after positive RT-PCR (<30 days, 30-90 days, and >90 days). Plots show the individual read-out (orange dots), a density estimate (orange area), the 25-,50- and 75-percentiles (black box), and the mean (black dot). Counts n refer to the number of observations above/below manufacturer’s and optimised cut-off for each of the temporal groups (without brackets: manufacturers’ cut-offs; within brackets: optimised cut-offs). Pairwise differences are considered only after adjusting for multiple testing. EI-S1-IgA values were highest on average in the category of <30 days and significantly decline thereafter. EI-S1-IgG values were widespread in the group <30 days, and on average do not decline significantly >90 days. Similarly, Ro-N-Ig does not decline during the study period; however, raw values increase over time.

**Figure 4:**
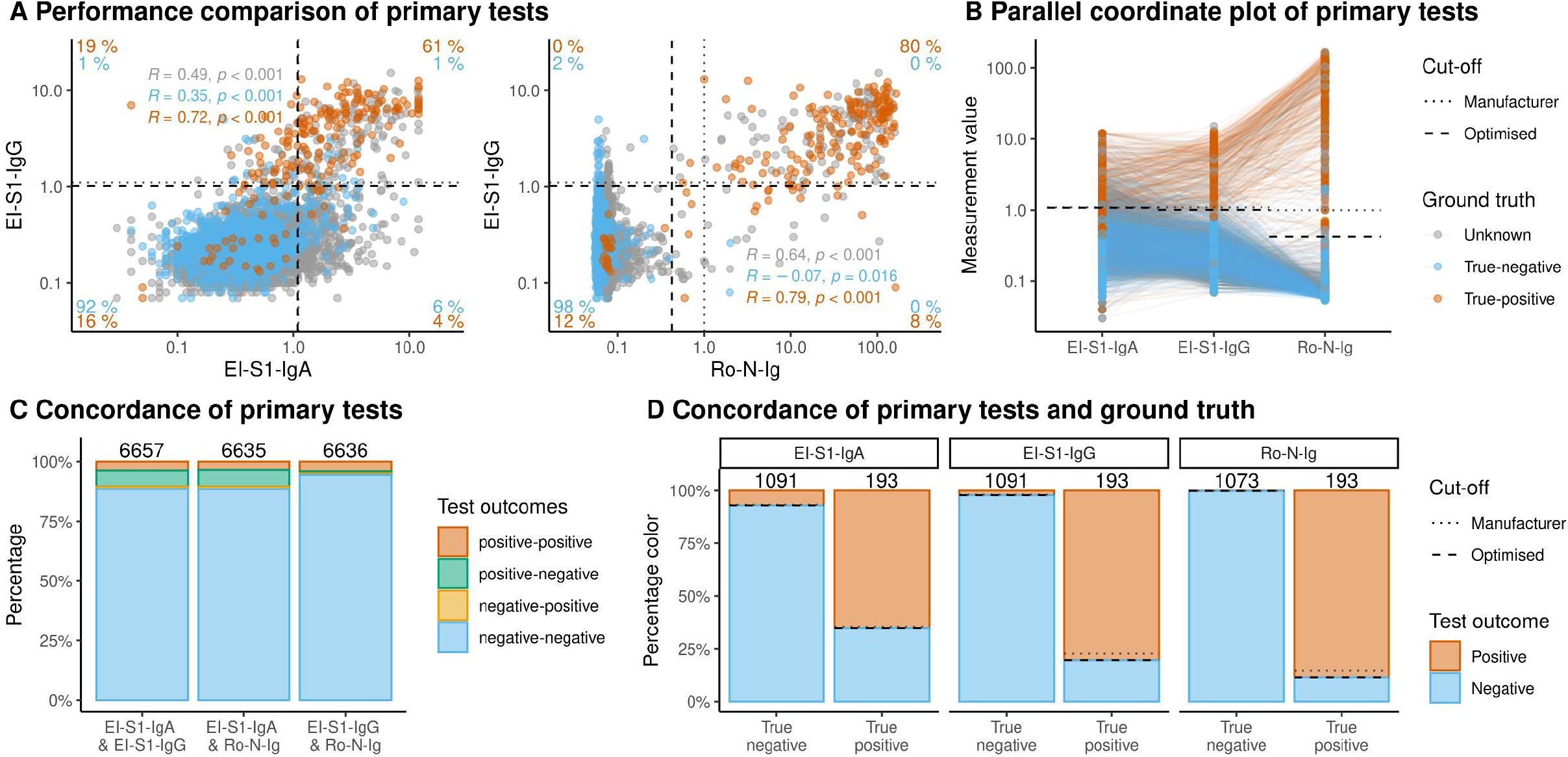
Comparison of primary tests. Results of primary tests compared to ground truth for true-negatives (*blue*), true-positives (*orange*), and individuals with unknown SARS-CoV-2 status (*grey*). The dotted lines represent the manufacturer’s cut-offs, the dashed lines the optimised cut-offs defined within this study. Pairwise scatter plots for primary tests: EI-S1-IgA vs. EI-S1-IgG (left; n=6657), and Ro-N-Ig vs. EI-S1-IgG (right; n=6636). Percentages in orange indicate fractions of true-positives in the respective quadrant with respect to all true-positives; blue for true-negatives. Percentages were calculated using the optimised cut-off. EI-S1-IgA classified 65% of true-positives correctly and 7% of the true-negatives as positive. EI-S1-IgG classified 80% of the true-positives correctly and 2% of true-negatives incorrectly. 61% of true-positives were identified by both tests unanimously. Comparing Ro-N-Ig and EI-S1-IgG, 80% of true-positives were concordantly classified as positive by both tests, while 98% of the true-negatives were classified as negative. Correlation of true-positives between Ro-N-Ig and EI-S1-IgG was R=0.79. The fraction of true-negatives falsely classified as positives in Ro-N-Ig was below 1%. Parallel coordinate plot of the primary tests. Magnitude of titres of individual samples across the different assay used were similar. The true-negative samples presented as a group in Ro-N-Ig. Only few samples display as high EI-S1-IgG and low Ro-N-Ig; on the contrary, most samples with low EI-S1-IgA titres, have higher titres in EI-S1-IgG and even higher Ro-N-Ig, presenting separately from the negative population. Concordance of primary tests (based on manufacturer’s cut-offs). Numbers of paired samples are indicated above the bars. In the two columns on the left, the unspecific reactivity of EI-S1-IgA is represented in green; the concordance of EI-S1-IgG and Ro-N-Ig was pronounced, with 98 (1.5%) discordant results (right bar). Test results of the primary tests (based on optimised cut-offs). Highest concordance was seen in Ro-N-Ig, lowest in EI-S1-IgA. The proportion of false-negatives in the true-positive cohort was 34.7%, 19.7%, and 11.4%, respectively. Applying optimised (dashed line) and manufacturer’s (dotted line) cut-off results in a reduction of false-negatives in EI-S1-IgG and Ro-N-Ig for the optimised cut-off.

To assess the dynamics of the antibody levels in RT-PCR positive subjects, we analysed the measurement values between the RT-PCR test and blood sampling for three time intervals: <30 days, 30-90 days, and >90 days. Across the whole cohort, we did not observe an initial rise of EI-S1-IgA and EI-S1-IgG but found that EI-S1-IgA decreased at later time points (p=0.02), while the distribution of EI-S1-IgG remained steady (figure 3). For Ro-N-Ig COI we detected an increase in antibody levels (p<0.001), most notably between <30 and 30-90 days (p=0.02).

### Performance of confirmatory tests

A sample subset (n=362; figure 1) underwent confirmatory testing. The overall confirmatory test performance is presented in figure 5 and table 1. The sensitivity was 73.8% for direct NT (1:10 dilution), 96.3% for GS-cPass, and 94.9% for MG-RBD. All three tests had a specificity close to 100% (figure 5).

**Figure 5:**
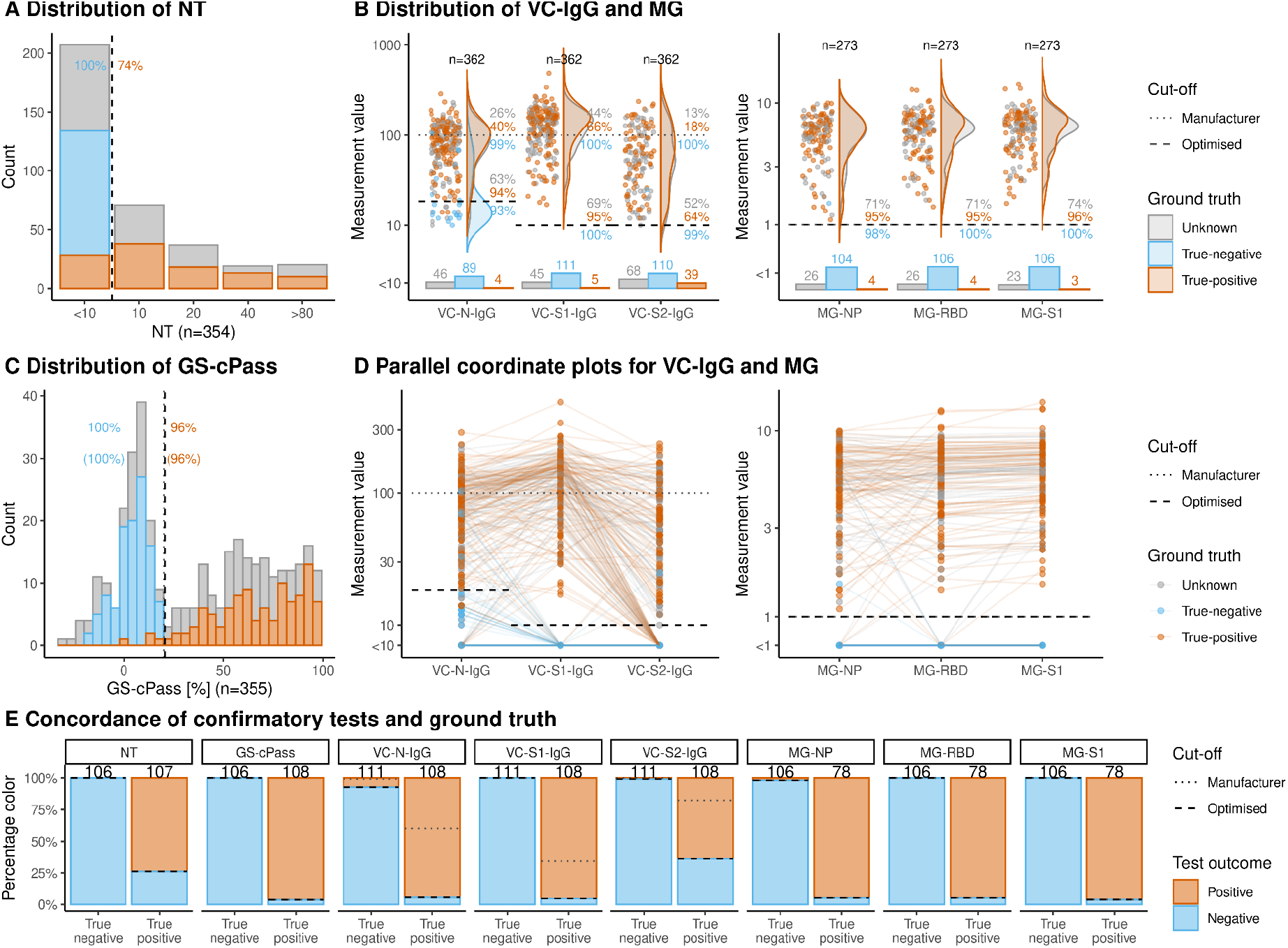
Confirmatory tests. Results of confirmatory tests compared to ground truth for true-negatives (*blue*), true-positives (*orange*), and individuals with unknown SARS-CoV-2 status (*grey*). Black dotted and dashed lines represent the manufacturer’s and the optimised cut-offs, respectively. Orange/blue numbers indicate percentages of true-positives/-negatives correctly detected by the test. (A,C) Distribution of results of NT and GS-cPass. At 1:10 dilution, NT endpoint was categorical with a specificity of 100% in this cohort; sensitivity was 73.8%. GC-cPass manufacturer’s cut-off yielded a specificity of 99.1% and a sensitivity of 96%. Adjustments of the cut-off did not improve the performance of the test systems (shown in parentheses). (B) Distribution of results of the VC-array (left) and the MG-line blot (right). Bar charts below violin plots represent information for the categorical part of the tests. Grey numbers give the percentages of positive samples with unknown SARS-CoV-2 as determined by the manufacturer’s and optimised cut-offs. Percentages were calculated over the total number of samples of unknown SARS-CoV-2 status with available test results. In the VC-array, S1-IgG / N-IgG performed best as confirmatory tests with 95.4% / 93.5% sensitivity, and 100% / 91.9% specificity, respectively, applying optimised cut-offs. With manufacturer’s cut-offs, sensitivity was 65.7% / 39.8%, respectively. With the optimised cut-off, VC-S2-IgG had a sensitivity of 63.9% and a specificity of 99.1%. Performance of VC-S2-IgA and VC-S2-IgM are presented for reference in supplemental figure 4 (appendix p.17), demonstrating a potential use in diagnostics settings. MG-N, −S1 and -RBD had sensitivities of 94.9% or higher, and specificities of 98.1% or higher, no optimisation of cut-offs could not be optimised here. (D) Parallel coordinate plot of VC-Array and MG. Many subjects do not show S2 reaction although strong reactivity against N and S1 can be detected. Comparing individual subjects in NP, RBD and S1 reaction indicates that several individuals developed strong N and S1 reactions whereas RBD was not bound. General concordance between S1 and RBD was, however, observed. (E) Description of concordance between results comparing each confirmatory test with the ground truth. The colour coding is based on the optimised cut-off.

NT-titres in our cohort were low – mostly 1:10 – and few had high neutralisation titres of 1:80 or above (figure 5A). Measurement values for MG-S1 and MG-RBD were often similar, with few samples showing a reaction against S1 (and N) but not RBD (3.7%; 5/134). For the VC-array, sensitivities of both VC-S1-IgG and VC-N-IgG were improved markedly by optimising cut-offs, with gains of >30% (VC-N-IgG 39.8%/93.5%; VC-S1-IgG 65.8/93.4%; table 1, figure 5). Notably, a considerable number of samples (36.0%; 40/111) were reactive against S1 and N but not S2 (figure 5D).

Figure 6 and supplemental figures 4,5, and 10 (appendix pp.16) compare confirmatory tests. For surrogates of viral neutralisation, 3.2% (3/95) were positive in NT and GS-cPass but not in MG-RBD (figure 6D).

**Figure 6:**
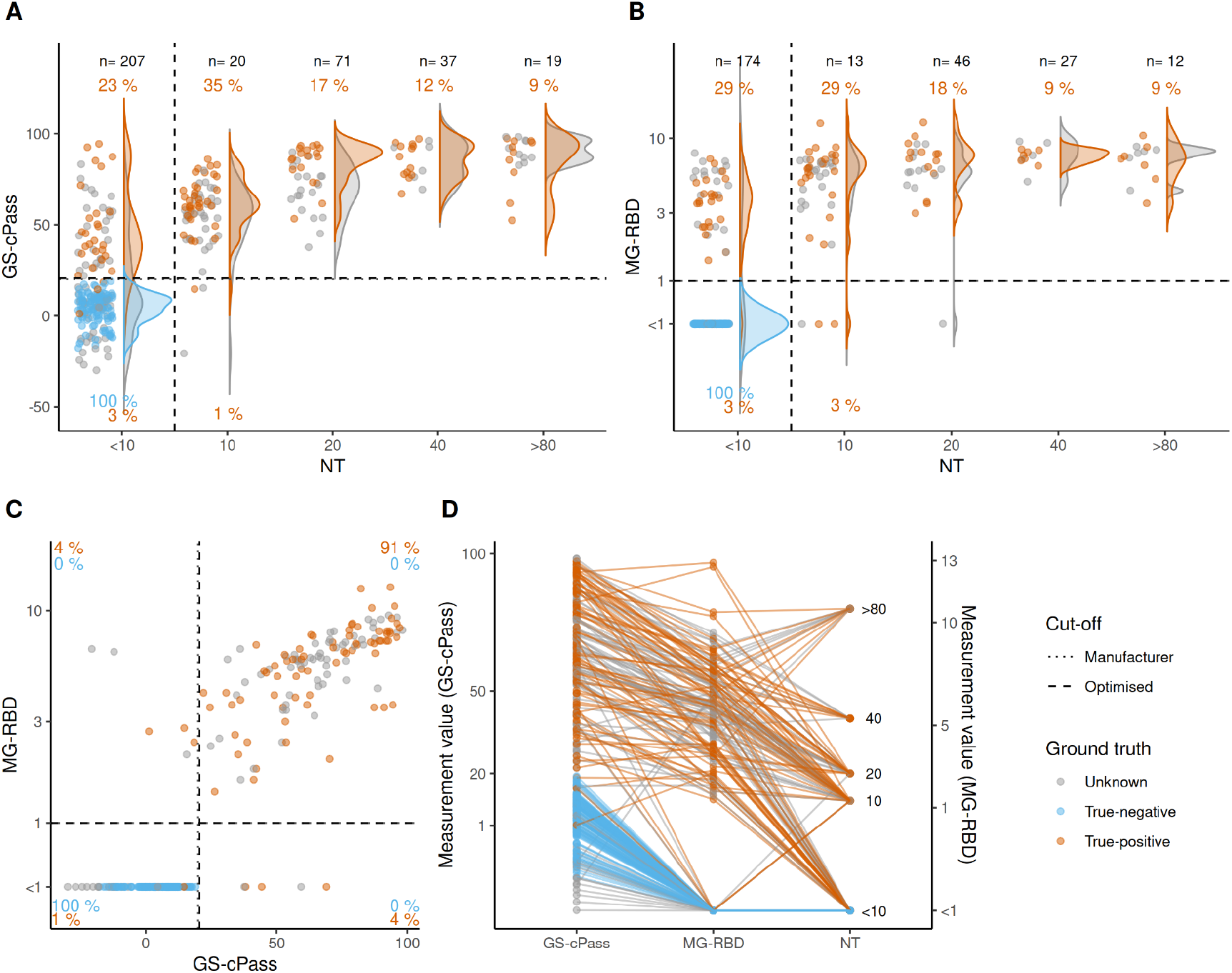
Comparison of confirmatory tests. Comparison of confirmatory tests for true-negatives (*blue*), true-positives (*orange*), and individuals with unknown SARS-CoV-2 status (*grey*). (A) Association between the categorical endpoint of NT and the continuous results of GS-cPass. The test results were positively related; in cases of identical classification agreement with ground truth was frequent. However, more than 20% of true-positives were missed by NT (1:10 dilution; n=354). (B) Association between the categorical endpoint of NT and the continuous results of MG-RBD. In true-positives with low direct neutralization capacity (NT <10), distinction between negative and positive populations was observed with GS-cPass (n=272), highlighting the limited sensitivity of NT (1:10 dilution). (C) Association between GS-cPass and MG-RBD presented with discordant results in 8% (of true-positives) at intermediate test read-outs. The distribution in higher titre ranges were narrow (n=272). (D) Parallel coordinate plot of the two neutralization surrogate tests and NT. Samples with high/low raw values demonstrated similarly in all three tests, as lines present horizontally. However, a subset of individuals with relatively high GS-cPass values and positive outcome in NT were below the cut-off in the MG-RBD assay.

### Associations of confirmatory and primary tests

To examine pretest probability of assays following positive initial testing, the measurement values of all primary and confirmatory tests were correlated (figure 7, appendix pp.19 supplementary figures 6, 7, 9). Overall, we observed good correlations, particularly for GS-cPass and MG-RBD with EI-S1-IgG (figure 7B, C), and MG-N with Ro-N-Ig (figure 7H).

**Figure 7:**
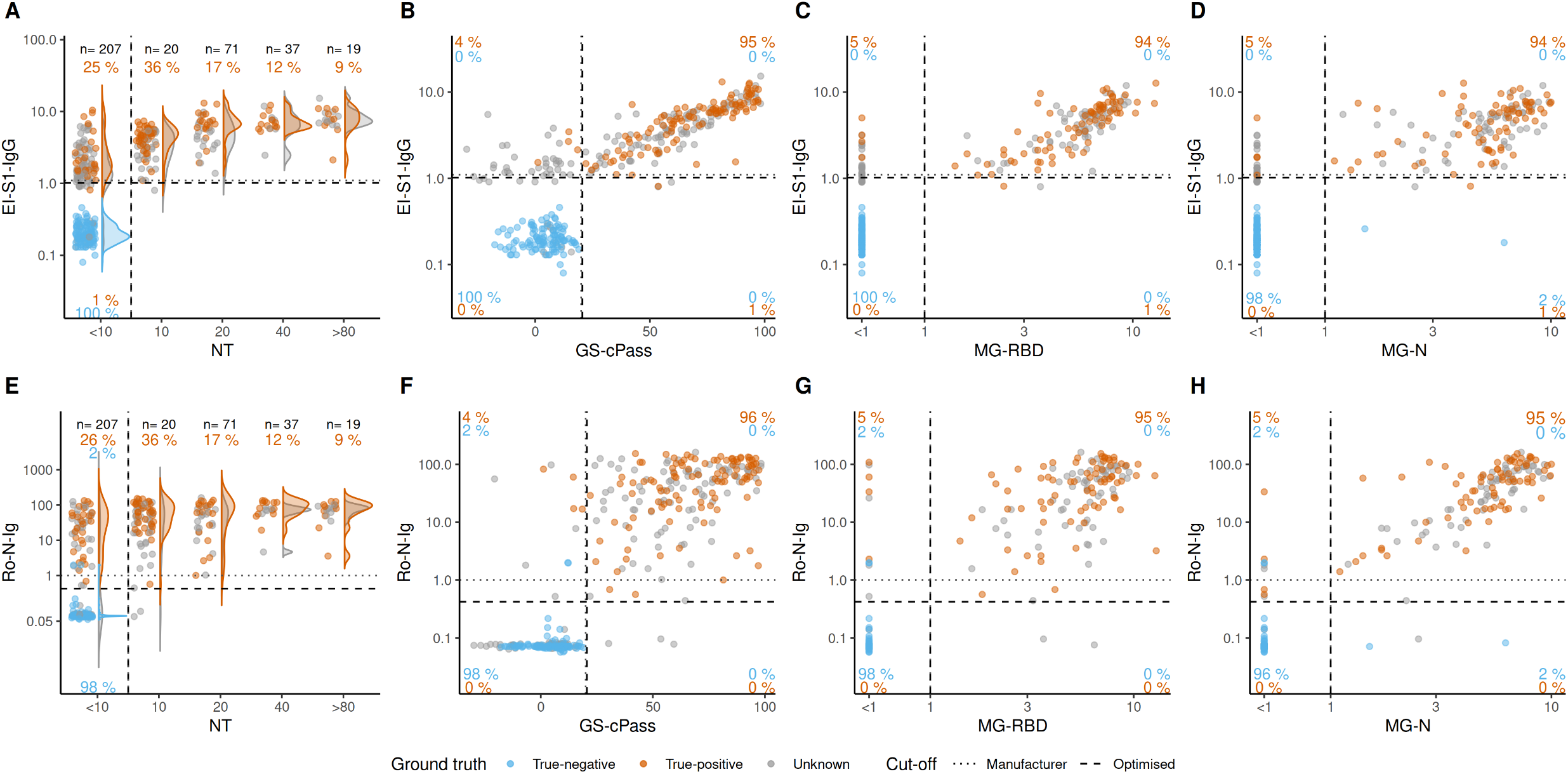
Comparison of primary tests (EI-S1-IgG, Ro-N-Ig) with confirmatory tests (NT, GS-cPass MG-RBD, MG-N). Comparison of primary with confirmatory tests for true-negatives (*blue*), true-positives (*orange*), and individuals with unknown SARS-CoV-2 status (*grey*). (A) EI-S1-IgG and NT show positive correlation (n=354), although 1% of the true-positives were missed by NT (1:10 dilution). (B, C) Correlation of EI-S1-IgG with GS-cPass (n=361) and MG-RBS (n=272) presented as dose dependent for the double-positive values. Concordance for the true-positive subset was >94%. (D) Association of EI-S1-IgG to MG-N (n=355) presented dose-dependent, but not as markedly. (E-H) Association between Ro-N-Ig and the confirmatory tests (n=362, n=273, n= 354, n=354) did not present as dose-dependent, compared to EI-S1-IgG. Concordance between the tests was >95%; except NT, which presented similar patterns to EI-S1-IgG. We observed a population in the upper left quadrant, (B, C, D), clearly negative in the confirmatory tests.

The categorical concordance for GS-cPass, MG-RBD, and MG-N with both Ro-N-Ig and EI-S1-IgG was similar (94% or above), while the concordance of NT with both primary tests was lower (74%; figure 7A-H). Concordances were improved by applying the optimised cut-offs, especially for VC-S1-IgG and VC-S2-IgG.

To derive an algorithm for an optimised testing-strategy, we also investigated potential gains when applying multiple primary tests. Interestingly, combining different primary tests via machine learning techniques (random forest and support vector machine) barely improved the performance beyond what was achieved by Ro-N-Ig alone (table 1), neither could we demonstrate an added value of performing more than one confirmatory test.

### Common cold line blot

Cross-reactivity between common cold CoV anti-N and SARS-CoV-2 anti-N was assessed by line blot against 229E, NL63, OC43, and HKU1 for various samples subjected to confirmatory testing (n=273/362; appendix p.27, supplemental figure 9). The distribution for 229E and NL63 showed a significant association with SARS-CoV-2 infection and positivity (p<0.001 for both).

## Discussion

We performed head-to-head comparison of seven seroassays for SARS-CoV-2 in a well-defined cohort with 6,658 samples. The overwhelming majority of infections could be detected by antibodies even more than three months after infection. In addition, we generated optimised cut-offs which improved sensitivity in primary tests and enhanced the performance of several confirmatory tests. We furthermore showed that surrogate tests, such as GeneScript®cPass™ or RBD line-blot, should be considered instead of the infrastructurally challenging NT in large scale studies, such as vaccine trials or epidemiological surveys. In addition, we observed limited seasonality effects and a significant cross-reactivity with common-cold CoV 229E and NL63 in SARS-CoV-2 true-positive subjects.

We calculated diagnostic accuracy indicators for seroassays based on true-positive and true-negative samples and derived optimised cut-offs for the evaluated assays. We subsequently applied these to a set of samples of unknown infection status.(17) As reported previously, there is little diagnostic gain from EI-S1-IgA if EI-S1-IgG is used on the same samples.(4) Our study suggests that Ro-N-Ig performs more reliably than EI-S1-IgA and EI-S1-IgG; hence the application of the latter two is of questionable use for low-prevalence settings, considering their lower specificity.

For seroprevalence studies it is essential to know how long the measured antibody response remains stable and detectable. There have been different reports of a rapid decline in titre over time.(10, 21) In this analysis, we observed similar declines for EI-S1-IgA titres, which were highest within the first 30 days but dropped significantly thereafter. In contrast, for both EI-S1-IgG and Ro-N-Ig, average titres did not decrease over a period of more than 90 days.

We assessed confirmatory test performances by a subset of true-negative samples. The assays targeting the highly-specific receptor binding domain (NT, GS-cPass, MG-RBD) are considered direct or surrogate markers for viral neutralisation.(22) The cell-culture free tests performed particularly well with sensitivities of 96.3% for GS-cPass and 94.9% for MG-RBD, using the manufacturer’s thresholds. In contrast, we improved the sensitivity for the VC-array markedly by applying optimised cut-offs, with a gain of >50% for VC-N-IgG (39.8%-93.5%) and almost 30% for VC-S1-IgG (65.8-93.4%).

To generate reliable data for surrogate neutralisation markers (GS-cPass, MG-RBD) feasible for epidemiological purposes, true-positive samples with low neutralising activity are preferable. Our cohort consists mainly of oligo- or asymptomatic COVID-19 subjects – NT-titres observed were generally low as expected – suggesting an NT dilution decrease from 1:10 to 1:5 might improve sensitivity in similar settings. As MG-RBD, GS-cPass, and NT performed similarly and correlated well, suggesting an interchangeability of one another, for example for certain epidemiological questions. NT requires complex BSL-3-laboratory infrastructures (virus-culture) which currently represents a critical bottleneck, while GC-cPass and MG-RBD can be performed under much less stringent safety requirements. While NT is a direct representation of viral neutralisation, GS-cPass assesses the antibody-mediated inhibition of ACE2-interaction with SARS-CoV-2-S1-RBD and is therefore a cell-free neutralisation surrogate marker.(23) Both assays are independent of antibody-subclasses. In contrast, MG-RBD solely detects the RBD-IgG-antibody interaction. The discrepant results in our sample set (figure 6D) suggest either a different neutralisation mode than binding to the RBD as used in MG-RBD, or neutralisation due to other subclasses than IgG.

The influence of previous or acute infections with common-cold CoV and a resulting cross-reactivity in SARS-CoV-2 assays has been discussed previously.(15, 16) Biologically, it is impacted by the target used, wherein whole virus- or nucleocapsid proteins are particularly prone to cross-reactivity; this also applies for SARS-CoV-2.(6, 14, 15, 24) Comparing true-negative samples from before and after the common cold season, we observed a marginal seasonal increase in the measurement values of EI-S1-IgA, EI-S1-IgG, and Ro-N-Ig (supplemental figure 3). There was a clear association between SARS-CoV-2 RT-PCR positivity and reactivity against 229E and NL63 N-protein; this suggests that SARS-CoV-2 infected individuals develop a cross-reactivity in the assay against two of the four tested CoV strains. Alternatively, in subjects with previous common cold, this could be explained by a cross-stimulation of pre-existing cells specific to the respective common cold coronavirus strain. Similar findings were described recently in a systematic review.(6)

Using the best-performing primary tests (EI-S1-IgG and Ro-N-Ig), we observed a relevant number of discordantly classified samples (1.5%; 98/6,636; appendix p. 27 supplemental figure 8). Constellations of Ro-N-Ig negative and EI-S1-IgG positive were observed frequently (56.1%; 55/98), with many samples being true-negatives (41.8%; 23/55), but none being true-positive (0%; 0/55). The overwhelming majority of those with unknown infection status were negative in NT and GS-cPass (87.5%; 28/32). As the testing principle of EI-S1-IgG is based on S1-reactivity, positive virus neutralisation or a neutralisation surrogate can be expected if the reaction is specific, suggesting unspecific cross-reactivity in this group.

In contrast, discordant serology with Ro-N-Ig positive and EI-S1-IgG negative was less frequent (43.9%; 43/98), mostly from true-positives >40 days after positive RT-PCR (16.3%; 16/98). This is strongly suggestive of an inability of these individuals to mount a detectable response against S1 during the natural infection. Late or lacking seroconversion has been described previously, mostly in oligo- or asymptomatic subjects.(25, 26) Reports speculated about vastly varying proportions of subjects unable to mount an antibody response detectable by commonly used assays.(21, 27, 28) In a systematic review by Huang et al., the median detection time across different antibodies against SARS-CoV-2 was 11 days, similar to SARS-CoV-1;(6) however, serological kinetics vary across the severity gradients of symptoms.(6, 9-11) Additionally, misclassification of subjects might have occurred due to RT-PCR-sample mix-up, unspecific reactivity, or contamination in the PCR-test; plausible scenarios in the current pandemic situation where clinical and laboratory infrastructures are overburdened with high sample volumes. In oligo- or asymptomatic subjects, a positive RT-PCR is often the only evidence of infection with SARS-CoV-2 and therefore a misclassification might not be detected.

In our cohort of mostly oligosymptomatic true-positive subjects, 11.4% (22/193) were not detected in the primary serological tests. Of these, 36.4% (8/22) were below 30 days after RT-PCR. A total of 6.0% (7/116) showed no evidence of seroconversion in the group >90 days after RT-PCR. For one subject, the RT-PCR date was unknown. Overall, in our dataset of samples >30 days after positive RT-PCR, a modest 8.1% (13/160) remained negative.

In addition to identifying suitable tests dependent upon the approach chosen, applying optimised cut-offs might be a tool to enhance test performance.(4, 29) For EI-S1-IgG, Meyer et al. proposed optimised thresholds for both lower and higher cut-offs.(4) However, a seroprevalence study in Geneva compared both recommended and optimised cut-offs, but did not see any qualitative change.(30) The authors proposed evaluating the manufacturer’s cut-off before routine testing, and highlighted the dilemma of securing both rule-in and rule-out properties to mitigate the risk of incorrect classification in a situation with highly-dynamic pre-test probabilities.(4) For both EI-S1-IgG and Ro-N-Ig we demonstrated that sensitivity improved using optimised cut-offs, while specificity remained similar. Even though these changes seem minimal, they might translate into a high number of incorrect diagnoses when testing is performed on a large scale: this is especially pertinent in low-prevalence settings, as a particularly high specificity is crucial to achieve a high positive predictive value. It may also be preferable to have a more sensitive cut-off for a primary test and confirm the positives with a highly-specific secondary test system in low prevalence settings.(17)

Our study has several of limitations. Firstly, the samples are mostly derived from subjects with mild to no symptoms and therefore conclusions drawn here might best apply to an epidemiological rather than a diagnostic approach. Further evaluation using specimens from severely ill subjects or from high-prevalence settings will need to be conducted and compared to the enclosed results.

Secondly, the dataset used to estimate optimised cut-offs and performance of the tests is not an unbiased random sample, as all were sampled in the city of Munich, Germany. It cannot be ruled out that blood donors are generally healthier than the general population. Furthermore, not all confirmatory tests were performed on all samples; only a subset, namely those with positive results in at least one primary test and a dedicated negative/positive cohort, were tested using these systems. Finally, we did not have information on underlying health conditions known to increase the quantity of polyclonal antibodies.

In conclusion, our study provides a head-to-head cross-comparison of tests and can be used as a resource to enable the refinement of testing-strategies for individual and public-health use. We also investigated the correlations of the different test systems in-depth, considering confounders such as seasonality and titres against common-cold CoV strains. By combining a diagnostic accuracy approach in a well-defined sample set with true-positive as well as true-negative specimens and extrapolating the established findings to samples derived from a population-based seroprevalence cohort, we were able to identify reliable test systems for our Prospective COVID-19 Cohort Munich (KoCo19).(17)

## Supporting information

Appendic and Supplemental Material

## Acknowledgements

We wholeheartedly thank all study participants for their trust, time, data, and samples. We are grateful for the financial support of the Bavarian State Ministry of Science and the Arts, the University Hospital of LMU Munich, the Helmholtz Centre Munich, the University of Bonn, the University of Bielefeld, and the German Ministry for Education and Research. This study would also not have been possible without the passionate contribution of the staff of the Division of Infectious Diseases and Tropical Medicine at the University Hospital, LMU Munich, Helmholtz Centre Munich, Bundeswehr Institute of Microbiology, as well as all medical students involved. We thank Judith Eckstein for outstanding support regarding public relations. We thank the teams from the press offices of LMU, University Hospital of LMU Munich, and of Helmholtz Centre Munich. We thank the KoCo19 advisory board members Stefan Endres, Stephanie Jacobs, Bernhard Liebl, Michael Mihatsch, Matthias Tschöp, Manfred Wildner, and Andreas Zapf. We thank Accenture for the development of the KoCo19 web-based survey application. We are grateful to the Statistical Office of the City of Munich, Germany, for providing statistical data on the Munich general population. We thank Helmut Küchenhoff for a critical review of an earlier version of the manuscript and Jared Anderson for English language corrections. We are grateful to the Munich police for their support in the fieldwork and for the Munich Surgical Imaging GmbH, Cisco Systems, and the graphic/photo/IT infrastructure departments at the University Hospital of LMU Munich provided support during video production and online events. For their assistance with our fieldwork, we thank the BMW Group for providing free cars as a part of their campaign “BMW hilft Helfenden.” We would also like to thank Mercedes-Benz Munich who supported the project infrastructure with Mercedes-Benz Rent. Lastly, we thank the IT Infrastructure Department of the LMU University Hospital Munich. MG acknowledges the support from the Joachim Herz Foundation through the Add-on Fellowship for Interdisciplinary Science.

## References

1. Huang C, Wang Y, Li X, Ren L, Zhao J, Hu Y, et al. Clinical features of patients infected with 2019 novel coronavirus in Wuhan, China. The Lancet. 2020;395(10223):497–506.

2. Amanat F, Stadlbauer D, Strohmeier S, Nguyen THO, Chromikova V, McMahon M, et al. A serological assay to detect SARS-CoV-2 seroconversion in humans. Nat Med. 2020;26(7):1033–6.

3. Chen X, Pan Z, Yue S, Yu F, Zhang J, Yang Y, et al. Disease severity dictates SARS-CoV-2-specific neutralizing antibody responses in COVID-19. Signal Transduction and Targeted Therapy. 2020;5(1):180.

4. Meyer B, Torriani G, Yerly S, Mazza L, Calame A, Arm-Vernez I, et al. Validation of a commercially available SARS-CoV-2 serological immunoassay. Clinical Microbiology and Infection. 2020;26(10):1386– 94.

5. (FIND) FfIND. SARS-CoV-2 diagnostic pipeline 2020 [Available from: https://www.finddx.org/covid-19/pipeline/.

6. Huang AT, Garcia-Carreras B, Hitchings MDT, Yang B, Katzelnick LC, Rattigan SM, et al. A systematic review of antibody mediated immunity to coronaviruses: kinetics, correlates of protection, and association with severity. Nature Communications. 2020;11(1):4704.

7. Cheng MP, Yansouni CP, Basta NE, Desjardins M, Kanjilal S, Paquette K, et al. Serodiagnostics for Severe Acute Respiratory Syndrome-Related Coronavirus 2 : A Narrative Review. Ann Intern Med. 2020;173(6):450–60.

8. Nuccetelli M, Pieri M, Grelli S, Ciotti M, Miano R, Andreoni M, et al. SARS-CoV-2 infection serology: a useful tool to overcome lockdown?Cell Death Discovery. 2020;6(1):38.

9. Long Q-X, Tang X-J, Shi Q-L, Li Q, Deng H-J, Yuan J, et al. Clinical and immunological assessment of asymptomatic SARS-CoV-2 infections. Nature medicine. 2020:1–5.

10. Seow J, Graham C, Merrick B, Acors S, Steel KJA, Hemmings O, et al. Longitudinal evaluation and decline of antibody responses in SARS-CoV-2 infection. medRxiv. 2020:2020.07.09.20148429.

11. Eyre DW, Lumley SF, O’Donnell D, Stoesser NE, Matthews PC, Howarth A, et al. Stringent thresholds for SARS-CoV-2 IgG assays result in under-detection of cases reporting loss of taste/smell. medRxiv. 2020:2020.07.21.20159038.

12. Zhao J, Yuan Q, Wang H, Liu W, Liao X, Su Y, et al. Antibody responses to SARS-CoV-2 in patients of novel coronavirus disease 2019. Clin Infect Dis. 2020.

13. Ainsworth M, Andersson M, Auckland K, Baillie JK, Barnes E, Beer S, et al. Performance characteristics of five immunoassays for SARS-CoV-2: a head-to-head benchmark comparison. The Lancet Infectious Diseases.

14. Meyer B, Drosten C, Müller MA. Serological assays for emerging coronaviruses: challenges and pitfalls. Virus research. 2014;194:175–83.

15. Grifoni A, Weiskopf D, Ramirez SI, Mateus J, Dan JM, Moderbacher CR, et al. Targets of T Cell Responses to SARS-CoV-2 Coronavirus in Humans with COVID-19 Disease and Unexposed Individuals. Cell. 2020;181(7):1489-501.e15.

16. Klompus S, Leviatan S, Vogl T, Kalka I, Godneva A, Shinar E, et al. Cross-reactive antibody responses against SARS-CoV-2 and seasonal common cold coronaviruses. medRxiv. 2020:2020.09.01.20182220.

17. Pritsch M, et al. Prevalence and risk factors of infection in the representative COVID-19 cohort Munich. Adjacent manuscript.

18. Radon K, Saathoff E, Pritsch M, Guggenbühl Noller JM, Kroidl I, Olbrich L, et al. Protocol of a population-based prospective COVID-19 cohort study Munich, Germany (KoCo19). BMC Public Health. 2020;20(1):1036.

19. Haselmann V, Özçürümez MK, Klawonn F, Ast V, Gerhards C, Eichner R, et al. Results of the first pilot external quality assessment (EQA) scheme for anti-SARS-CoV2-antibody testing. Clinical chemistry and laboratory medicine. 2020.

20. Benjamini Y, Yekutieli D. The control of the false discovery rate in multiple testing under dependency. Annals of statistics. 2001:1165-88.

21. Long QX, Liu BZ, Deng HJ, Wu GC, Deng K, Chen YK, et al. Antibody responses to SARS-CoV-2 in patients with COVID-19. Nat Med. 2020;26(6):845–8.

22. Chi X, Liu X, Wang C, Zhang X, Li X, Hou J, et al. Humanized single domain antibodies neutralize SARS-CoV-2 by targeting the spike receptor binding domain. Nature Communications. 2020;11(1):4528.

23. Tan CW, Chia WN, Qin X, Liu P, Chen MIC, Tiu C, et al. A SARS-CoV-2 surrogate virus neutralization test based on antibody-mediated blockage of ACE2–spike protein–protein interaction. Nature Biotechnology. 2020;38(9):1073–8.

24. Braun J, Loyal L, Frentsch M, Wendisch D, Georg P, Kurth F, et al. SARS-CoV-2-reactive T cells in healthy donors and patients with COVID-19. Nature. 2020.

25. Staines HM, Kirwan DE, Clark DJ, Adams ER, Augustin Y, Byrne RL, et al. Dynamics of IgG seroconversion and pathophysiology of COVID-19 infections. medRxiv. 2020:2020.06.07.20124636.

26. Tuaillon E, Bolloré K, Pisoni A, Debiesse S, Renault C, Marie S, et al. Detection of SARS-CoV-2 antibodies using commercial assays and seroconversion patterns in hospitalized patients. The Journal of infection. 2020;81(2):e39–e45.

27. Gallais F, Velay A, Wendling M-J, Nazon C, Partisani M, Sibilia J, et al. Intrafamilial Exposure to SARS-CoV-2 Induces Cellular Immune Response without Seroconversion. medRxiv. 2020:2020.06.21.20132449.

28. Okba NM, Müller MA, Li W, Wang C, GeurtsvanKessel CH, Corman VM, et al. Severe acute respiratory syndrome coronavirus 2-specific antibody responses in coronavirus disease patients. Emerging infectious diseases. 2020;26(7):1478–88.

29. Plebani M, Padoan A, Negrini D, Carpinteri B, Sciacovelli L. Diagnostic performances and thresholds: the key to harmonization in serological SARS-CoV-2 assays? medRxiv. 2020:2020.05.22.20106328.

30. Stringhini S, Wisniak A, Piumatti G, Azman AS, Lauer SA, Baysson H, et al. Seroprevalence of anti-SARS-CoV-2 IgG antibodies in Geneva, Switzerland (SEROCoV-POP): a population-based study. Lancet. 2020.

