## Appendic and Supplemental Material for "A Serology Strategy for Epidemiological Studies Based on the Comparison of the Performance of Seven Different Test Systems - The Representative COVID-19 Cohort Munich"

### Appendix and Supplementary Material:

#### Cohorts

Samples for testing were derived from three different sample sets (supplemental table 1, figure 1). For known negatives (true-negatives), specimens from 991 healthy blood donors, sampled at two distinguished time points were used (i. October 2019, to represent the pre-common cold period and ii. March 2020, pre-COVID19 and post-common cold season). For known positives (true-positives), volunteers ≥18 years that tested RT-PCR-positive for SARS-CoV-2 in Munich were sampled (n=193). Furthermore, specimens from the Prospective COVID-19 Cohort Munich (KoCo19) were included in the analyses (n=5,474; of which an additional subset of 100 true-negatives was identified). In total, 6,658 samples were included in the analyses for the primary tests (Euroimmun Anti-S1-SARS-CoV-2-ELISA-IgA (EI-S1-IgA; n=6,657), Euroimmun Anti-SARS-CoV-2-ELISA-IgG (EI-S1-IgG; n=6,658), and Elecsys Anti-SARS-CoV-2 Roche N pan−Ig (Ro-N-Ig; n=6,636)) (supplemental table 2). A large part of samples that tested positive in primary tests, in addition to a subset of true-negatives, were subjected to confirmatory testing (n=362; figure 1, supplemental table 2).

#### Assay Performances

**Euroimmun anti-S1 IgA/IgG (Euroimmun Anti-SARS-CoV-2-ELISA):**
EI-S1-IgG and EI-S1-IgA test kits were utilised according to the manufacturer's instructions using an Euroanalyzer-1 robot (Euroimmun, Lübeck, Germany). Raw values presented in all figures of the manuscript are the quotients of the optical density measurements given by the manufacturer’s software.

**Roche anti-N pan-Ig (Elecsys Anti-SARS-CoV-2):**
Ro-N-Ig (Roche, Mannheim, Germany) testing was conducted in accordance with the manufacturer’s guidelines. Measurements were performed on the cobas 400-e411 and/or 8000-e801 modules (Roche, Basel, Switzerland) there were no differing classifications between two employed instruments (Figure 2B, Supplemental Figure 1B) and the performance of both instruments was comparable. Values indicated in all tests correspond to the Cut-Off-Index (COI) of the individual samples.

**GenScript® cPass:**
SARS-CoV-2 surrogate virus neutralisation test (GenScript®, Piscataway, New Jersey, USA; hereafter called ”GS-cPass”) was performed according to the manufacturer’s instructions. Photometric measurements were performed using the Tecan Sunrise (Tecan, Männedorf, Switzerland). Binding inhibition was calculated in percentage.

**Micro neutralisation:**
Manual micro virus neutralisation assay (NT) analysis was performed in 96-well culture plates (Greiner bio-one, Frickenhausen, Germany) as previously described, with the exception that confluent cells were incubated instead of adding cells after the neutralisation reaction.[1] Samples with a titre < 1:10 dilution were classified as "NT negative" and samples with a titre ≥1:10 were classified as "NT positive". In brief, virus stocks (50 TCID/50 µl) were prepared from SARS-CoV-2 cultured samples (1st passage, strain MUC IMB-1, cultured in Vero E6 cells) and stored at -80° C for later use. Plasma samples were diluted twofold from of 1:10 to 1:80 in Minimal Essential Medium (MEM, plus Non-Essential Amino Acids Solution and Antibiotic-Antimycotic Solution; all Invitrogen, Thermo Fisher Scientific, Darmstadt, Germany) and tested in duplicates. On each plate, a known positive and a known negative plasma sample were tested as controls. In addition, a mock control and a virus back-titration were added to each plate. The subsequently diluted plasma samples were pre-incubated with virus for one hour

at 37°C in 5% $CO_2$ and 95% humidity. The suspension was then transferred into the wells with the confluent Vero-E6 cells. After 72 hours of incubation at 37°C (5% $CO_2$), the supernatants was discarded and the plates were fixed and stained in a solution of 13% formalin/PBS plus 0.1% crystal violet. The highest plasma dilution showing complete inhibition of CPE in both microtitre plate wells corresponded to the NAbs titre. Samples with a titre < 1:10 were classified as "NT negative" and samples with a titre ≥1:10 were classified as "NT positive".

**VIRAMED SARS-CoV-2 ViraChip® microarray:**
SARS-CoV-2 ViraChip® microarray (VIRAMED Biotech AG, Planegg, Germany; hereafter called VC-N-IgA/IgM/IgG; VC-S1-IgA/IgM/IgG; VC-S2-IgA/IgM/IgG) was based on an enzyme-immunoassay for the qualitative detection of IgG, IgA or IgM antibodies against the specific SARS-CoV-2 recombinant antigens S1, S2, and N in human serum. The assay was performed on a Dynex ELISA Processor DSX® (Dynex Technologies, Denkendorf, Germany) according to the manufacturer’s instructions. We have reported the values provided by the automated ELISA processor, which are in arbitrary units.

**Mikrogen *recom*Line SARS-CoV-2 IgG line immunoassay:**
The *recom*Line SARS-CoV-2 IgG line immunoassay (Mikrogen, Neuried, Germany; hereafter named MG-S1, MG-N, MG-RBD) was based on nitrocellulose strips with recombinant SARS-CoV-2 antigens S1, N and RBD as well as recombinant N antigens derived from seasonal human CoV NL63, OC43, 229E, and HKU1. The assay for the qualitative detection of human IgG against the respective antigens was performed using the fully automated *recom*Line strip processor Carl (Mikrogen, Neuried, Germany) according to the manufacturer’s instructions. The raw values presented are in arbitrary units. The test results below the cut-off of 1 are categorised as negative and information regarding their quantitative values is comitted.

#### Statistical Analysis

##### Optimisation of cut-off values

Using the raw values of both the primary and confirmatory tests from the true-positive and true-negative cohorts, several statistical techniques were applied to identify optimised cut-offs thresholds and calculate the performance of classifiers based on single and combinations of tests.

The EI-S1-IgA, EI-S1-IgG, and Ro-N−Ig tests were available for 1,266 subjects with known SARS-CoV-2 status (table 1 and supplemental table 2). For better comparability, 18 samples that had no available result for Ro-N-Ig were omitted. As these tests were also conducted on all KoCo19 study subjects, the results for classifiers based on these tests could be used to predict the serological status of the Munich cohort and to adjust the crude seroprevalence estimates based on the specificity and sensitivity of these classifiers.[2] The sample sizes available for the confirmatory tests are given in table 1 and supplemental table 2.

For each of the single tests, a nonparametric bootstrap approach was used to optimise the cut-off threshold as well as estimate the corresponding specificity, sensitivity and overall prediction accuracy. More precisely, 10,000 bootstrap samples were drawn (with replacement, without balancing positive/negative true test outcomes). For each, a cut-off was determined that maximised the overall prediction accuracy. We refer to cut-offs obtained in single bootstrap samples as bootstrap cut-offs to distinguish them from the final optimised cut-off. The latter is eventually obtained by taking the median of 10,000 bootstrap cut-offs.

Some of the tests yield left-censored measurements, i.e., values below a detection limit are not resolved but summarised in one category. To determine each bootstrap cut-off, we replaced the left-censored values in each bootstrap test with a random realisation using a uniform distribution between zero and the detection limit. The optimisation with the resulting virtual uncensored sample can yield bootstrap cut-offs above the detection limit or below. While the former values were accepted, the latter underwent post-processing as they cannot be implemented in serological studies. For bootstrap cut-offs below the detection limit we evaluated the classification accuracy for the interval bounds – i.e., zero and detection limit – which could be implemented. The bounds yielding a higher prediction accuracy was used as bootstrap cut-off for the respective bootstrap sample.

For each test, the bootstrap cut-off was used to predict the serological status of the observations not included in the respective bootstrap sample (out-of-bag observations). After considering the predictions for each observation over all bootstrap replications, majority votes were chosen as final single predictions. These predictions were compared to the true serological status to calculate specificity, sensitivity and overall prediction accuracy. To this end, the optimised cut-off and the three performance criteria were estimated based on different data, namely, the former was estimated based on bootstrap samples and the latter were estimated based on the respective out-of-bag observations.

The performance of the optimised cut-off threshold was compared to the performance of the cut-off determined by the manufacturer. Since the manufacturer calibrated the cut-off using their own data, no bootstrap was applied to this: The three performance indices were computed by comparing the true serological status to the predictions based on the cut-off for the entire sample. Overall, the calculation of the performance indices for both optimised and manufacturer's cut-offs rely on out-of-sample predictions. The difference in the validation data sets may lead to differences in performance indices even if both the optimised and the manufacturer's cut-offs are the same. We know that while the term "optimised cut-offs" suggest that they are better, it can be that the manufacturer cut-offs reach be change a higher sensitivity, specificity or overall accuracy on the dataset. This can happen as the optimisation is performed on the bootstrapped data, is however only the case for situations which both cut-offs anyhow perform very similar.

To evaluate the potential benefit of combining several primary tests, we trained a random forest and a support vector machine which took the measurements for all three primary tests (EI-S1-IgA, EI-S1-IgG, and Ro-N−Ig) as inputs In order to tuning the parameters for the random forest, the number of selected variables at each split and the minimum node size were taken into account. For the support vector machine, a Gaussian radial basis function kernel along with the kernel coefficient gamma and the penalty parameter of the error term as tuning parameters were selected. For both the random forest and the support vector machine, a grid search was applied to find the combination of the two tuning parameters that minimised the out-of-bag prediction error resulting from 1,000 bootstrap samples. The final estimates for the performance criteria were then obtained by running the respective classifier with the optimised tuning parameters using 2,000 bootstrap replicates and the majority votes from the resulting out-of-bag predictions, as described above regarding the classifiers based on a single test.

**Software**

For analyses and visualisations, we used the R software, version 4.0.2. For data visualisation, we used in particular the packages "ggplot2"[3] and "RainCloudPlots"[4].

#### References

1. Haselmann V, Özçürümez MK, Klawonn F, et al. Results of the first pilot external quality assessment (EQA) scheme for anti-SARS-CoV2-antibody testing. *Clinical chemistry and laboratory medicine* 2020.
2. Pritsch M, et al. Prevalence and risk factors of infection in the representative COVID-19 cohort Munich. *Adjacent manuscript*.
3. Wickham H. ggplot2: elegant graphics for data analysis: springer; 2016.
4. Allen M, Poggiali D, Whitaker K, Marshall T, Kievit R. Raincloud plots: a multi-platform tool for robust data visualization. Wellcome Open Res. 4, 63. 2019.

| Nature of cohort | Recruitment and study details | Sample numbers and definitions | Days from positive PCR median (min-max; mean) |
|---|---|---|---|
| Healthy blood donors (=known SARS-CoV-2 negatives) | October 2019 | Total n=500<br>*True-positives = 0*<br>*True-negatives = 500*<br>*Unknown = 0* | NA |
| | March 2020 *(after common cold season)* | Total n=491<br>*True-positives = 0*<br>*True-negatives = 491*<br>*Unknown = 0* | NA |
| SARS-CoV-2 infected subjects (=known SARS-CoV-2 positives) | Volunteers >18 yrs that tested positive for SARS-CoV-2 in Munich (156 households) | Total n=193<br>*True-positives = 193*<br>*True-negatives = 0*<br>*Unknown = 0* | n=187<br><br>median 99 days (7-126; mean = 86) |
| Representative population-based study & substudies (Prospective COVID-19 Cohort Munich; KoCo19), | 3,003 randomly selected Munich households (April-June 2020) & 92 households from substudies | Total n=5474<br>*True-positives = 0*<br>*True-negatives = 100*<br>*Unknown = 5374* | NA |
| **Total samples used** | **Total n = 6658** | ***True-positives = 193***<br>***True-negatives = 1091***<br>***Unknown = 5374*** | |

***Supplemental Table 1*: Cohort details.**
Characterisation of study participants. Subjects with a positive RT-PCR were considered as true-positives, while blood donors (sampled in the pre-COVID-19 era) were classified as true-negatives. In addition, we included subjects recruited into the Representative COVID-19 Cohort Munich (KoCo19), 100 of which were considered as true-negatives. For subjects with several longitudinal measurements, the blood sample with the most complete dataset was retained. For similar datasets, the earliest measurement was considered. For operational replicates, the latest measurement was used.

| Test | Sample size | Positive result optim. / manuf. cut-off | Negative result optim. / manuf. cut-off | True-positives | True-negatives |
|---|---|---|---|---|---|
| EI-S1-IgA | 6,657 | 687 / 674 | 5970 / 5983 | 193 | 1091 |
| EI-S1-IgG | 6,658 | 321 / 309 | 6337 / 6349 | 193 | 1091 |
| RO-N-Ig | 6,636 | 307 / 289 | 6329 / 6347 | 193 | 1073 |
| NT | 355 | 148* | 207 | 107 | 106 |
| GS-cPass | 355 | 197 / 197 | 158 / 158 | 108 | 106 |
| VC-N-IgA | 361 | NA / 26 | NA / 355 | 108 | 110 |
| VC-N-IgM | 362 | NA / 18 | NA / 344 | 108 | 111 |
| VC-N-IgG | 362 | 125 / 81 | 237 / 281 | 108 | 111 |
| VC-S1-IgA | 361 | NA / 37 | NA / 324 | 108 | 110 |
| VC-S1-IgM | 362 | NA / 17 | NA / 345 | 108 | 111 |
| VC-S1-IgG | 362 | 201 / 134 | 161 / 228 | 108 | 111 |
| VC-S2-IgA | 361 | NA / 29 | NA / 332 | 108 | 110 |
| VC-S2-IgM | 362 | NA / 15 | NA / 347 | 108 | 111 |
| VC-S2-IgG | 362 | 145 / 37 | 217 / 225 | 108 | 111 |
| MG-NP | 273 | 139 / 139 | 134 / 134 | 78 | 106 |
| MG-RBD | 273 | 137 / 137 | 136 / 136 | 78 | 106 |
| MG-S1 | 273 | 141 / 141 | 132 / 132 | 78 | 106 |
| 229E | 273 | NA / 129 | NA / 144 | 78 | 106 |
| NL63 | 273 | NA / 127 | NA / 146 | 78 | 106 |
| OC43 | 273 | NA / 79 | NA / 194 | 78 | 106 |
| HKU1 | 273 | NA / 130 | NA / 143 | 78 | 106 |

***Supplemental Table 2*****: Number and description of tests.**

For subjects with multiple blood samples and test results from different time points, measurements were excluded (only one value per individual and test system). Confirmatory tests were performed on a subset of samples. Samples from true-positives and true-negatives were used as controls for confirmatory tests.

* For NT, dilutions starting at 1:10 were used (see Methods).

| **Assay** | **Instrument(s) used** | **Readout** |
|---|---|---|
| EI-S1-IgG | Euroanalyzer-1 (Euroimmun, Lübeck, Germany) | quotient of the optical density |
| EI-S1-IgA | Euroanalyzer-1 (Euroimmun, Lübeck, Germany) | quotient of the optical density |
| Ro-N-Ig | cobas 400-e411 (Roche, Basel, Switzerland) | cut-off-index (COI) |
| | cobas 8000-e801 (Roche, Basel, Switzerland) | cut-off-index (COI) |
| NT | NA | dilution |
| GS-cPass | Tecan Sunrise (Tecan, Männedorf, Switzerland) | inhibition [%] |
| VC-N-IgA/IgM/IgG | Dynex ELISA Processor DSX® (Dynex Technologies, Denkendorf, Germany) | arbitrary unit |
| VC-S1-IgA/IgM/IgG | Dynex ELISA Processor DSX® (Dynex Technologies, Denkendorf, Germany) | arbitrary unit |
| VC-S2-IgA/IgM/IgG | Dynex ELISA Processor DSX® (Dynex Technologies, Denkendorf, Germany) | arbitrary unit |
| MG-S1 | recomLine strip processor Carl (Mikrogen, Neuried, Germany) | quotient and category (<1) |
| MG-N | recomLine strip processor Carl (Mikrogen, Neuried, Germany) | quotient and category (<1) |
| MG-RBD | recomLine strip processor Carl (Mikrogen, Neuried, Germany) | quotient and category (<1) |

***Supplemental Table 3*: Platform used and units given for the respective assays**

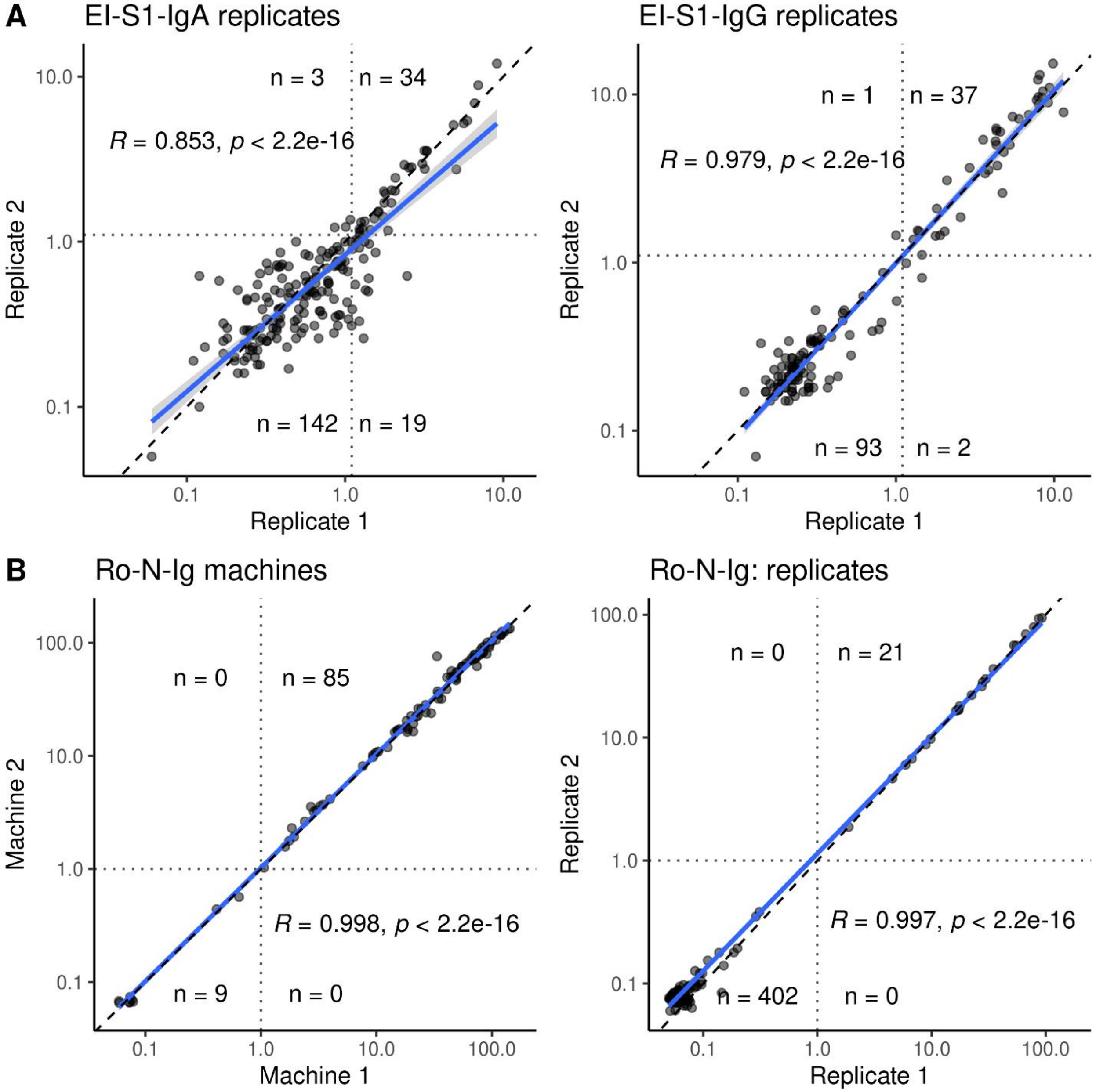
A
EI-S1-IgA replicates
n = 3
n = 34
R = 0.853, p < 2.2e-16
n = 142
n = 19
Replicate 2
Replicate 1
10.0
1.0
0.1
EI-S1-IgG replicates
n = 1
n = 37
R = 0.979, p < 2.2e-16
n = 93
n = 2
Replicate 2
Replicate 1
B
Ro-N-Ig machines
n = 0
n = 85
R = 0.998, p < 2.2e-16
n = 9
n = 0
Machine 2
Machine 1
100.0
Ro-N-Ig: replicates
n = 0
n = 21
R = 0.997, p < 2.2e-16
n = 402
n = 0
Replicate 2
Replicate 1

***Supplemental Figure 1*: Reliability analysis of operational replicates .**

Discrepant/non-discrepant results derived from the same blood samples measured in two replicates.

(A) Scatterplot of replicates of EI-S1-IgA (left) and EI-S1-IgG(right). The correlation for EI-S1-IgA was R=0.853, but classification was discrepant in 22 cases (11.1%). The correlation for EI-S1-IgG was higher with R=0.979, with 3 subjects changing category (2.3%). Replicates for both tests were performed on the same platform; however, with varying lot numbers and operators.

(B) Scatterplot of replicates of Ro-N-Ig between modules e411 (machine 1) and e801 (machine 2) (left), and on module e801 (right). Although the 511 samples were tested on two different modules with kits of varying lot numbers and operators, the correlation was R=0.99, with not a single discrepant classification.

**Suppl.Fig.2A**

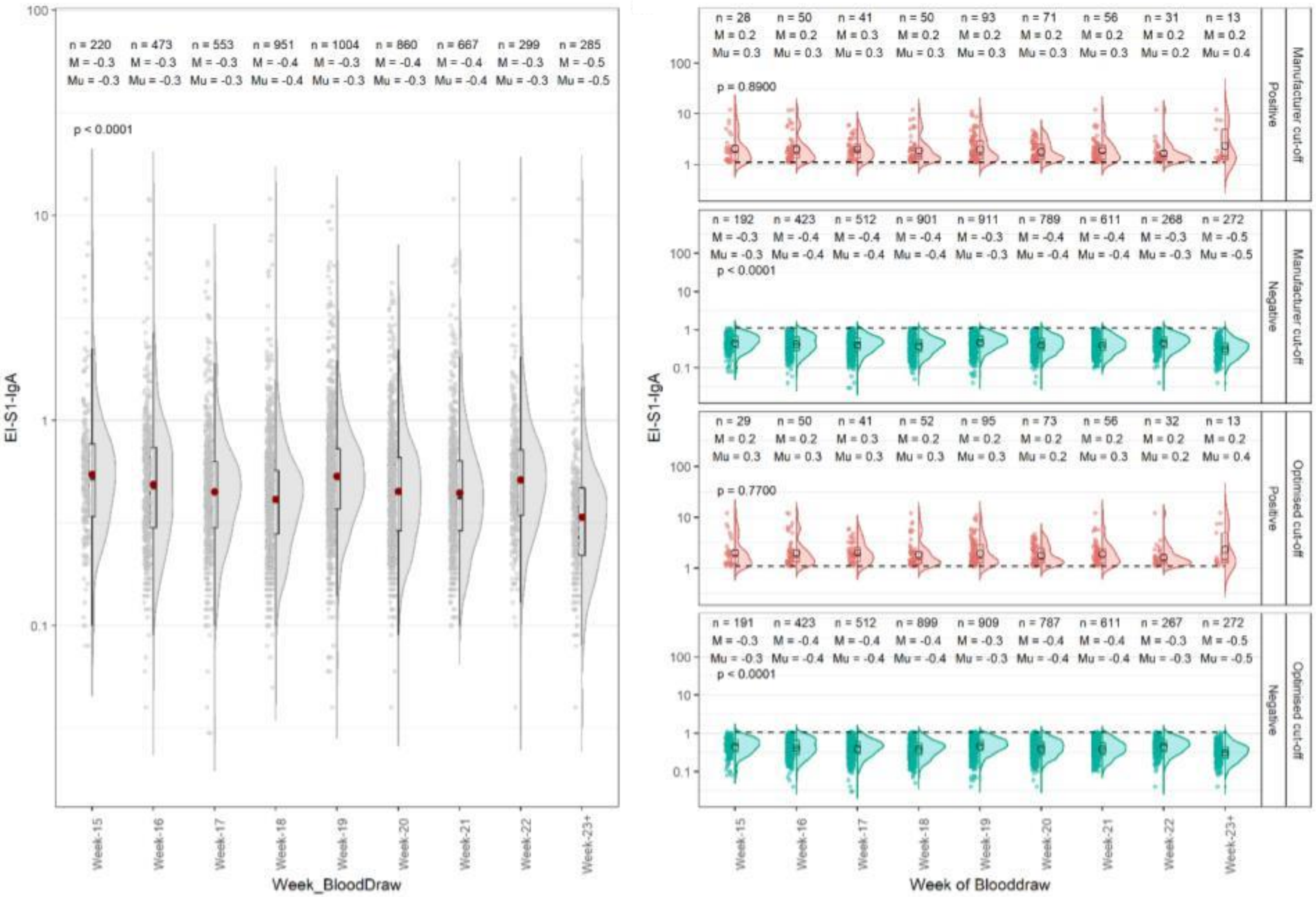

n = 220 M = -0.3 Mu = -0.3
n = 473 M = -0.3 Mu = -0.3
n = 553 M = -0.3 Mu = -0.3
n = 951 M = -0.4 Mu = -0.4
n = 1004 M = -0.3 Mu = -0.3
n = 860 M = -0.4 Mu = -0.3
n = 667 M = -0.4 Mu = -0.4
n = 299 M = -0.3 Mu = -0.3
n = 285 M = -0.5 Mu = -0.5
p < 0.0001
EI-S1-IgA
Week_BloodDraw
Week-15
Week-16
Week-17
Week-18
Week-19
Week-20
Week-21
Week-22
Week-23+
Manufacturer cut-off
Positive
n = 28 M = 0.2 Mu = 0.3
n = 50 M = 0.2 Mu = 0.3
n = 41 M = 0.3 Mu = 0.3
n = 50 M = 0.2 Mu = 0.3
n = 93 M = 0.2 Mu = 0.3
n = 71 M = 0.2 Mu = 0.3
n = 56 M = 0.2 Mu = 0.3
n = 31 M = 0.2 Mu = 0.2
n = 13 M = 0.2 Mu = 0.4
p = 0.8900
Manufacturer cut-off
Negative
n = 192 M = -0.3 Mu = -0.3
n = 423 M = -0.4 Mu = -0.4
n = 512 M = -0.4 Mu = -0.4
n = 901 M = -0.4 Mu = -0.4
n = 911 M = -0.3 Mu = -0.3
n = 789 M = -0.4 Mu = -0.4
n = 611 M = -0.4 Mu = -0.4
n = 268 M = -0.3 Mu = -0.3
n = 272 M = -0.5 Mu = -0.5
p < 0.0001
Optimised cut-off
Positive
n = 29 M = 0.2 Mu = 0.3
n = 50 M = 0.2 Mu = 0.3
n = 41 M = 0.3 Mu = 0.3
n = 52 M = 0.2 Mu = 0.3
n = 95 M = 0.2 Mu = 0.3
n = 73 M = 0.2 Mu = 0.2
n = 56 M = 0.2 Mu = 0.3
n = 32 M = 0.2 Mu = 0.2
n = 13 M = 0.2 Mu = 0.4
p = 0.7700
Optimised cut-off
Negative
n = 191 M = -0.3 Mu = -0.3
n = 423 M = -0.4 Mu = -0.4
n = 512 M = -0.4 Mu = -0.4
n = 899 M = -0.4 Mu = -0.4
n = 909 M = -0.3 Mu = -0.3
n = 787 M = -0.4 Mu = -0.4
n = 611 M = -0.4 Mu = -0.4
n = 267 M = -0.3 Mu = -0.3
n = 272 M = -0.5 Mu = -0.5
p < 0.0001
EI-S1-IgA
Week of Blooddraw

**Suppl.Fig.2B**

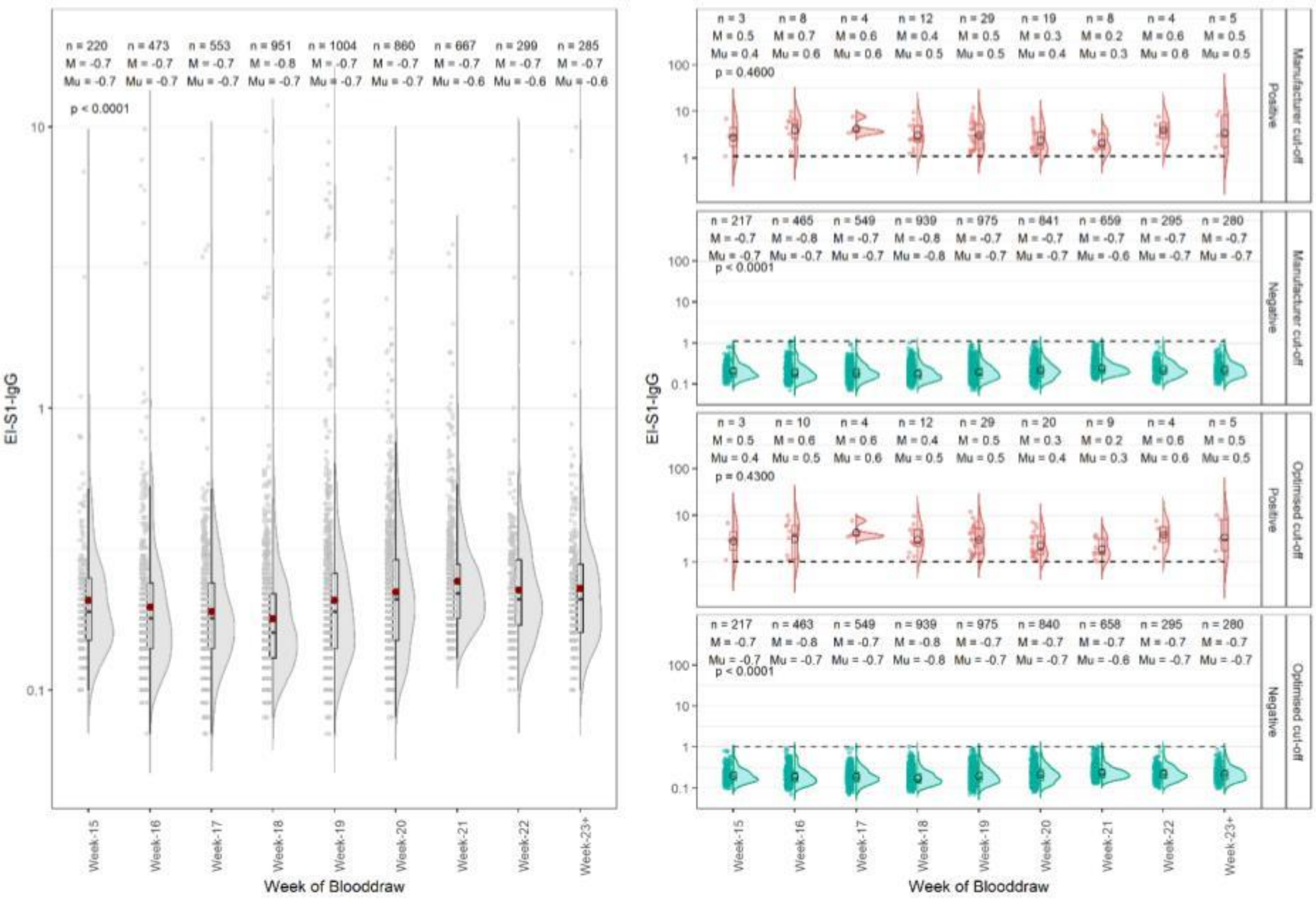
n = 220
M = -0.7
Mu = -0.7
n = 473
M = -0.7
Mu = -0.7
n = 553
M = -0.7
Mu = -0.7
n = 951
M = -0.8
Mu = -0.7
n = 1004
M = -0.7
Mu = -0.7
n = 860
M = -0.7
Mu = -0.7
n = 667
M = -0.7
Mu = -0.6
n = 299
M = -0.7
Mu = -0.6
n = 285
M = -0.7
Mu = -0.6
p < 0.0001
EI-S1-IgG
Week-15
Week-16
Week-17
Week-18
Week-19
Week-20
Week-21
Week-22
Week-23+
Week of Blooddraw
p = 0.4600
p < 0.0001
p = 0.4300
p < 0.0001
Positive
Negative
Manufacturer cut-off
Optimised cut-off
EI-S1-IgG
Week of Blooddraw

**Suppl.Fig.2C**

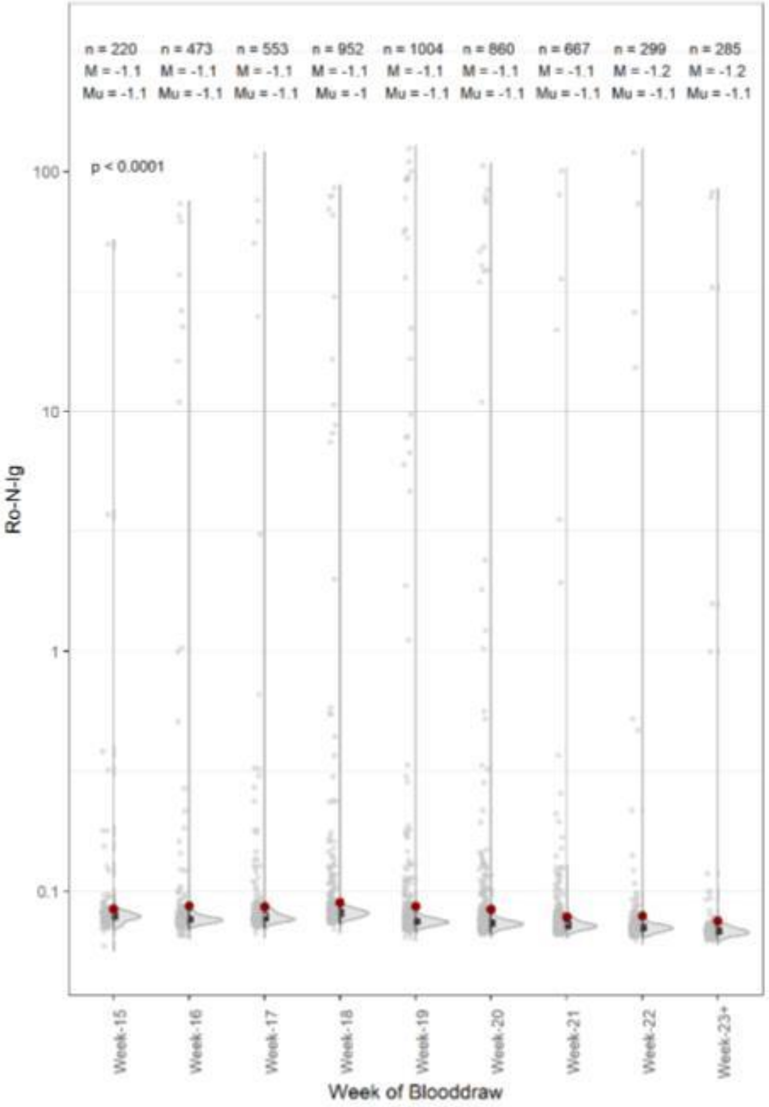
n = 220 M = -1.1 Mu = -1.1
n = 473 M = -1.1 Mu = -1.1
n = 553 M = -1.1 Mu = -1.1
n = 952 M = -1.1 Mu = -1
n = 1004 M = -1.1 Mu = -1.1
n = 860 M = -1.1 Mu = -1.1
n = 667 M = -1.1 Mu = -1.1
n = 299 M = -1.2 Mu = -1.1
n = 285 M = -1.2 Mu = -1.1
p < 0.0001
Ro-N-Ig
Week-15
Week-16
Week-17
Week-18
Week-19
Week-20
Week-21
Week-22
Week-23+
Week of Blooddraw

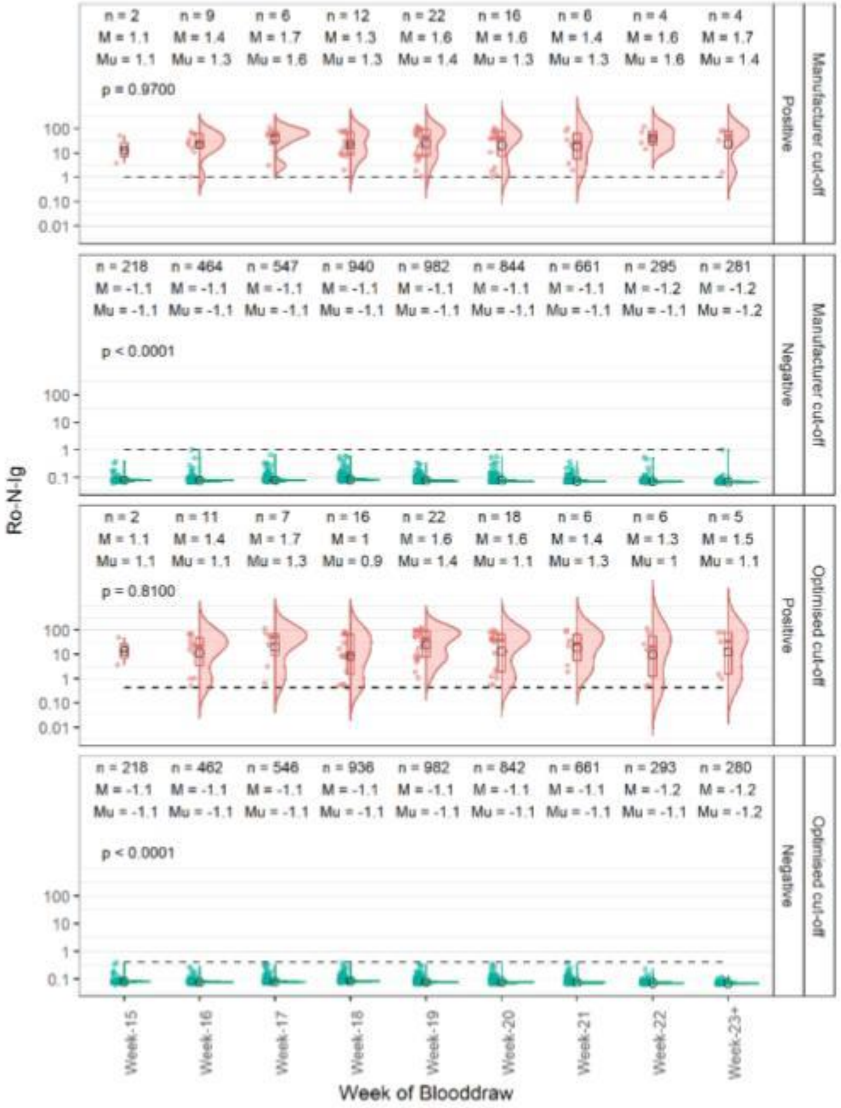
Manufacturer cut-off
Positive
p = 0.9700
Negative
p < 0.0001
Optimised cut-off
p = 0.8100
Ro-N-Ig
Week of Blooddraw

***Supplemental Figure 2*: Temporal analysis by recruitment week.**

The time-dependent distribution of the read-out values was investigated to exclude test inaccuracies due to cross-reactivity of assays for SARS-COV-2 with common cold coronavirus or other seasonal effects. The distribution of all raw values (in grey on the left side), as well as an individual analysis of positively (red) and negatively (green) tested samples was evaluated. Distributions are depicted for the manufacturer's cut-off (first two figures, upper right) as well as the optimised cut-off (last two figures, lower right). $n$ denotes the count of the outcomes, $M$ the median (also observed as the horizontal line in the boxplot) and $Mu$ the mean (denoted by the red dot) for each calendar week in the study. Our analyses confirm that within our sample set, the measured background does not vary considerably within the prospective sample period of the KoCo19 study.

(A) Values obtained with EI-S1-IgA
(B) Values obtained with EI-S1-IgG
(C) Values obtained with Ro-N-Ig

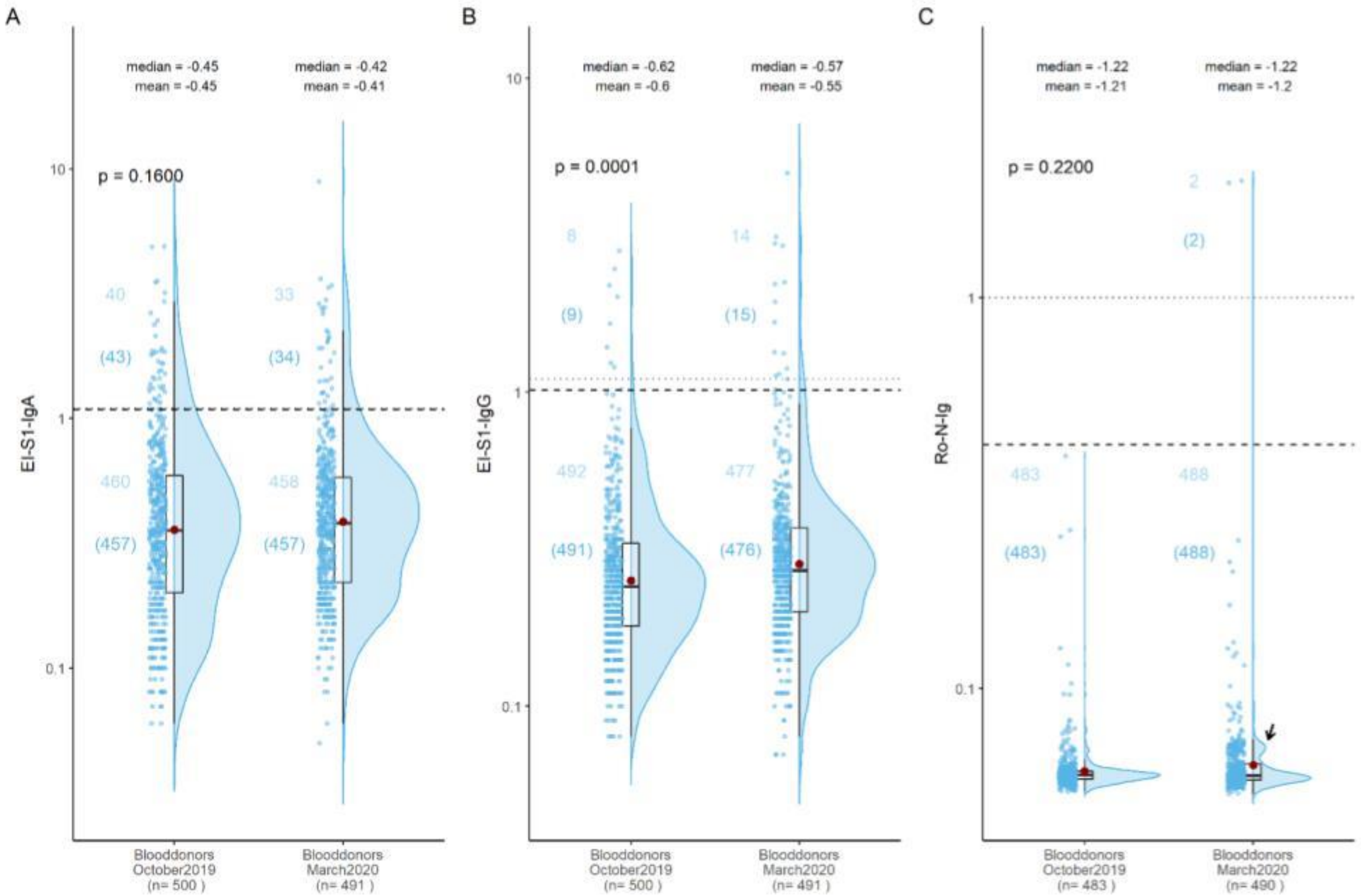
A
median = -0.45
mean = -0.45
median = -0.42
mean = -0.41
p = 0.1600
40
(43)
33
(34)
460
(457)
458
(457)
EI-S1-IgA
10
1
0.1
Blooddonors
October2019
(n= 500 )
Blooddonors
March2020
(n= 491 )
B
median = -0.62
mean = -0.6
median = -0.57
mean = -0.55
p = 0.0001
8
(9)
14
(15)
492
(491)
477
(476)
EI-S1-IgG
10
1
0.1
Blooddonors
October2019
(n= 500 )
Blooddonors
March2020
(n= 491 )
C
median = -1.22
mean = -1.21
median = -1.22
mean = -1.2
p = 0.2200
2
(2)
483
(483)
488
(488)
Ro-N-Ig
1
0.1
Blooddonors
October2019
(n= 483 )
Blooddonors
March2020
(n= 490 )

***Supplemental Figure 3*: Blood donor data over time.**
Measurement values for (A) EI-S1-IgA, (B) EI-S1-IgE, and (C) Ro-N-Ig in the true-negative blood donor cohorts from autumn 2019 and spring 2020. Counts n refer to the number of observations above/below manufacturer's and optimised cut-off for each of the temporal groups (without brackets: manufacturers' cut-offs; within brackets: optimised cut-offs). Pairwise differences were considered only after adjusting for multiple testing.
(A) For EI-S1-IgA, a trend towards higher baseline values in the spring cohort (p=0.16) presented,
(B) for EI-S1-IgG, this increase was significant (p=0.0001).
(C) For Ro-N-Ig, there was a slight but non-significant trend towards increased baseline values (p = 0.22); however, a second population with increased reactivity presented above the negative population. This sub-population seemed to increase in spring (arrow). All samples above the manufacturer's cut-off values for any of the tests were tested negative in direct virus neutralization, suggesting a non-specific reactivity.

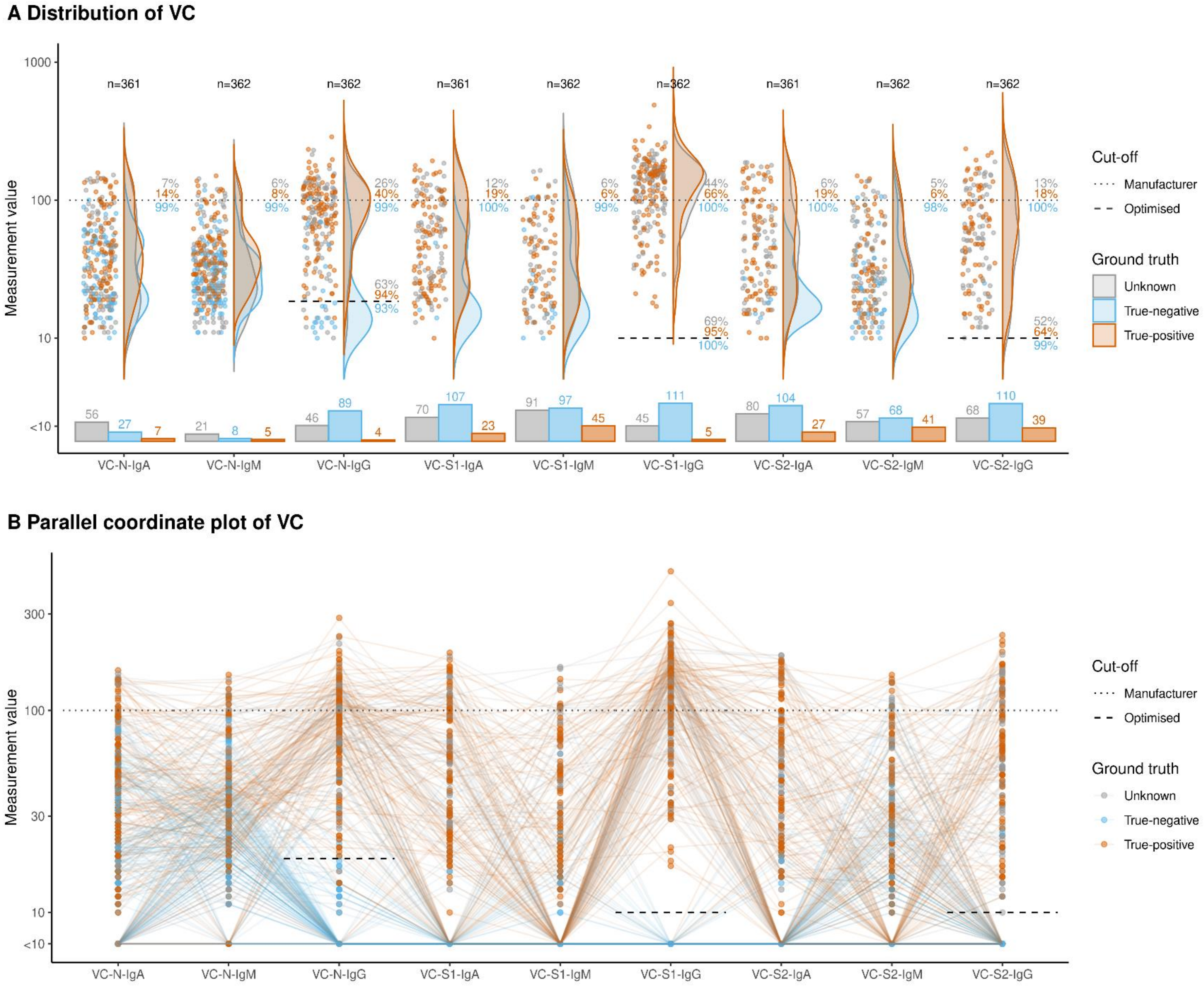
A Distribution of VC
Measurement value
n=361
n=362
n=362
n=361
n=362
n=362
n=361
n=362
n=362
VC-N-IgA
VC-N-IgM
VC-N-IgG
VC-S1-IgA
VC-S1-IgM
VC-S1-IgG
VC-S2-IgA
VC-S2-IgM
VC-S2-IgG
Cut-off
Manufacturer
Optimised
Ground truth
Unknown
True-negative
True-positive
B Parallel coordinate plot of VC
Measurement value

***Supplemental Figure 4*: Distribution and comparison of VC.**

Parallel coordinate plot of the test results for true-negatives (blue), true-positives (orange), and individual with unknown SARS-CoV-2 status (grey). Black dotted and dashed lines represent the original manufacturer's and the optimised positivity cut-offs. Orange (blue) numbers give the percentage of true-positives (true-negatives) correctly detected by the test. The orange values above the dotted line represents the percentage of positive test results for the true positive cohort, the blue number below demonstrate the percentages of negatives in the true negative cohort. Grey numbers indicate the percentages of positive samples with unknown SARS-CovV-2. These percentages were calculated over the total number of samples with unknown SARS-CoV-2 with available test results. Bar charts below violin plots represent the information for the categorical part of the test.

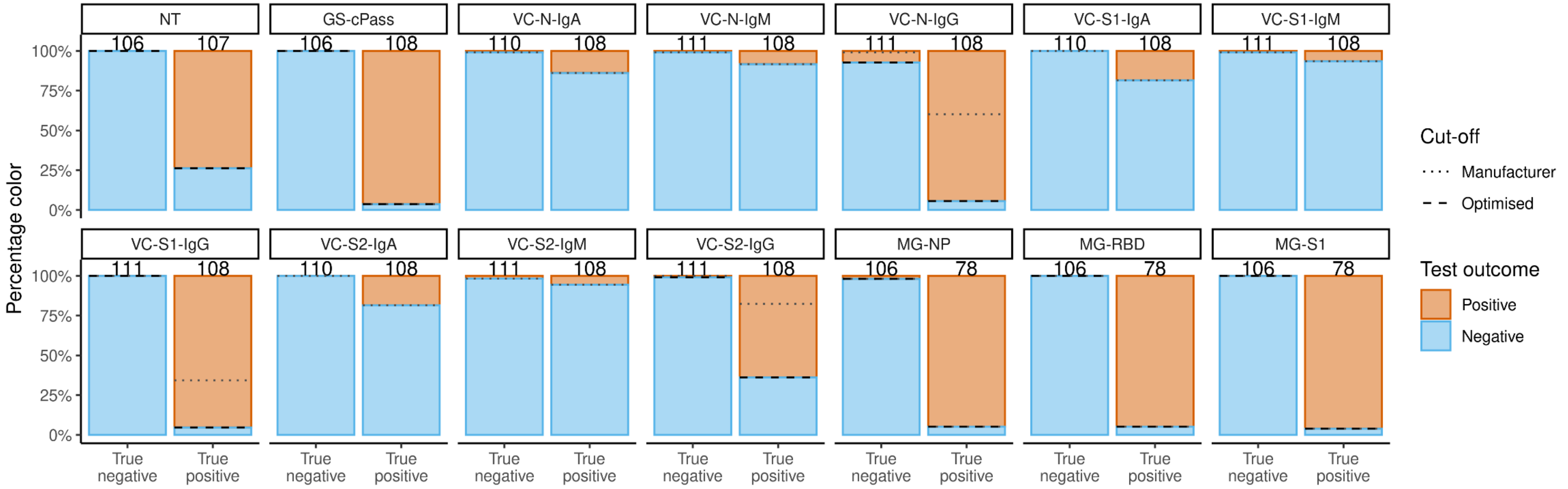


***Supplemental Figure 5: Description and comparison of performances of confirmatory tests.***
Test results according to ground truth colour coded by optimised cut-off for the primary tests. The dotted lines represent the manufacturer's cut-offs, the dashed lines the optimised cut-offs defined within this study.
Best overall performance was obtained with GS-cPass, MG-RBD, and MG-S1 as well as VC-S1-IgG. Direct NT presented as 100% specific (1:10 dilution), with a decreased sensitivity compared to other tests used. VC-N-IgG and VC-S1-IgG improve applying optimised cut-offs.

**Suppl.Fig.6A**

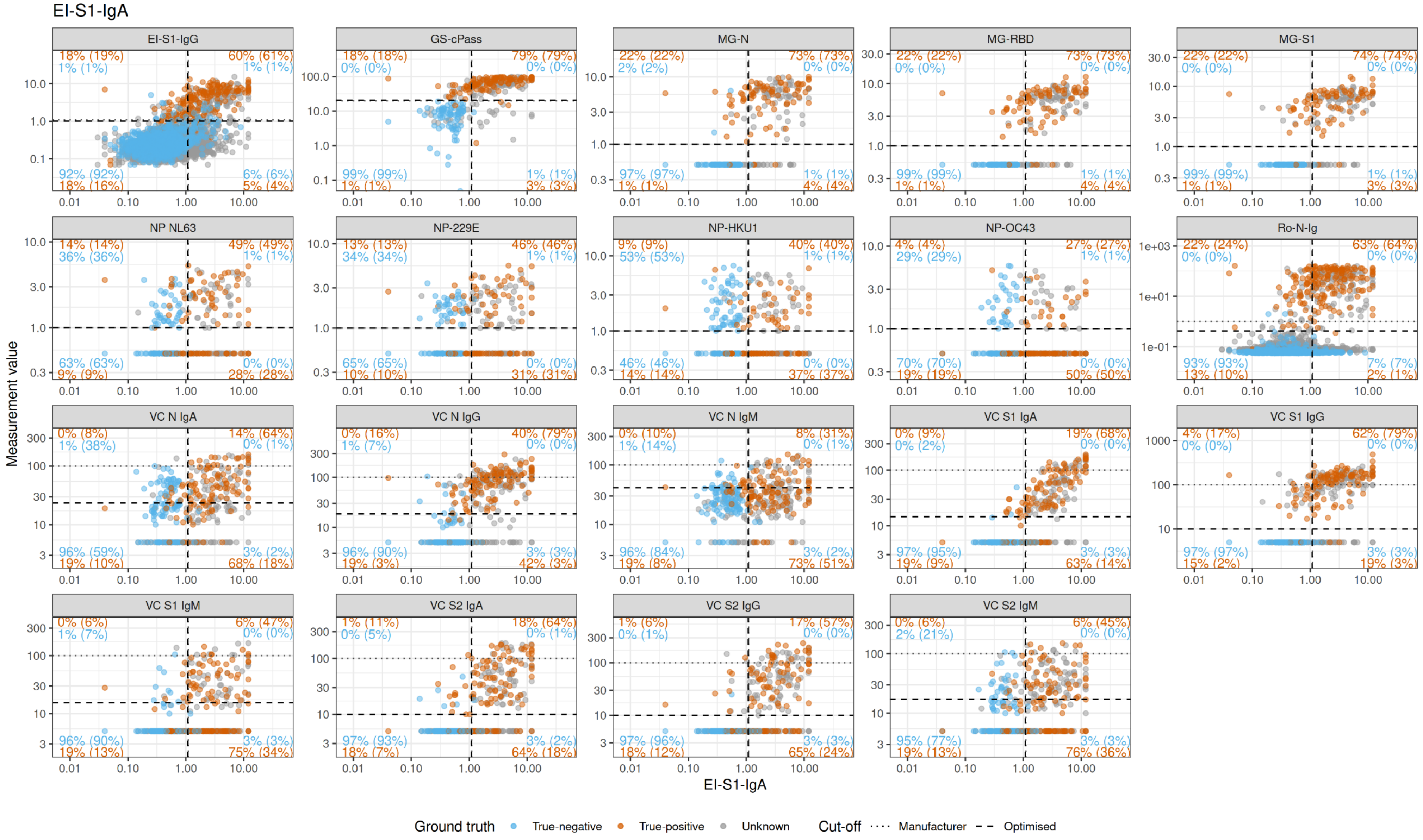
EI-S1-IgA
EI-S1-IgG
GS-cPass
MG-N
MG-RBD
MG-S1
NP NL63
NP-229E
NP-HKU1
NP-OC43
Ro-N-Ig
VC N IgA
VC N IgG
VC N IgM
VC S1 IgA
VC S1 IgG
VC S1 IgM
VC S2 IgA
VC S2 IgG
VC S2 IgM
Measurement value
EI-S1-IgA
Ground truth
True-negative
True-positive
Unknown
Cut-off
Manufacturer
Optimised

**Suppl.Fig.6B**

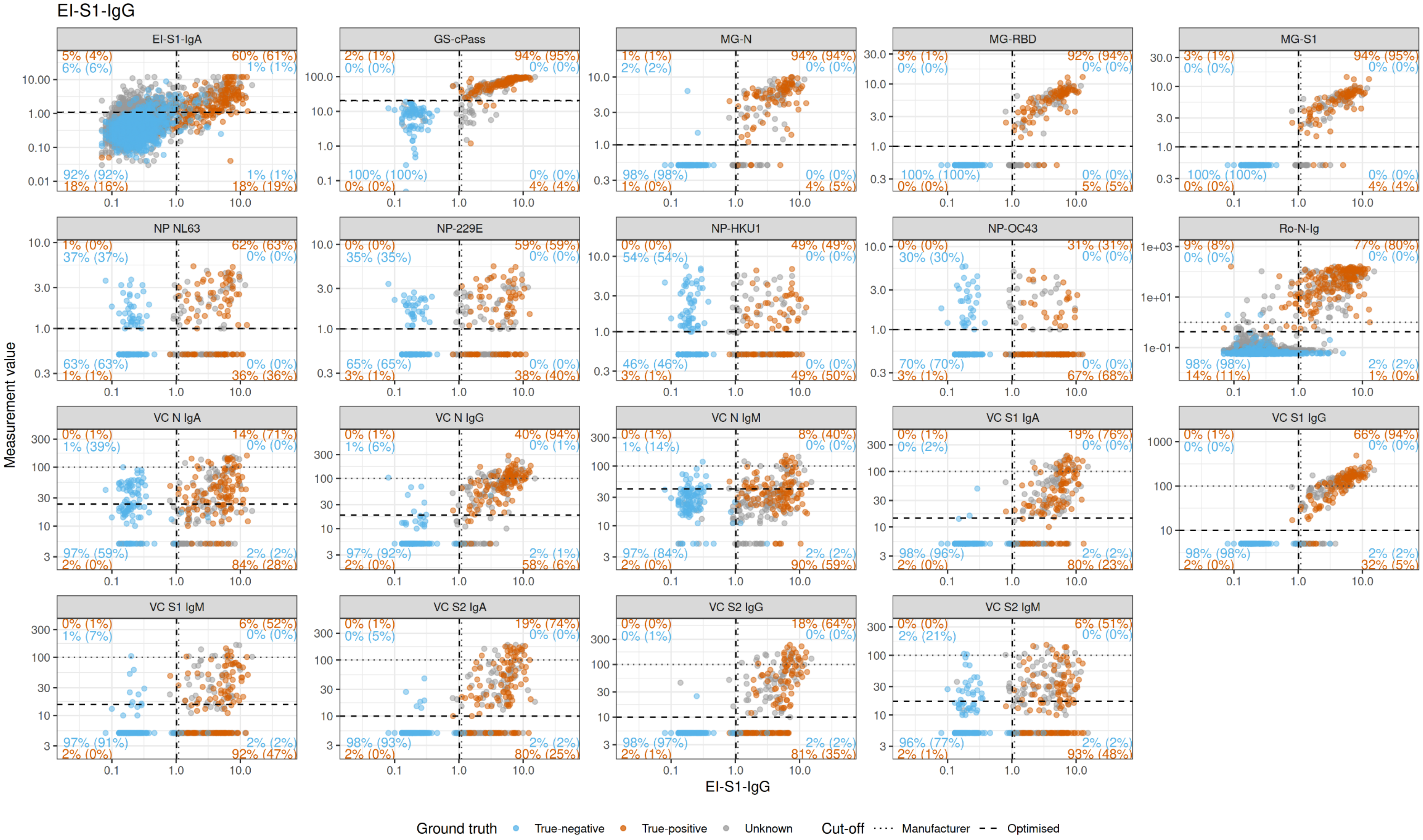
EI-S1-IgG
EI-S1-IgA
5% (4%) 6% (6%) 60% (61%) 1% (1%) 92% (92%) 18% (16%) 1% (1%) 18% (19%)
GS-cPass
2% (1%) 0% (0%) 94% (95%) 0% (0%) 100% (100%) 0% (0%) 0% (0%) 4% (4%)
MG-N
1% (1%) 2% (2%) 94% (94%) 0% (0%) 98% (98%) 1% (0%) 0% (0%) 4% (5%)
MG-RBD
3% (1%) 0% (0%) 92% (94%) 0% (0%) 100% (100%) 0% (0%) 0% (0%) 5% (5%)
MG-S1
3% (1%) 0% (0%) 94% (95%) 0% (0%) 100% (100%) 0% (0%) 0% (0%) 4% (4%)
NP NL63
1% (0%) 37% (37%) 62% (63%) 0% (0%) 63% (63%) 1% (1%) 0% (0%) 36% (36%)
NP-229E
0% (0%) 35% (35%) 59% (59%) 0% (0%) 65% (65%) 3% (1%) 0% (0%) 38% (40%)
NP-HKU1
0% (0%) 54% (54%) 49% (49%) 0% (0%) 46% (46%) 3% (1%) 0% (0%) 49% (50%)
NP-OC43
0% (0%) 30% (30%) 31% (31%) 0% (0%) 70% (70%) 3% (1%) 0% (0%) 67% (68%)
Ro-N-Ig
9% (8%) 0% (0%) 77% (80%) 0% (0%) 98% (98%) 14% (11%) 2% (2%) 1% (0%)
VC N IgA
0% (1%) 1% (39%) 14% (71%) 0% (0%) 97% (59%) 2% (0%) 2% (2%) 84% (28%)
VC N IgG
0% (1%) 1% (6%) 40% (94%) 0% (1%) 97% (92%) 2% (0%) 2% (1%) 58% (6%)
VC N IgM
0% (1%) 1% (14%) 8% (40%) 0% (0%) 97% (84%) 2% (0%) 2% (2%) 90% (59%)
VC S1 IgA
0% (1%) 0% (2%) 19% (76%) 0% (0%) 98% (96%) 2% (0%) 2% (2%) 80% (23%)
VC S1 IgG
0% (1%) 0% (0%) 66% (94%) 0% (0%) 98% (98%) 2% (0%) 2% (2%) 32% (5%)
VC S1 IgM
0% (1%) 1% (7%) 6% (52%) 0% (0%) 97% (91%) 2% (0%) 2% (2%) 92% (47%)
VC S2 IgA
0% (1%) 0% (5%) 19% (74%) 0% (0%) 98% (93%) 2% (0%) 2% (2%) 80% (25%)
VC S2 IgG
0% (0%) 0% (1%) 18% (64%) 0% (0%) 98% (97%) 2% (1%) 2% (2%) 81% (35%)
VC S2 IgM
0% (0%) 2% (21%) 6% (51%) 0% (0%) 96% (77%) 2% (1%) 2% (2%) 93% (48%)
Measurement value
EI-S1-IgG
Ground truth
True-negative
True-positive
Unknown
Cut-off
Manufacturer
Optimised

**Suppl.Fig.6C**

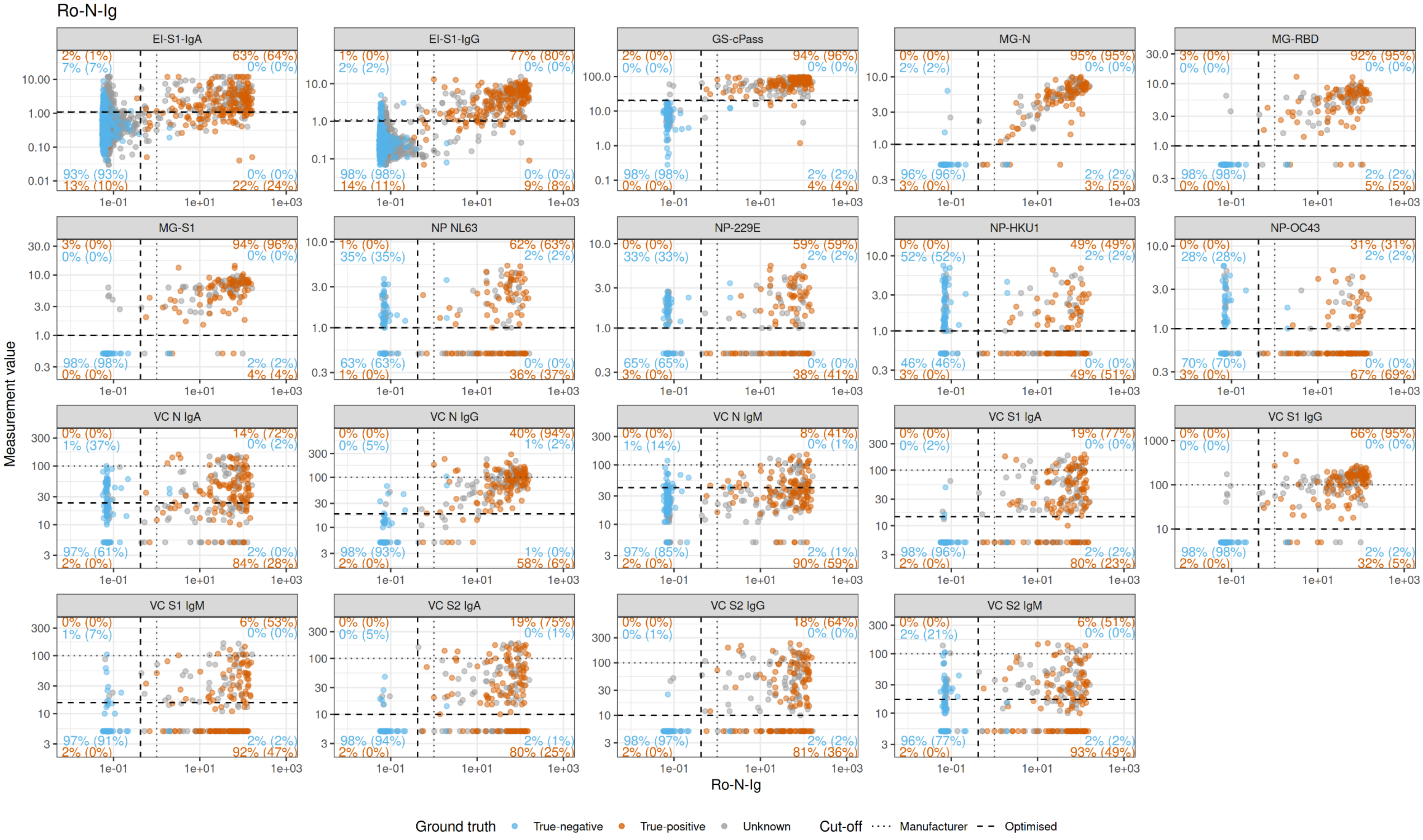
Ro-N-Ig
EI-S1-IgA
EI-S1-IgG
GS-cPass
MG-N
MG-RBD
MG-S1
NP NL63
NP-229E
NP-HKU1
NP-OC43
VC N IgA
VC N IgG
VC N IgM
VC S1 IgA
VC S1 IgG
VC S1 IgM
VC S2 IgA
VC S2 IgG
VC S2 IgM
Measurement value
Ro-N-Ig
Ground truth
True-negative
True-positive
Unknown
Cut-off
Manufacturer
Optimised

***Supplemental Figure 6: Scatterplots of confirmatory tests vs primary (using optimised and manufacturer's cut-off).***

True-negatives in blue, true-positives in orange, unknown SARS-CoV-2 status in grey. Black dotted and dashed lines represent the manufacturer's and the optimised positivity cut-offs. Orange/blue numbers indicate the percentages of true-positives/negatives correctly detected. The orange value above the dotted line represents the percentage of positive test results for the true-positive cohort, the blue number below is the percentage of negatives in the true-negative cohort.

(A) Values obtained with EI-S1-IgA
(B) Values obtained with EI-S1-IgG
(C) Values obtained with Ro-N-Ig

**Suppl.Fig.7A**

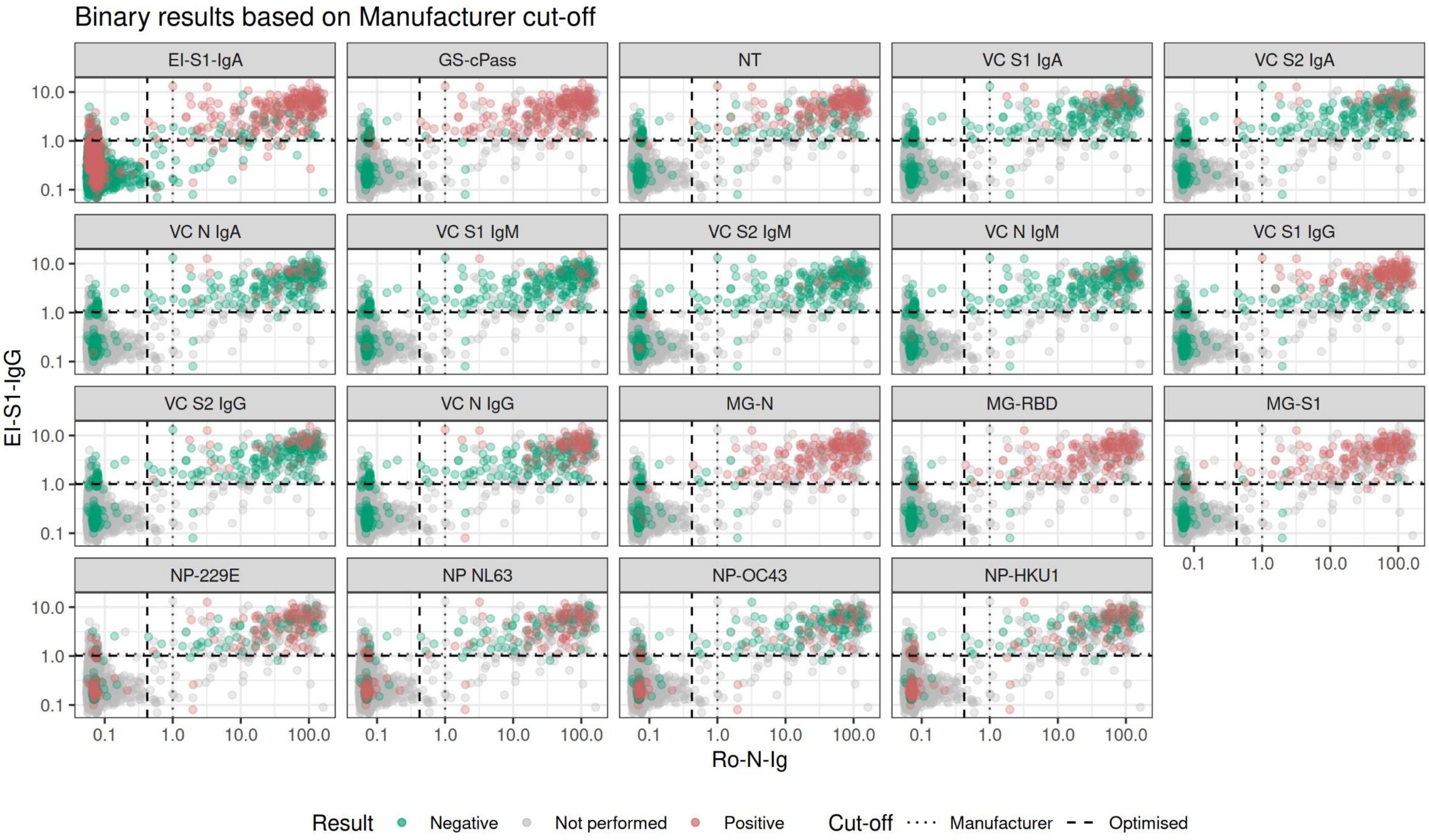
Binary results based on Manufacturer cut-off
EI-S1-IgA
GS-cPass
NT
VC S1 IgA
VC S2 IgA
VC N IgA
VC S1 IgM
VC S2 IgM
VC N IgM
VC S1 IgG
VC S2 IgG
VC N IgG
MG-N
MG-RBD
MG-S1
NP-229E
NP NL63
NP-OC43
NP-HKU1
EI-S1-IgG
10.0
1.0
0.1
Ro-N-Ig
0.1
1.0
10.0
100.0
Result
Negative
Not performed
Positive
Cut-off
Manufacturer
Optimised

**Suppl.Fig.7B**

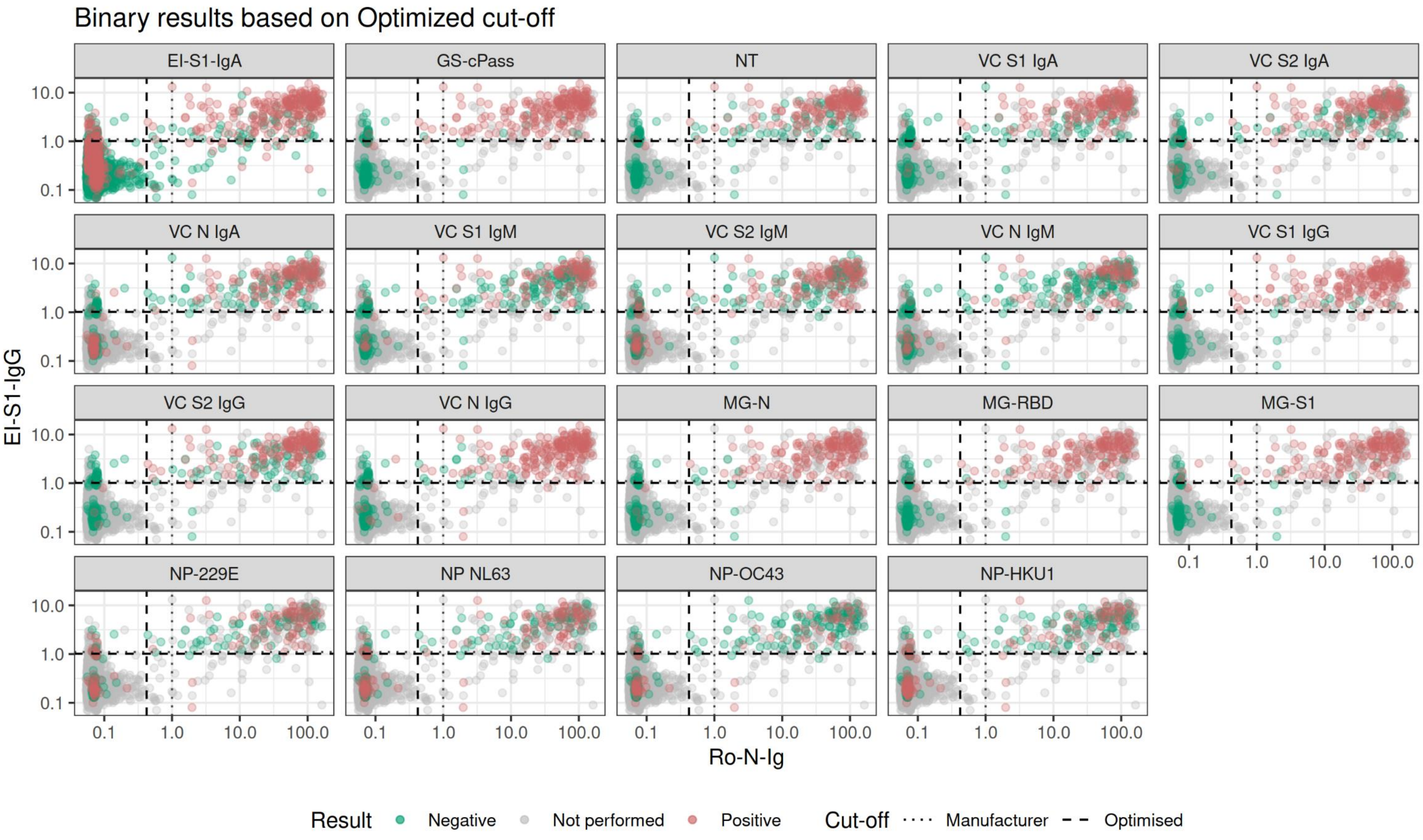
Binary results based on Optimized cut-off
EI-S1-IgA
GS-cPass
NT
VC S1 IgA
VC S2 IgA
VC N IgA
VC S1 IgM
VC S2 IgM
VC N IgM
VC S1 IgG
VC S2 IgG
VC N IgG
MG-N
MG-RBD
MG-S1
NP-229E
NP NL63
NP-OC43
NP-HKU1
10.0
1.0
0.1
0.1
1.0
10.0
100.0
EI-S1-IgG
Ro-N-Ig
Result
Negative
Not performed
Positive
Cut-off
Manufacturer
Optimised

***Supplemental Figure 7: Ro-N-Ig vs. EI-S1-IgG showing binary results for the tests***
(A) Manufacturer's cut-off
(B) Optimised cut-off

Disclaimer: this is a pre-print manuscript and has not been peer-reviewed yet

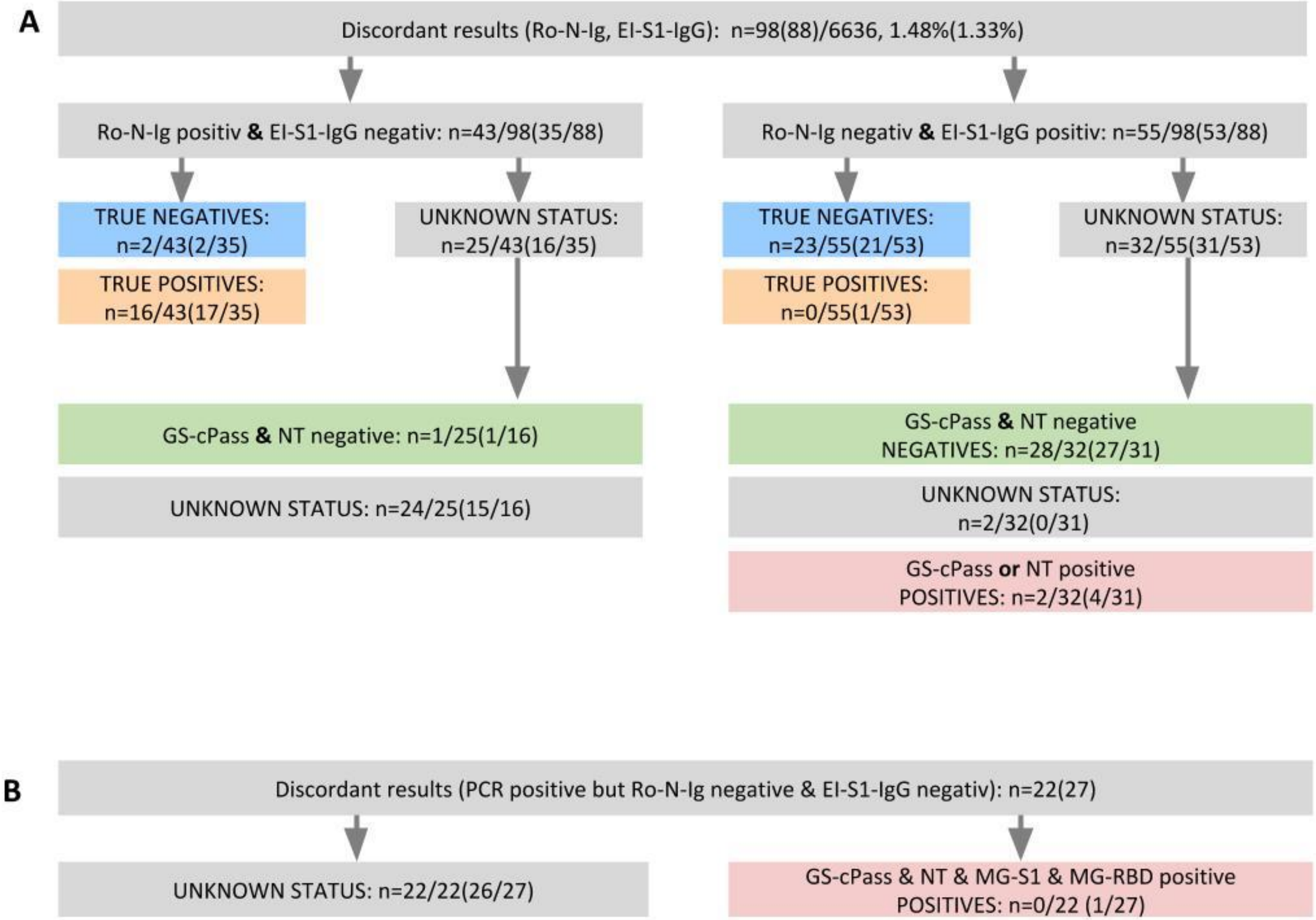


***Supplemental Figure 8: Analysis of discordant results.***

Values in brackets correspond to the manufacturer's cut-off, other values were determined with optimized cut-offs where appropriate.

(A) Discordance between EI-S1-IgG and Ro-N-Ig (n=98). Left branch: 43/98 cases tested positive in Ro-N-Ig but negative in EI-S1-IgG; 18/43 could be specified as true-positives/negatives; from the unknown status category, one sample presented negative in both GS-cPass and NT; 24/98 cases remained unexplained. Right branch: 55/98 cases tested negative in Ro-N-Ig but positive in EI-S1-IgG; 23/55 could be specified as true-positives; for 30/32 cases, further specifications demonstrated 28/32 to be negatives (GS-cPass and NT negative) and 2/32 to be positives (GS-cPass or NT positive).

(B) Discordance between RT-PCR positive result, but negatively tested in EI-S1-IgG and Ro-N-Ig. All 22/98 cases remained unexplained. For one person the RT-PCR date was not available.

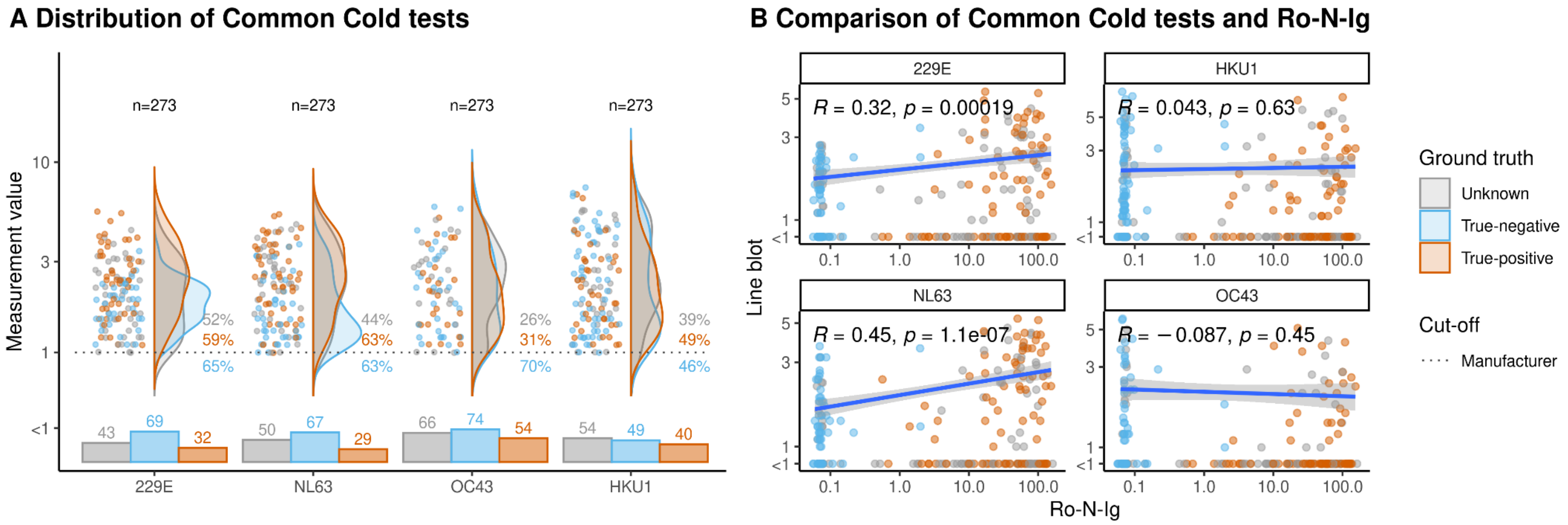


***Supplemental Figure 9: Common cold CoV line blot correlation***

(A) The distribution of common cold CoV line-blot raw values for true-positives, true-negatives and unknowns. The orange value above the dotted line represents the percentage of positive test results for true-positives, the blue number is the percentage of negatives in true-negatives. Grey numbers indicate the percentages of positive samples with unknown SARS-CoV-2 determined by manufacturer's and optimised cut-offs. These percentages were calculated over the total number of samples with unknown SARS-CoV-2 with available test results. Bar charts below violin plots represent the information for the categorical part of the test. No association between OC43 and HKU1 positivity is apparent, neither for SARS-CoV-2 positivity or negativity. In contrast, for 229E and NL63 the true-positive subjects show significantly higher raw values, translating into higher positivity rates than the true-negative subjects.

(B) Ro-N-Ig as a surrogate for anti SARS-CoV-2 N-protein reactivity correlated with anti N-protein reactivity for 229E, NL63, OC43, and HKU1. For 229E and NL63, very high line blot raw values presented solely in combination with high raw values in Ro-N-Ig (red circles), although a general linear association was not apparent. This was not the case for OC43 and HKU1: here, the true-negative subjects were distributed wider and reached high values in the line blot, without any reactivity in Ro-N-Ig.

**Suppl.Fig.10A**

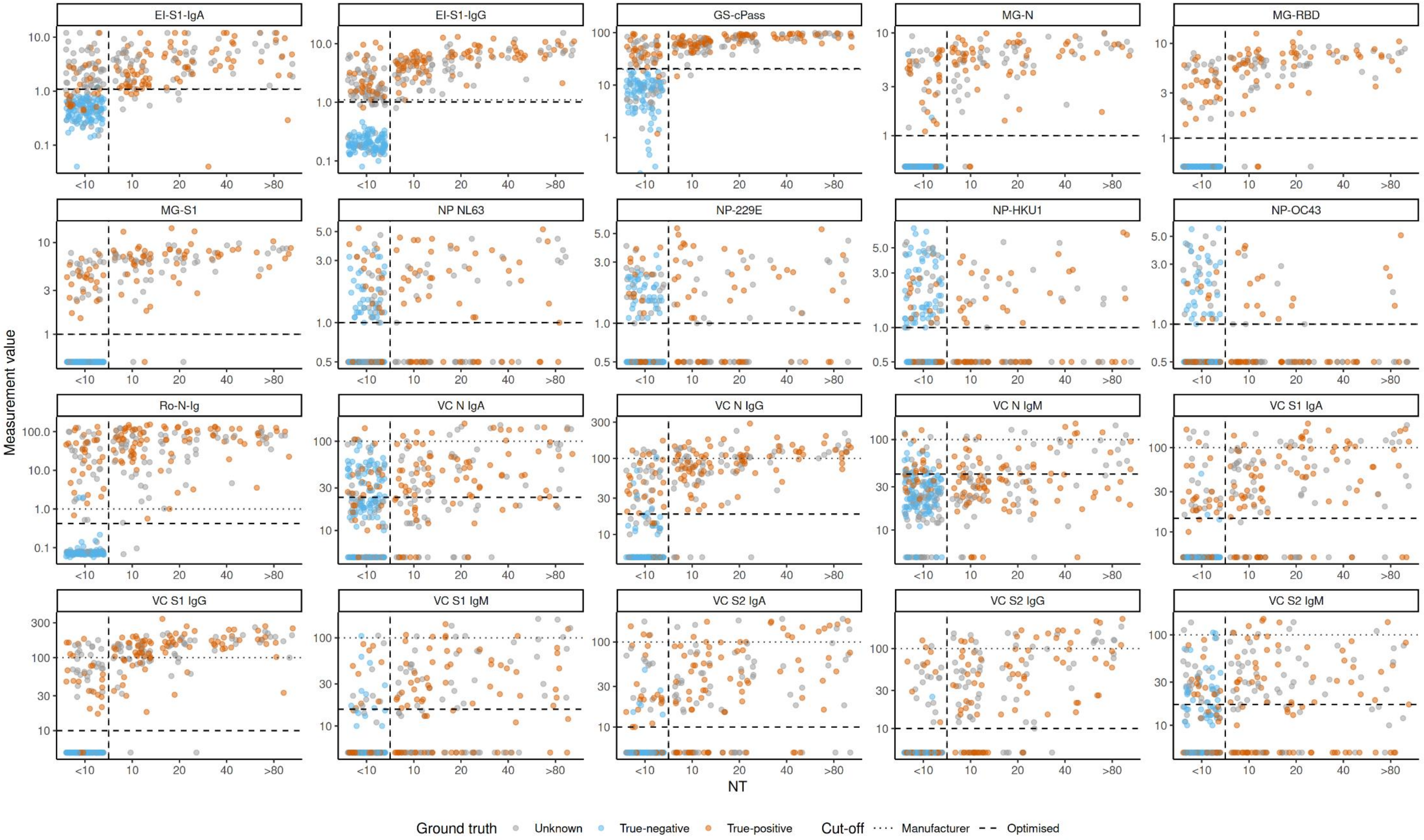

**Suppl.Fig.10B**

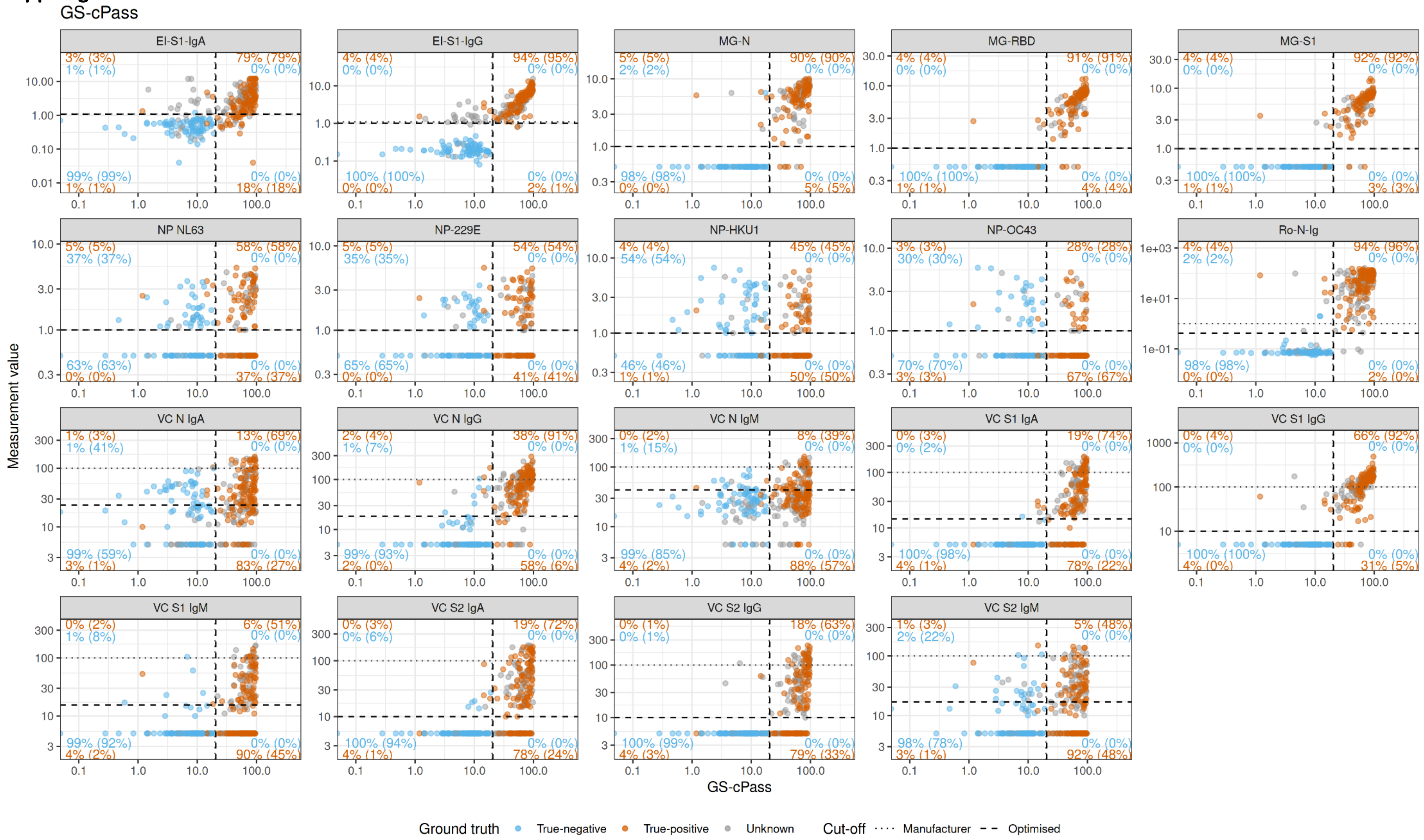

**Suppl.Fig.10C**

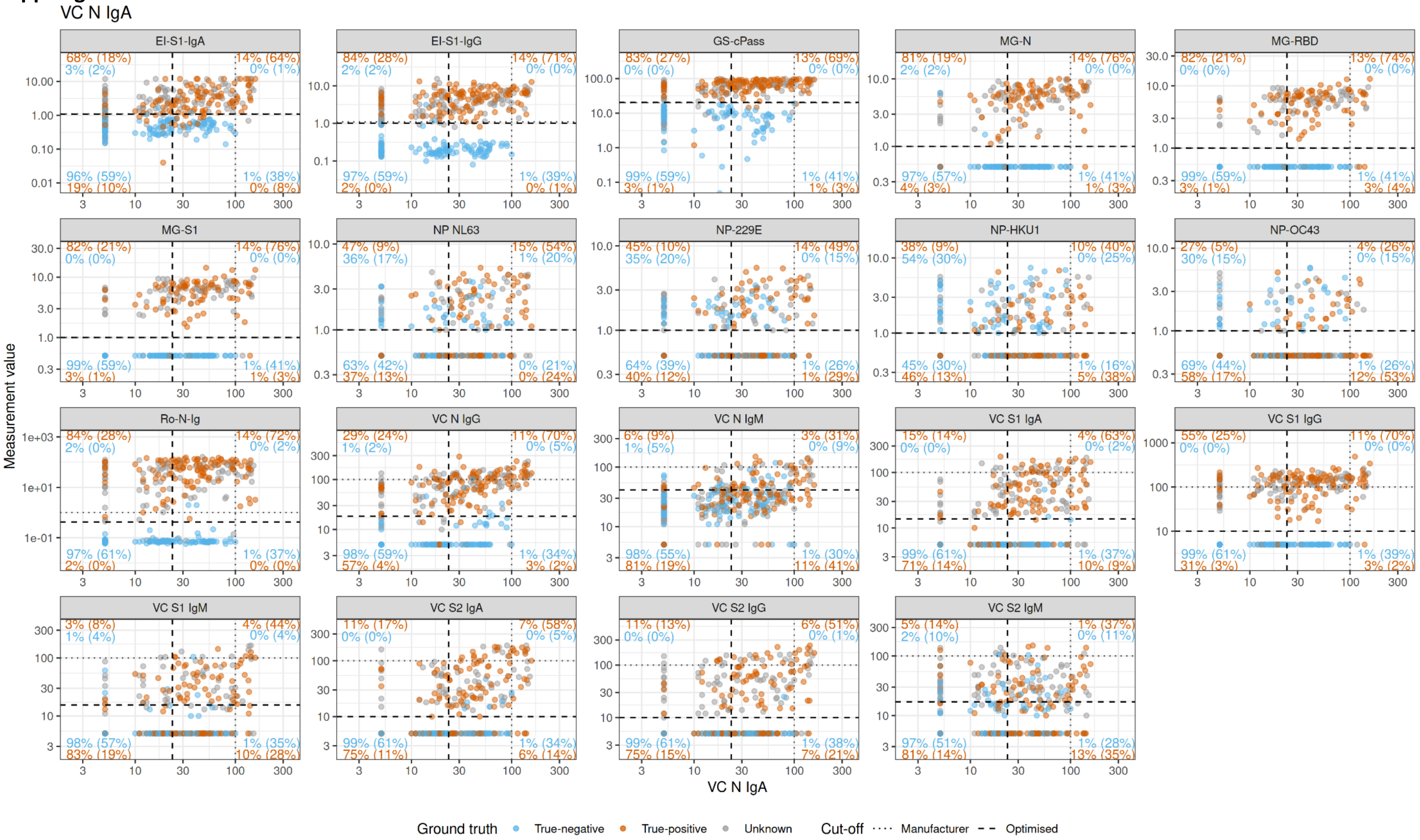

**Suppl.Fig.10D**

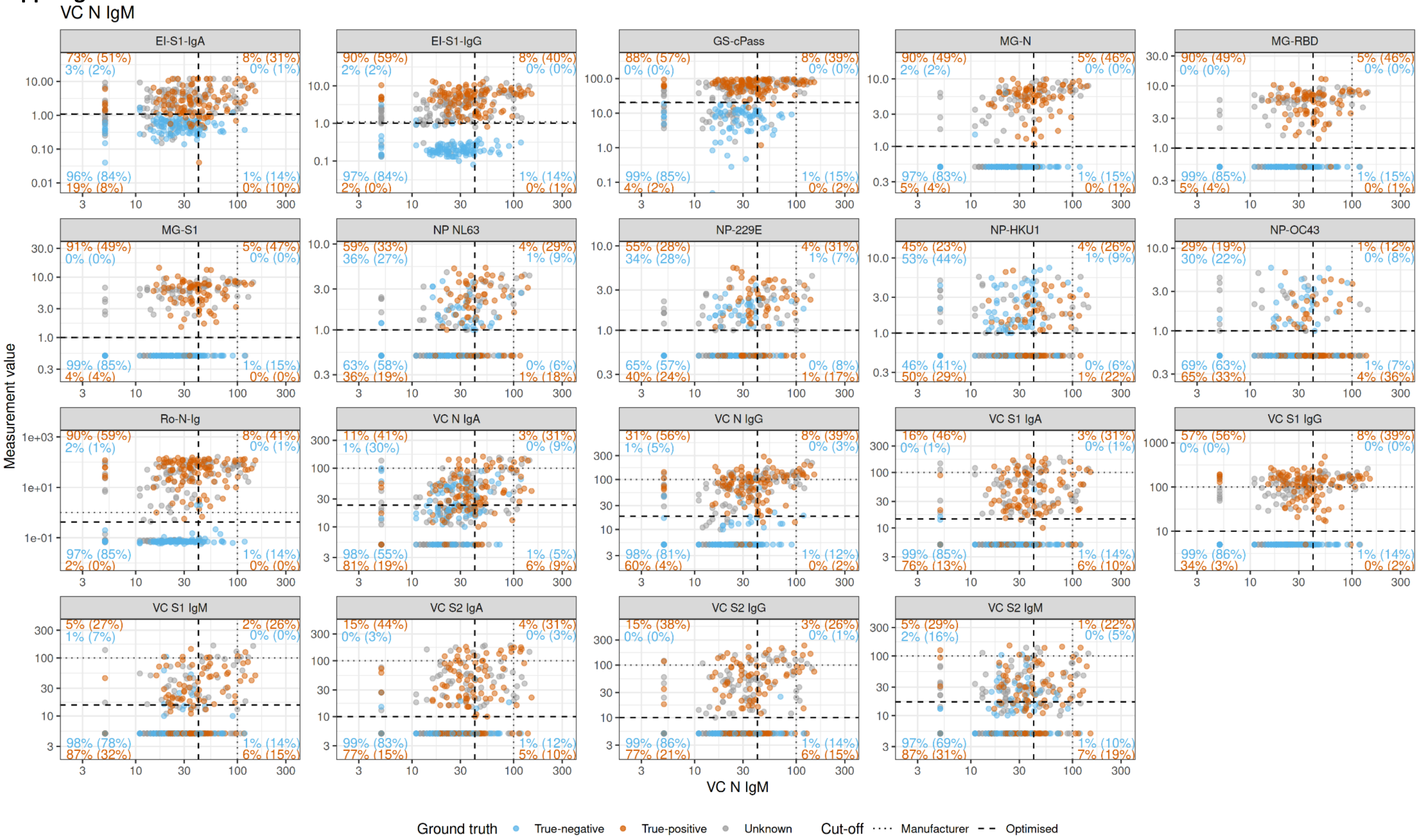

**Suppl.Fig.10E**

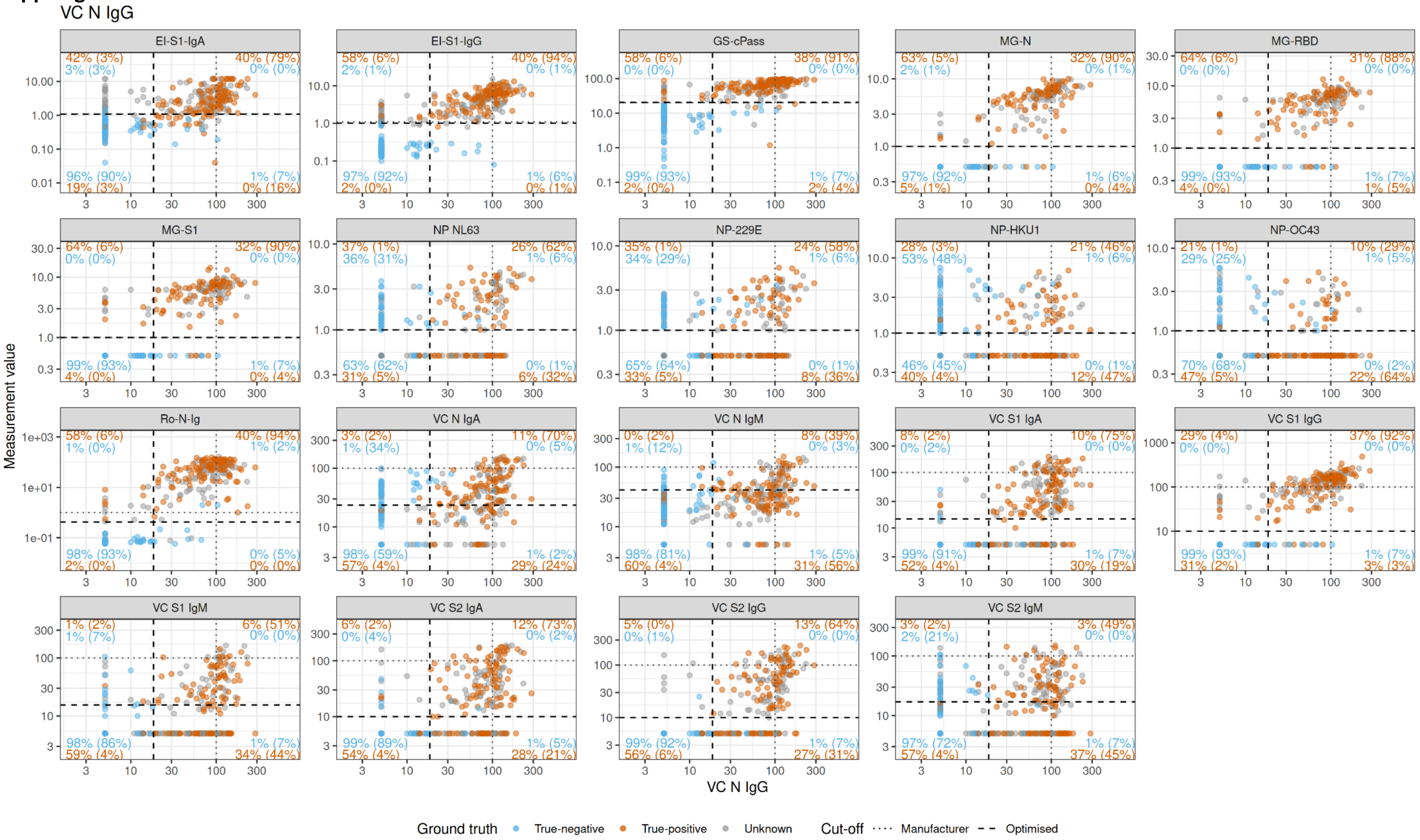

**Suppl.Fig.10F**

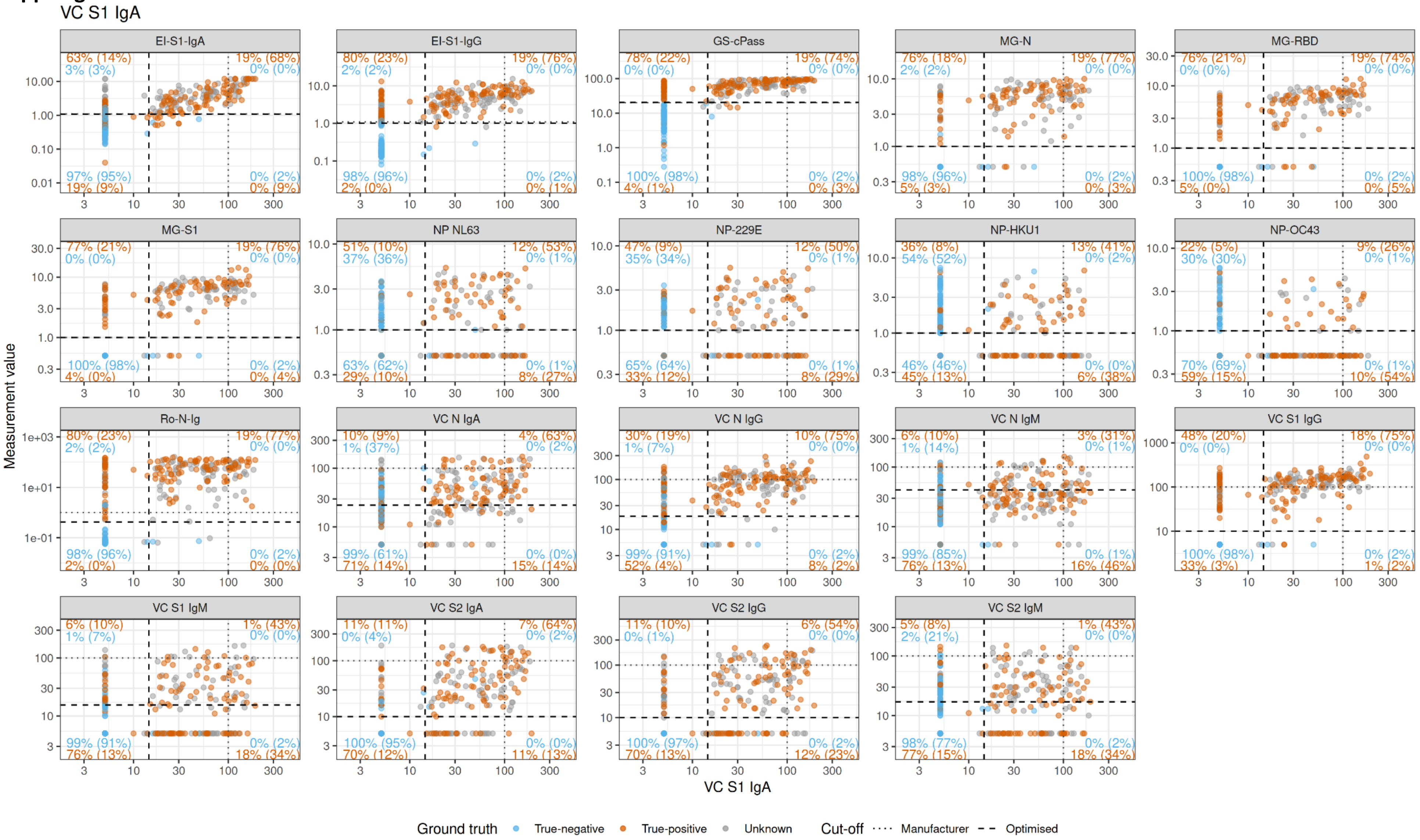

**Suppl.Fig.10G**

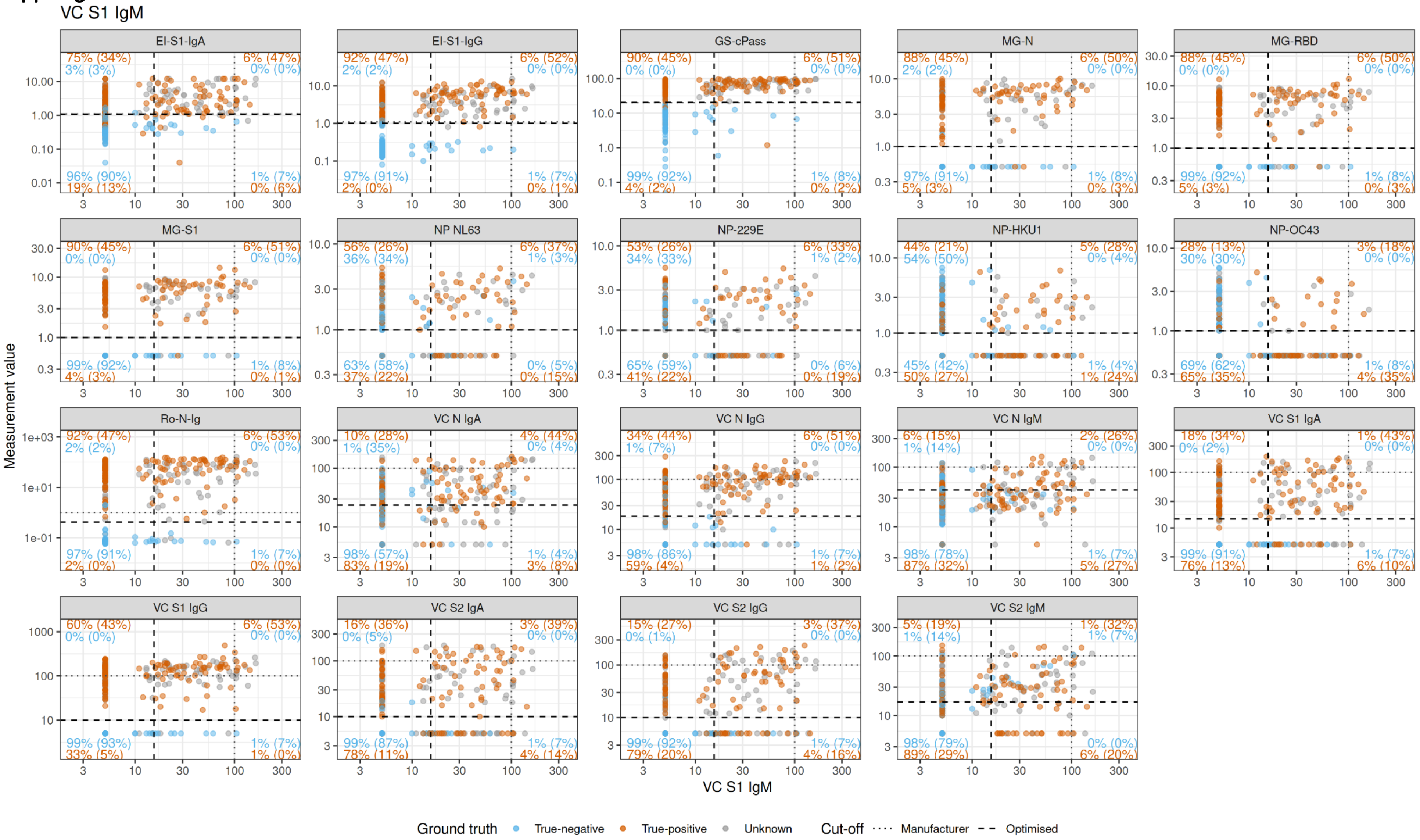

**Suppl.Fig.10H**

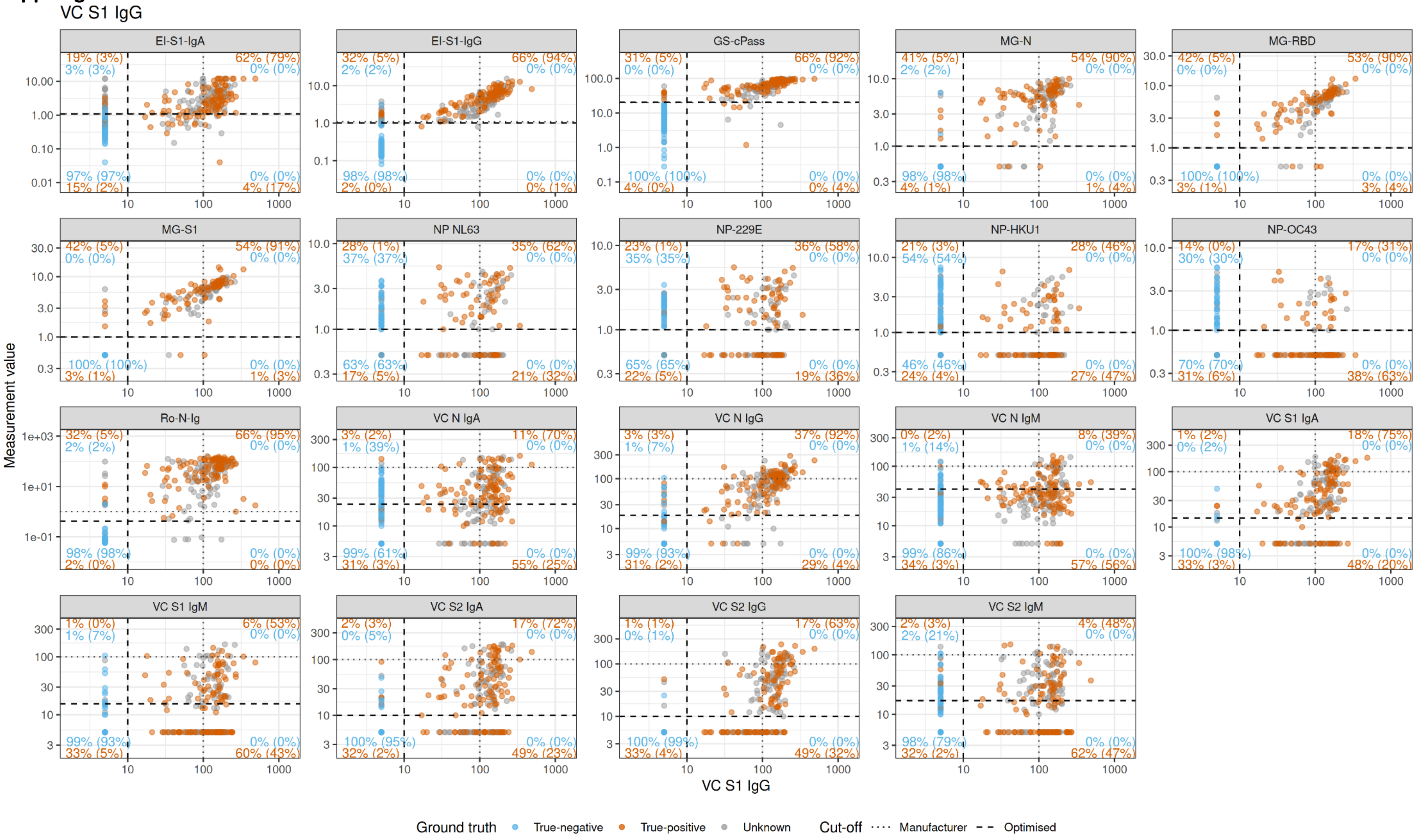

**Suppl.Fig.10I**

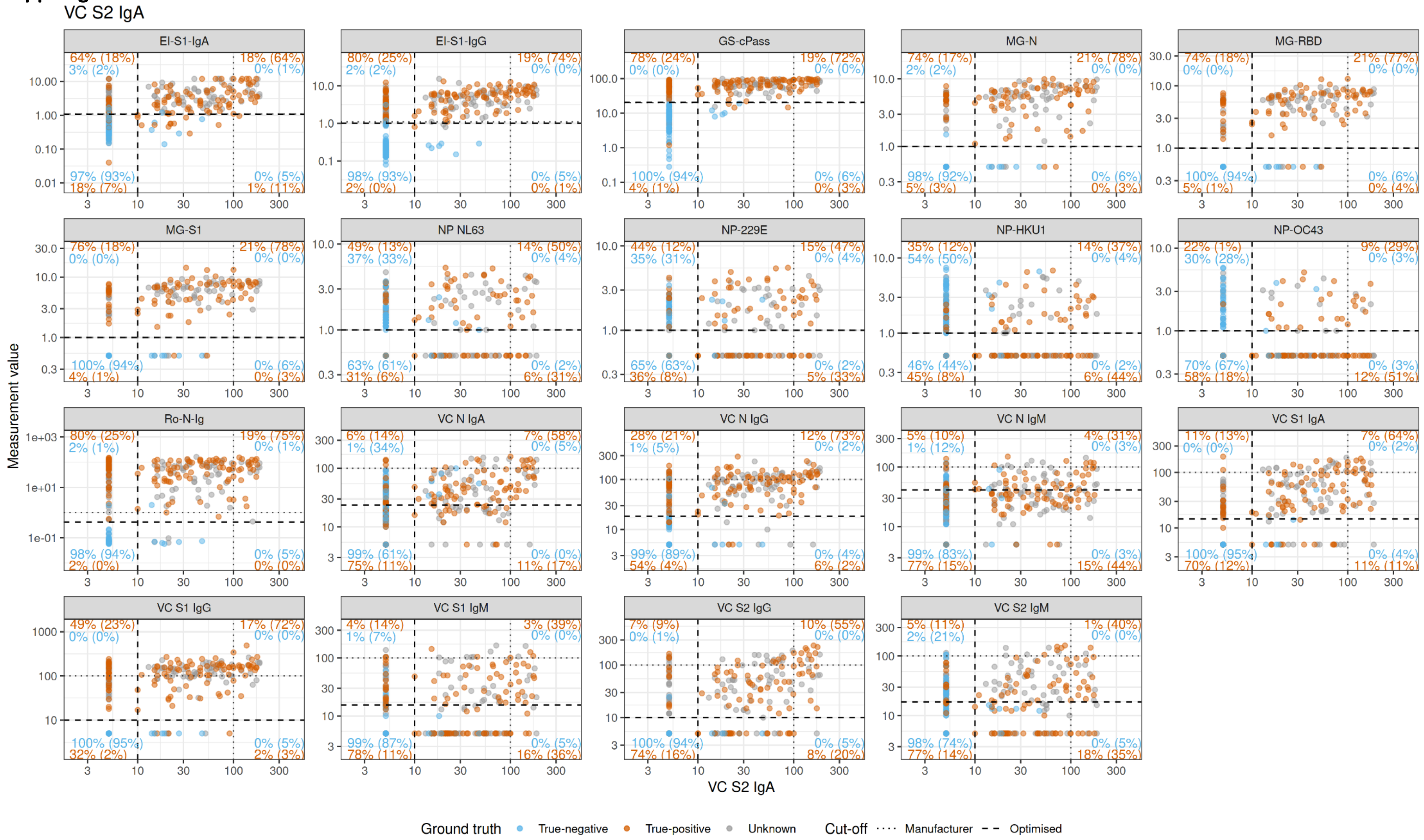

**Suppl.Fig.10J**

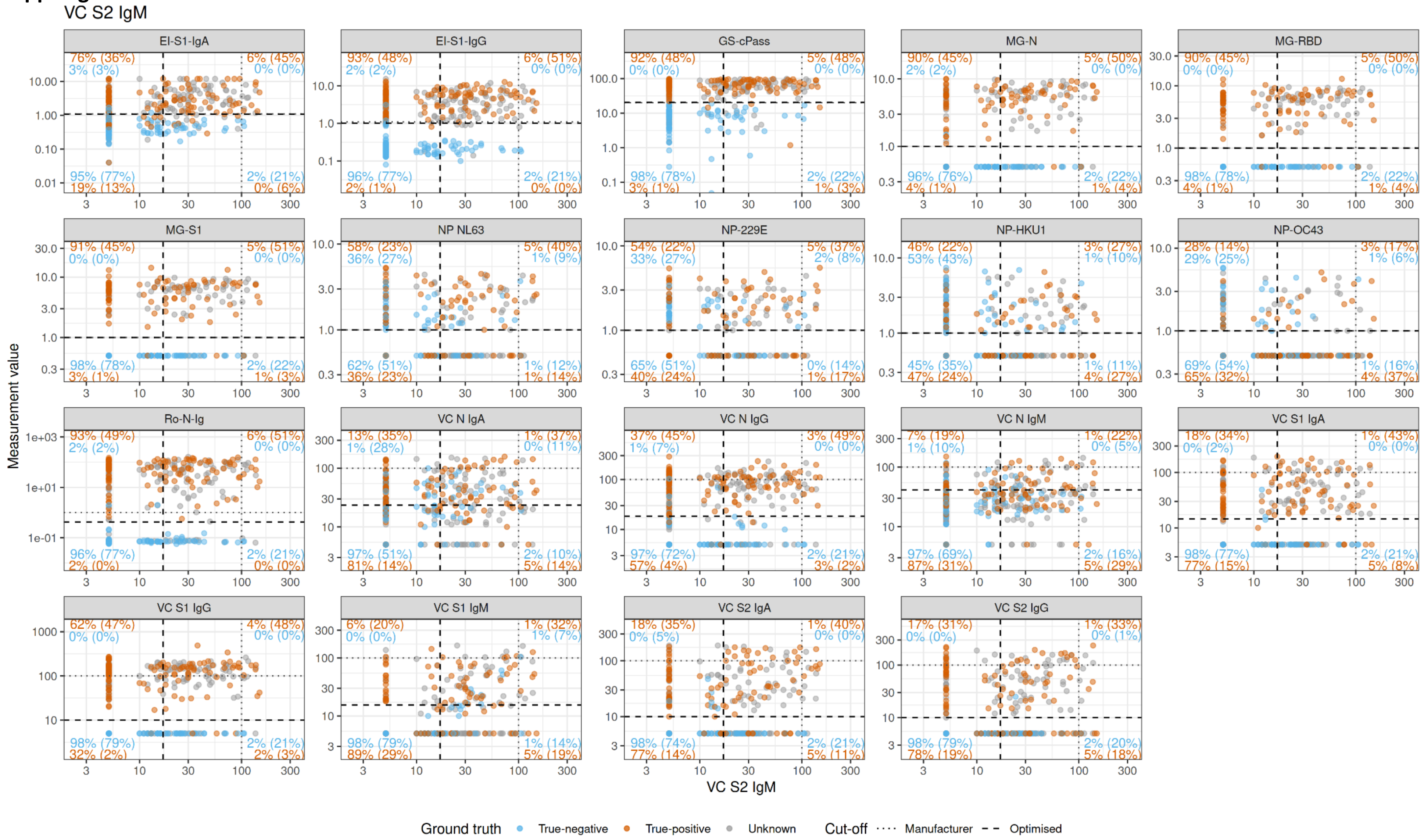

**Suppl.Fig.10K**

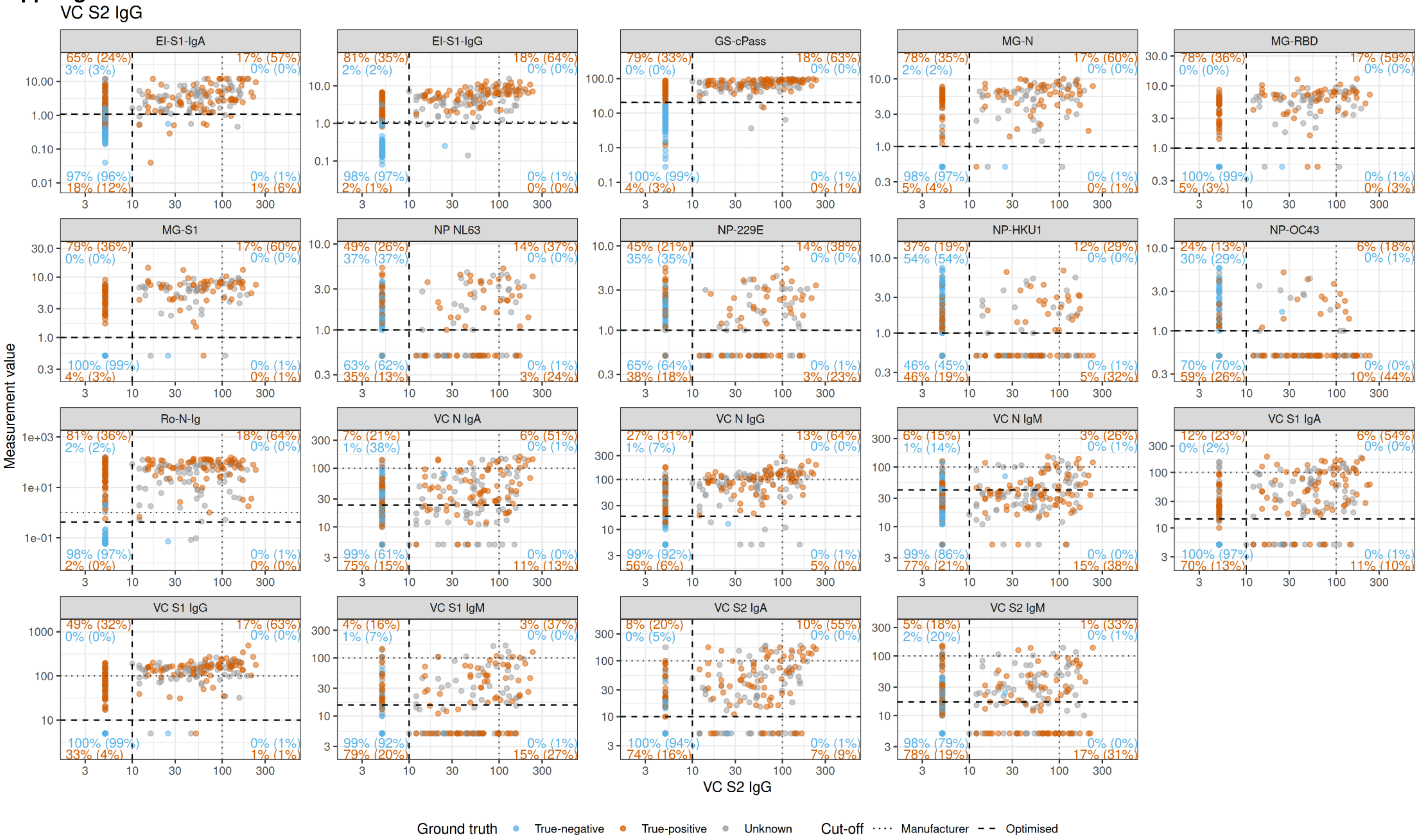

**Suppl.Fig.10L**

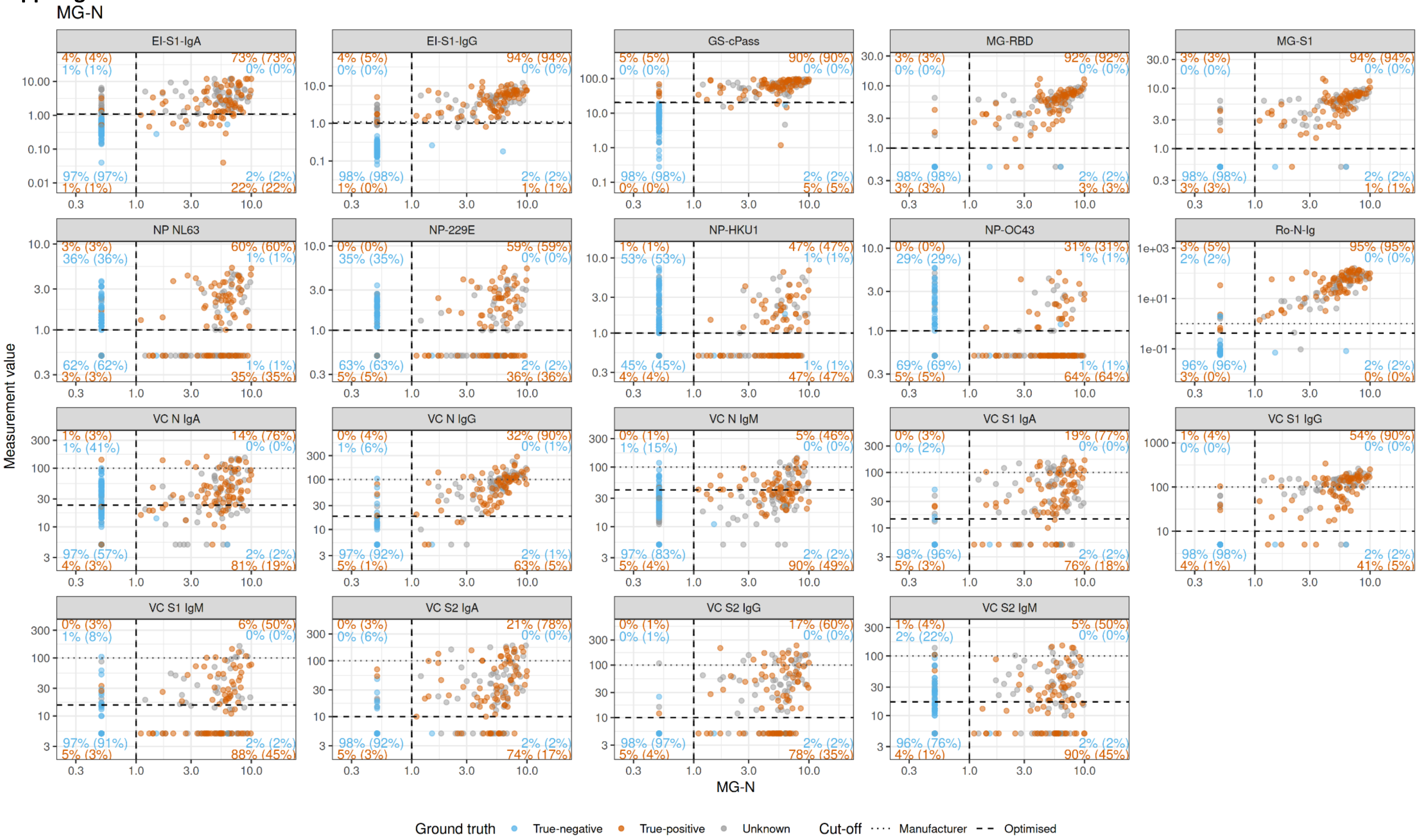

**Suppl.Fig.10M**

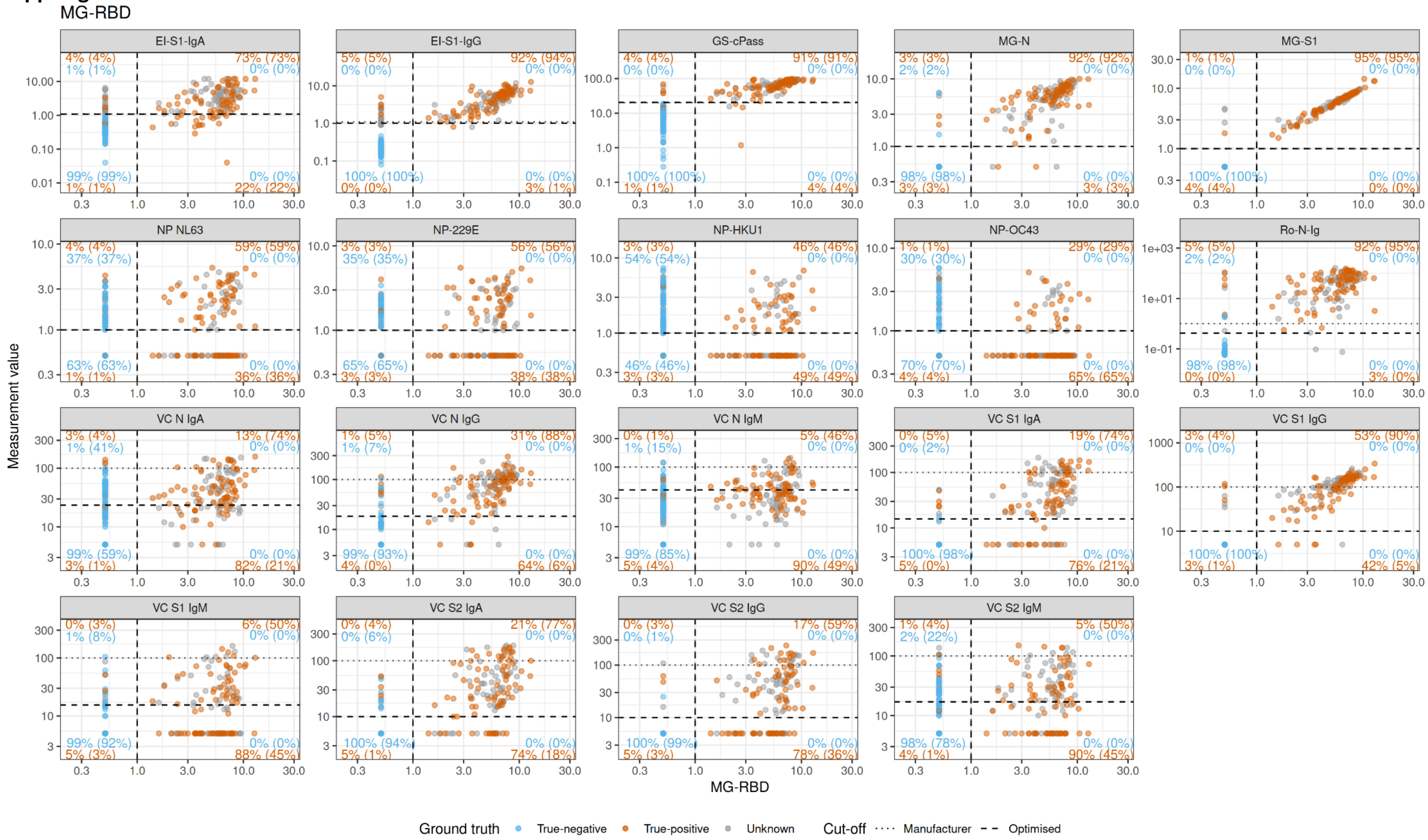

Disclaimer: this is a pre-print manuscript and has not been peer-reviewed yet

**Suppl.Fig.10N**

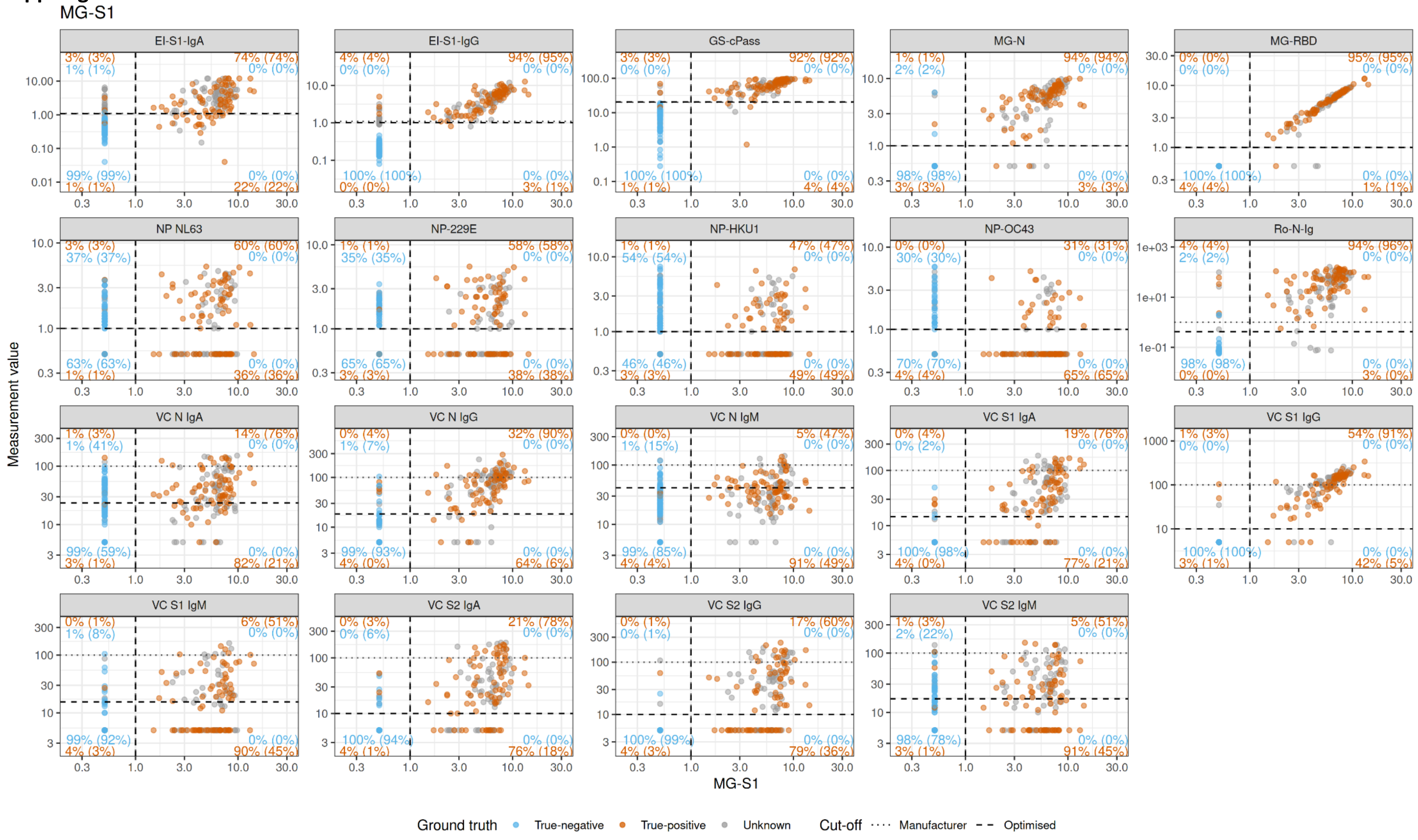

**Suppl.Fig.10O**

NP-OC43

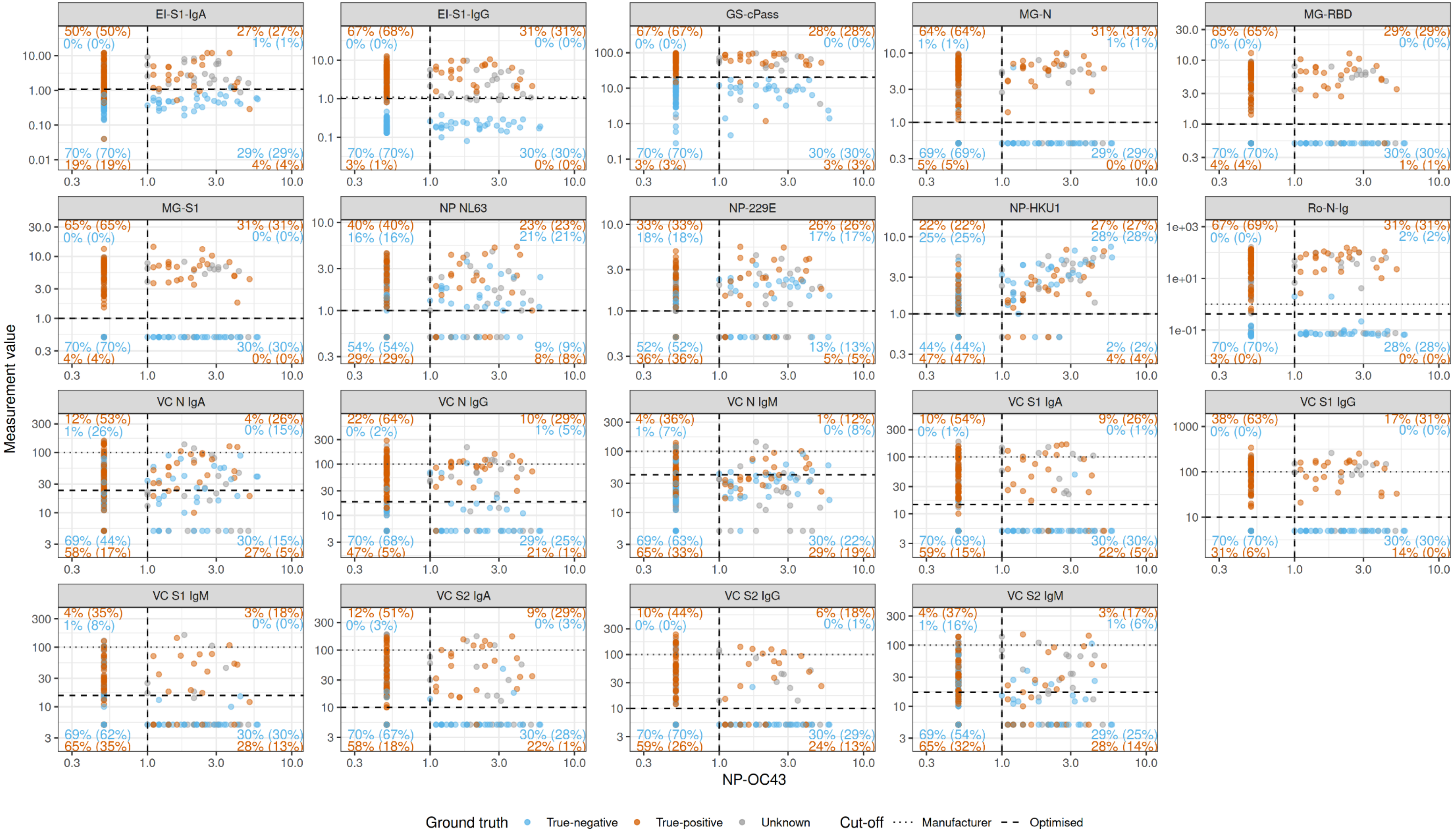

**Suppl.Fig.10P**

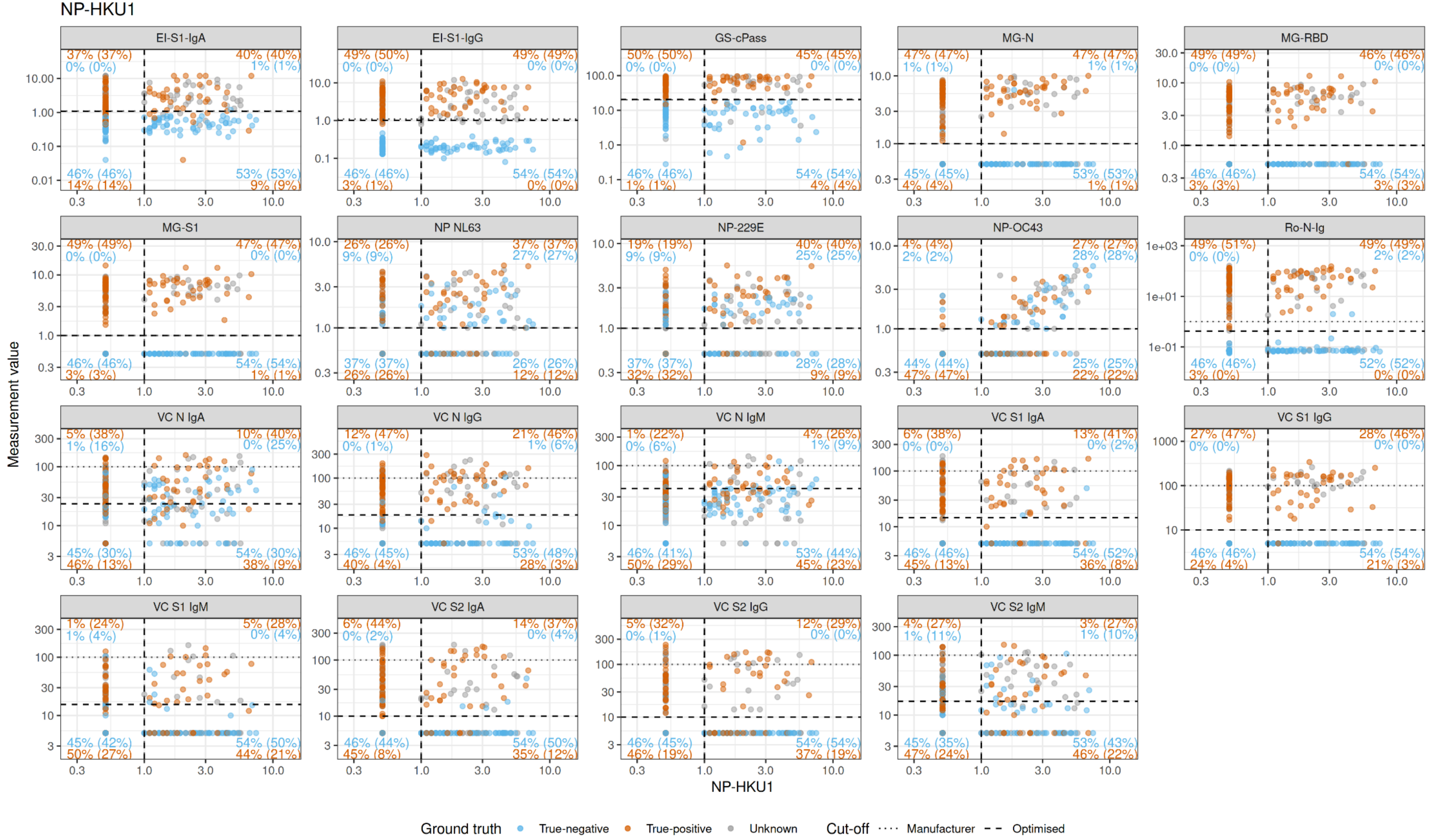

**Suppl.Fig.10Q**

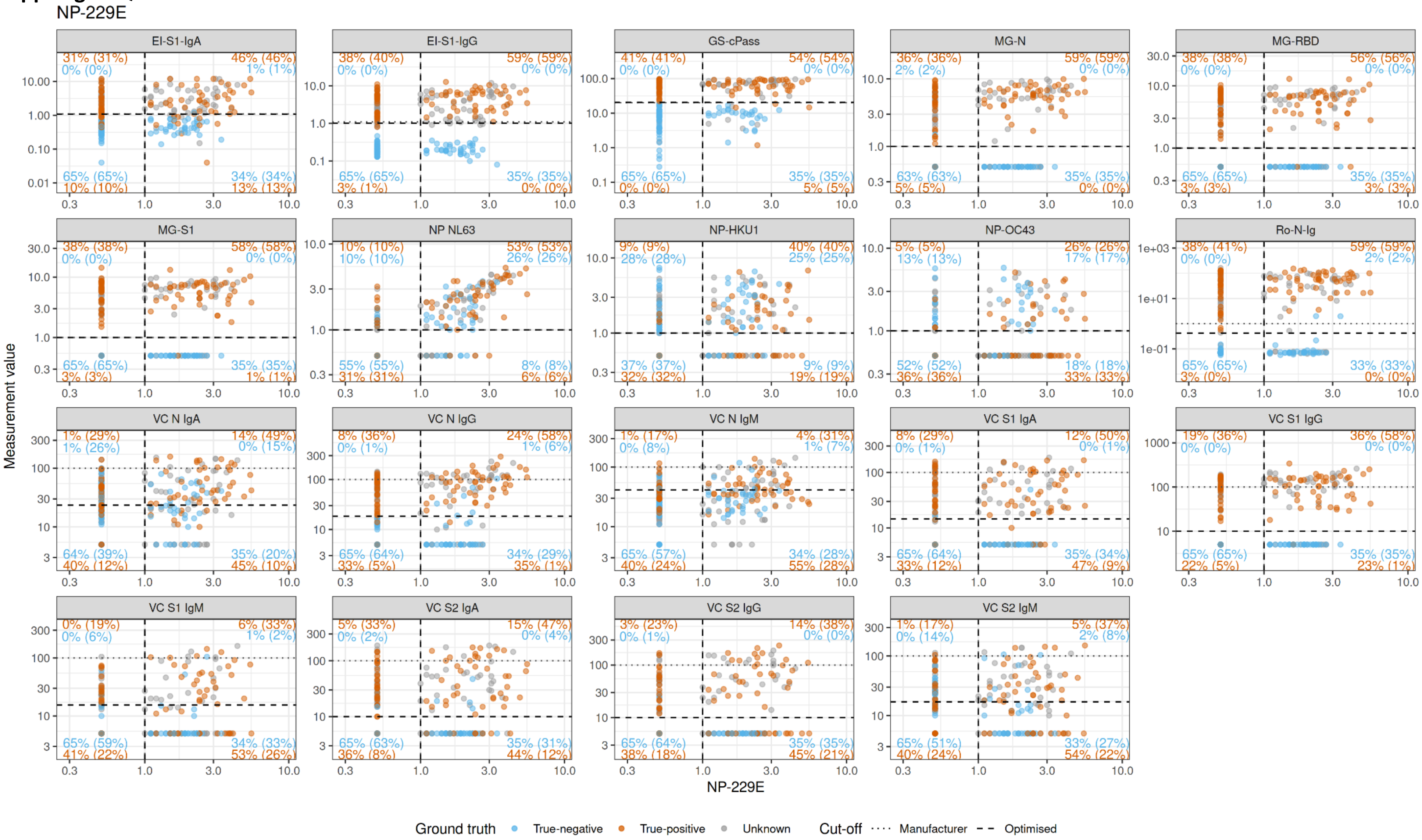

**Suppl.Fig.10R**

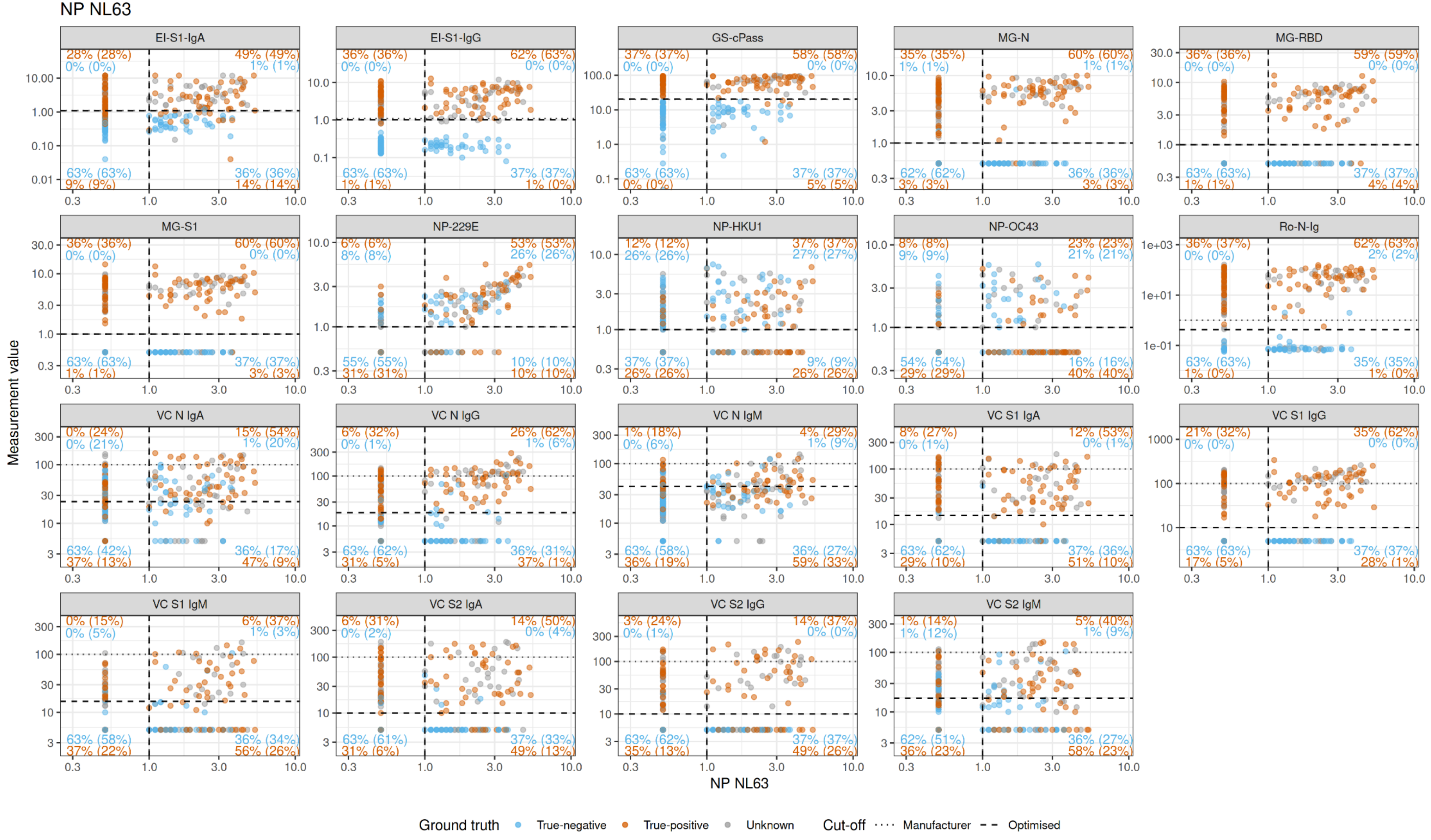

***Supplemental Figure 10: Scatterplots of confirmatory tests vs confirmatory test***

In blue the true-negatives, in orange the true-positives, in grey the values with unknown SARS-CoV-2 status. Black dotted and dashed lines represent the manufacturer's and the optimised positivity cut-offs, respectively. Orange/blue numbers give the percentages of true-positives/-negatives correctly detected by the tests. The orange values above the dotted line represent the percentages of positive test results for the true-positives, the blue number below is the percentage of negatives in the true-negatives.

(A) Values obtained with NT
(B) Values obtained with GS-cPass
(C) Values obtained with VC-N-IgA
(D) Values obtained with VC-N-IgM
(E) Values obtained with VC-N-IgG
(F) Values obtained with VC-S1-IgA
(G) Values obtained with VC-S1-IgM
(H) Values obtained with VC-S1-IG
(I) Values obtained with VC-S2-IgA
(J) Values obtained with VC-S2-IgM
(K) Values obtained with VC-S2-IgG
(L) Values obtained with MG-N
(M) Values obtained with MG-RBD
(N) Values obtained with MG-S1
(O) Values obtained with NP-OC43
(P) Values obtained with NP-HKU1
(Q) Values obtained with NP-229E
(R) Values obtained with NP-NL63
